# Using quantitative bias analysis to adjust for misclassification of COVID-19 outcomes: An applied example of inhaled corticosteroids and COVID-19 outcomes

**DOI:** 10.1101/2024.08.13.24311341

**Authors:** Marleen Bokern, Christopher T. Rentsch, Jennifer K. Quint, Jacob Hunnicutt, Ian Douglas, Anna Schultze

## Abstract

During the pandemic, there was concern that underascertainment of COVID-19 outcomes may impact treatment effect estimation in pharmacoepidemiologic studies. We assessed the impact of outcome misclassification on the association between inhaled corticosteroids (ICS) and COVID-19 hospitalisation and death in the UK during the first pandemic wave using probabilistic bias analysis (PBA).

Using data from Clinical Practice Research Datalink Aurum, we defined a cohort with chronic obstructive pulmonary disease (COPD) on 01 Mar 2020. We compared the risk of COVID-19 hospitalisation and death among users of ICS/long-acting β-agonist (LABA) and users of LABA/LAMA using inverse-probability of treatment weighted (IPTW) logistic regression. We used PBA to assess the impact of non-differential outcome misclassification. We assigned beta distributions to sensitivity and specificity and sampled from these 100,000 times for summary-level and 10,000 times for record-level PBA. Using these values, we simulated outcomes and applied IPTW logistic regression to adjust for confounding and misclassification. Sensitivity analyses excluded ICS+LABA+LAMA (triple therapy) users.

Among 161,411 patients with COPD, ICS users had increased odds of COVID-19 hospitalisations and death compared with LABA/LAMA users (OR for COVID-19 hospitalisation 1.59 (95% CI 1.31 – 1.92), OR for COVID-19 death 1.63, 95% CI 1.26 – 2.11). After IPTW and exclusion of people using triple therapy, ORs moved towards null. All implementations of QBA, both record and summary-level PBA, modestly shifted ORs away from the null and increased uncertainty.

The results provide reassurance that outcome misclassification was unlikely to change the conclusions of the study but confounding by indication remains a concern.

## 2. Introduction

At the beginning of the pandemic, there was concern that people with chronic respiratory diseases may be at increased risk of severe COVID-19 outcomes. However, early data suggested that patients with asthma or chronic obstructive pulmonary disease (COPD) were not substantially overrepresented among COVID-19 deaths or hospitalisations[1].

Inhaled corticosteroids (ICS), anti-inflammatory drugs widely used as maintenance medications in COPD[2], were investigated as a potential repurposed drug treatment for COVID-19, as it was unclear whether ICS were associated with COVID-19 outcomes due to their immunosuppressant effects[1,3]. These effects may have different consequences at different stages of infection[4]. Several observational studies investigated the impact of ICS on COVID-19 outcomes but found inconsistent results that may be affected by biases[5–9]. RCTs consequently suggested a strong protective effect of one inhaled ICS, budesonide, on severe COVID-19 outcomes in patients with mild COVID-19[10,11].

Many observational studies considered the role of potential measured and unmeasured confounding in detail[5–8], but the potential role of misclassification has received less attention to date. Outcome misclassification is likely to have occurred during the rollout and use of new ICD-10 codes denoting COVID-19 diagnoses particularly in the first wave of the pandemic[12,13]. In particular, there was concern that COVID-19 mortality would be underestimated by about one third when considering official reports of deaths registered as due to COVID-19 as opposed to excess all-cause mortality[13].

This study aims to assess the impact of ICS/LABA (long-acting β-agonist) use compared with LABA/LAMA (long-acting muscarinic antagonist) use on the risk of hospitalisation or death due to COVID-19 during the first wave of COVID-19 in the UK (March 2020 – August 2020). We conducted quantitative bias analysis (QBA) to measure the potential impact of outcome misclassification on the effect estimates.

## 3. Methods

The study protocol was registered on ENCEPP EU PAS (register number 47885), and we completed the RECORD-PE checklist (S1.1)[14].

### Study design

#### Data source

This study used data routinely collected patient data from primary care in the UK recorded in the Clinical Practice Research Datalink (CPRD) Aurum. CPRD Aurum includes data on 41 million patients (May 2022 build), with over 13 million patients currently registered (20% of the UK population)[15] from over 1,300 GP practices which use EMIS GP patient management software[15]. CPRD Aurum has been shown to be broadly representative of the English population in terms of age, gender, geographical spread and deprivation[16].

CPRD Aurum was linked to Hospital Episode Statistics (HES) Admitted Patient Care (APC) and Office for National Statistics (ONS) Death Registry[16,17]. HES APC holds information on all in-patient contacts at NHS hospitals in England[17,18]. The ONS death registry contains information on deaths occurring in England and Wales, including a cause of death documented using International Classification of Disease 10^th^ revision (ICD-10) codes[18,19]. Data was also linked to the Index of Multiple Deprivation (IMD), a postcode-level indicator of socioeconomic status.

#### Study population

We defined a cohort of people with a code for COPD before 01^st^ March 2020 (i.e, the index date) based on a validated algorithm to identify COPD in CPRD[20]. Patients had to be alive, aged 35 or older, and registered in CPRD Aurum on 01^st^ March 2020. Included patients needed to have 12 months’ continuous registration before the index date and a record of current or former smoking at any point before the index date. In the main analysis, we excluded people with asthma diagnoses recorded within three years before the index date, leukotriene receptor antagonist use within four months before the index date as this indicates asthma, or other chronic respiratory disease at any point before the index date. Patients were followed up until death (recorded in ONS or CPRD), deregistration, or 31^st^ August 2020, whichever came first. If death was registered in ONS, we used that date as the date of death. If death was missing in ONS but registered in CPRD, we used the date recorded in CPRD as the date of death. A study diagram[21] can be found in S1.2.

##### Exposure

Continuous treatment episodes were generated based on the recorded prescription issue date and information on the intended duration, prescribed amount, and dosage (S1.3).

We used the derived start and end dates for treatment exposures to identify people exposed to ICS/LABA or LABA/LAMA on the index date either as combined or separate inhalers. ICS/LABA was the exposure of interest and LABA/LAMA was the active comparator. People using ICS+LABA+LAMA (henceforth referred to as triple therapy) were included in the ICS/LABA group but were excluded in a sensitivity analysis.

##### Outcome

The outcomes were i) hospitalisation with a primary diagnosis code for COVID-19 as recorded in HES APC, and ii) death with a diagnosis code for COVID-19 as a cause of death anywhere on the death certificate recorded in the ONS Death Registry. Diagnostic codes for both COVID-19 hospitalisation and death were ICD-10 U07.1 and U07.2.

#### Covariates

Results were adjusted for the following baseline covariates: age, gender, BMI (most recent and within the previous 10 years, categorised as underweight (<18.5), normal weight (18.5-24.9), overweight (25-29.9), or obese (≥30)), smoking (current vs former), ethnicity, cancer (ever), diabetes (ever), chronic kidney disease (ever), cardiovascular disease (ever), hypertension (ever), asthma (not within the past 3 years), immunosuppression, receipt of influenza vaccine (past year), receipt of pneumococcal vaccine (past 5 years), index of multiple deprivation quintile (IMD), and COPD exacerbation in the past 12 months. COPD exacerbations were identified based on a validated algorithm using a combination of codes for COPD exacerbations, lower respiratory tract infections, respiratory symptoms, and antibiotic or oral corticosteroid prescriptions[22].

### Statistical analyses

We summarised the characteristics of the cohort using descriptive statistics by exposure group. There were missing data for BMI, ethnicity and IMD. Missing BMI values were assumed to be normal BMI in line with previous work[23]. Missing ethnicity and IMD were treated as a separate category.

We estimated propensity scores (PSs) and used inverse probability of treatment weighting (IPTW) to estimate the average treatment effect (ATE) adjusting for potential confounders. PSs were estimated using logistic regression including the covariates listed above. Weights were calculated as 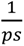 (ICS) and 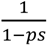 (LABA/LAMA), where the PS is the probability of receiving ICS. Overlap of the PSs across treatment groups was assessed graphically and by summarising by treatment group. PSs were trimmed to the region of common support[24].

Logistic regression was used to estimate odds ratios (ORs) and 95% confidence intervals (CIs) for the association between exposure and the outcomes. This was done to make estimates comparable across analyses, as simple bias analysis (SBA) and summary-level probabilistic bias analysis (PBA) are conducted based on 2x2 table, generating relative risks or ORs as relative effect estimates. Additionally, follow-up was short and censoring low (S1.6), so time-to-event was not taken into account in this analysis.

### 3.1. Quantitative bias analysis

#### Quantitative bias analysis for outcome misclassification

We used SBA and PBA[25] to investigate the potential impact of outcome misclassification using methods based on Fox et al[26]. We used estimates of sensitivity and specificity as bias parameters to describe our assumptions about the accuracy of COVID-19 hospitalisation as recorded in HES, and COVID-19 death as recorded in the ONS death registry. We assumed that outcome misclassification was non-differential with respect to the exposure, as hospitalisations and deaths would have been coded without knowledge of the exposure. This also implies that exposure is not associated with characteristics that may predict outcome validity. We assumed the occurrence and dates recorded in HES and ONS to be correct, and corrected only for misclassification of recorded causes of hospitalisations and deaths. Therefore, correction for outcome misclassification was conducted only among people who were hospitalised or died of any cause. This was done by conducting the QBA steps in a reduced dataset including only patients hospitalised or who died; patients without a hospitalisation or death were consequently added back in to calculate the effect estimates in the entire population. Code illustrating these steps is available on our GitHub repository.

Initially, we conducted SBA using available tools[27] and bias formulas using best estimates of sensitivity and specificity. We then performed both summary-level and record-level PBA that involved Monte-Carlo sampling of the bias parameter values from prespecified distributions for both outcomes. PBA, while more analytically and computationally complex than SBA, offers several advantages, including the ability to incorporate uncertainty regarding the values of bias parameters, and analytical flexibility. The simulation interval (SI) generated during PBA incorporates the systematic error arising from the modelled bias, the random error arising from the misclassification process, and the conventional random error. A full description of the method is available in S4.4 and S4.5.

#### Bias parameter values and distributions

##### Hospitalisations

In the absence of direct evidence from UK, values for sensitivity and specificity of hospital diagnoses were estimated based on validation studies conducted in North America and bounds given by the data[28,29]. Kadri et al. estimated a sensitivity of 98.01% (97.63% – 98.39%), and a specificity of 99.04% (98.95% - 99.13%) in an administrative database in the US during April and May 2020. The reference was SARS-CoV-2 PCR test results[28]. A validation study conducted in Alberta, Canada using hospital admissions between March 2020 and February 2021 estimated a sensitivity of ICD-10 code U07.1 of 82.5% (81.8% - 83.2%) when using SARS-CoV-2 PCR test results as the gold-standard[29]. Based on these validation studies, we estimated that sensitivity would have a median of 0.90 and lie between 0.80 and 0.96. The 2x2 table indicated that specificity had a lower bound of 95%, as only 5% of patients with a hospitalisation had a COVID-19 hospitalisation, so the proportion of false-positive COVID-19 hospitalisations could not exceed 5%. We assigned parameter values to beta distributions based on the mean and variance of the target distributions[30]. For sensitivity, we parameterised a beta distribution with median = 0.90, 2.5th percentile = 0.80 and 97.5th percentile = 0.96 (α = 47.7, β = 5.5). For specificity, we used a beta distribution with median = 0.98, 2.5^th^ percentile = 0.95, 97.5^th^ percentile = 0.999 (α = 1, β = 1, transformed to have a lower bound of 0.95). These distributions resulted in negative cell counts in the 2x2 table in approximately 28% of iterations, indicating incompatibility with the data[30]. As the negative cell counts were driven by false-positive COVID-19 hospitalisations (i.e., specificity), we subsequently increased the lower bound of the specificity distribution to 0.97 in order to obtain plausible results.

##### Deaths

For COVID-19 deaths, data on excess deaths[31] and reported COVID-19 deaths in England and Wales[32] was used to inform estimates of sensitivity and specificity. Assuming that all excess deaths in the UK between 01 March 2020 and 07 May 2020 were true COVID-19 deaths, we estimated a sensitivity of 72.2% and a specificity of 96.6% among those who died of any cause, as recorded in ONS or CPRD. The calculations of the parameter values can be found in S4.2. We assumed that the sensitivity and specificity estimates would be within a 10% range of the point estimate, and we parameterised a beta distribution for sensitivity with values median = 0.72, 92.5^th^ percentile = 0.60, 97.5^th^ percentile = 0.83 (α = 39.6, β = 15.4). For specificity, we used a beta distribution with median = 0.96, 2.5^th^ percentile = 0.91, 97.5^th^ percentile = 0.99 (α = 91.2, β = 3.9). Graphs of all bias parameter distributions can be found in S4.3.

**Table 1.**
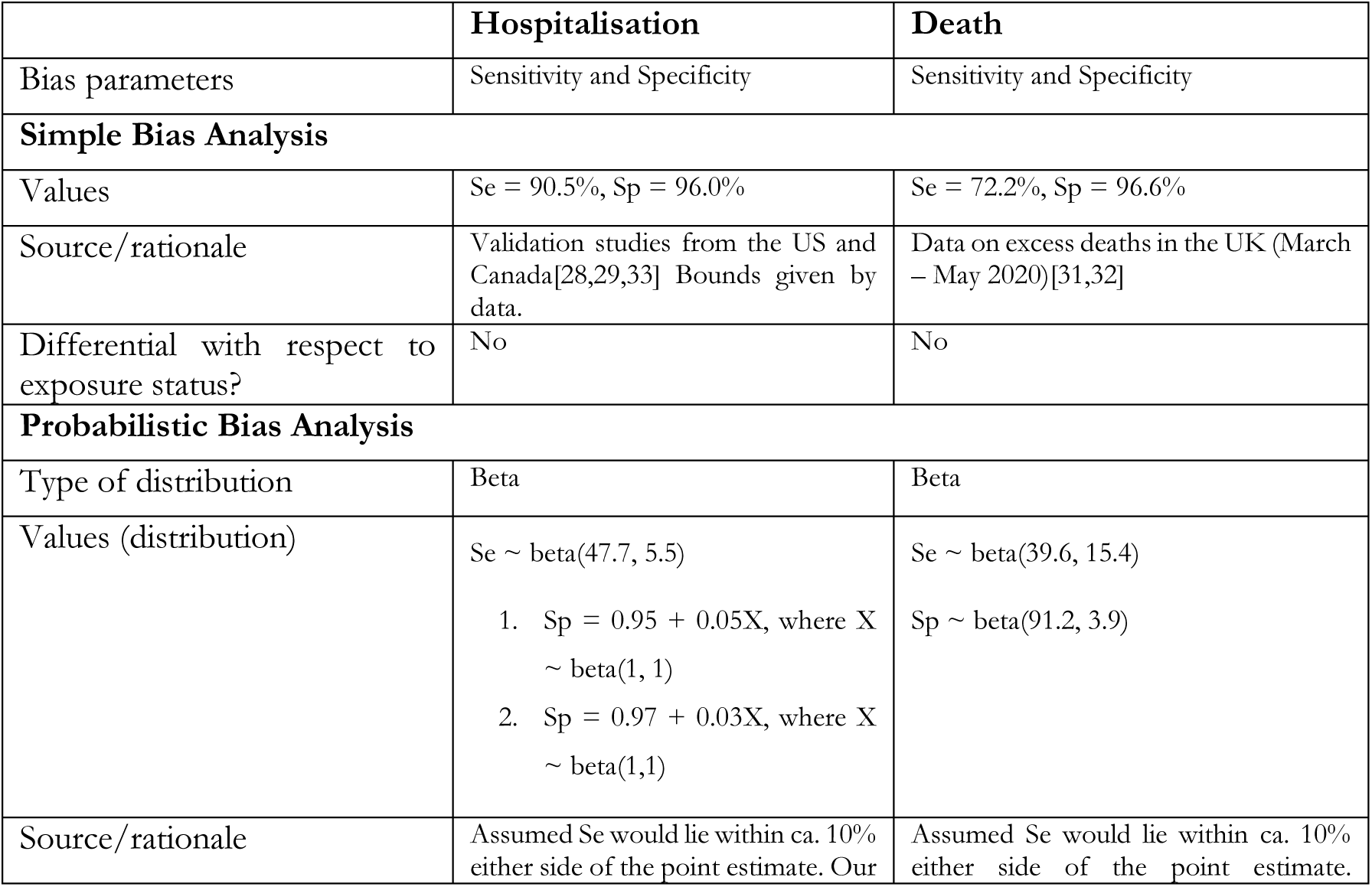
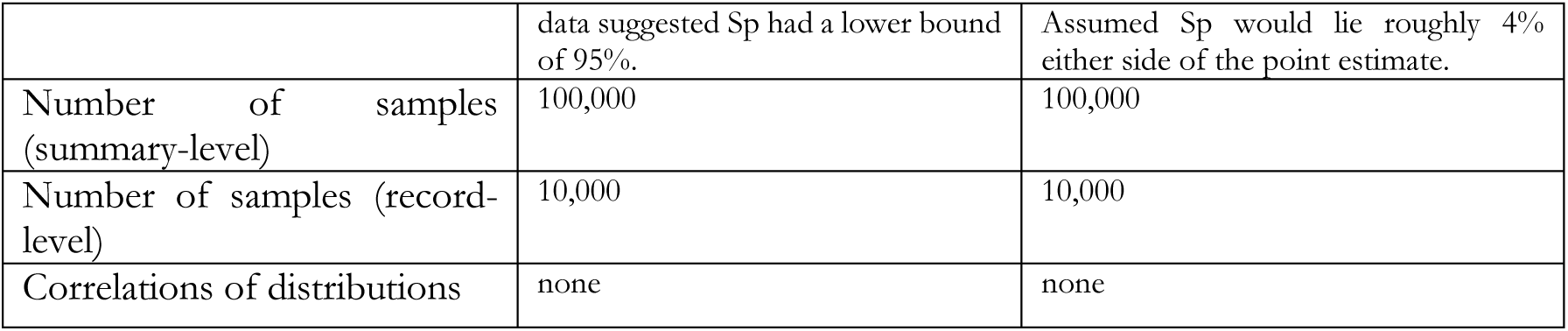
Decisions related to the implementation of quantitative bias analysis.

##### Quantitative bias analysis

For QBA, we constructed 2x2 tables for the relevant population by restricting to people who were hospitalised or died of any cause. Using available tools[27], we applied the best estimates of the bias parameters (Se = 90.5%, Sp = 96.0% for COVID-19 hospitalisations, Se = 72.2%, Sp = 96.6% for COVID-19 deaths) to conduct SBA. For summary-level PBA, we then sampled from the specified distributions 100,000 times, applied the sampled values to correct the 2x2 tables, and calculated the bias-adjusted effect estimate and the 95% SI. Patients with multiple hospitalisations during the follow-up period were counted as having a COVID-19 hospitalisation if any of those hospitalisations were recorded as being due to COVID-19. If none of the hospitalisations were recorded as due to COVID-19, we only counted that patient once. Therefore, the 2x2 table contained individual patients rather than individual hospitalisations. For record-level correction, we sampled from each distribution 10,000 times and corrected the outcome for each individual record following the steps described in Fox et al[26].

After each iteration within the simple, summary and record-level correction that was conducted on a dataset restricted to people who were hospitalised or died of any cause, we added back in patients without hospitalisations or deaths to conduct both unweighted and IPT-weighted logistic regression on the entire population, generating bias-adjusted ORs with 95% SIs. As patients could have multiple hospitalisations during the study period, we simulated the potential outcome misclassification for each hospitalisation, and selected the first one that was simulated as being due to COVID-19 to determine the outcomes.

##### Sensitivity analyses

To improve comparability of the treatment groups, we excluded people using triple therapy, as we expected they would be sicker than those using dual therapy. To assess the robustness of the results with respect to the definition of exposure, we changed the definition of the discontinuation date to 6 months after the last prescription issue date, not taking into account estimated treatment duration. We included people who met some of the exclusion criteria, i.e., people with COPD who also had asthma or other respiratory diseases, to increase the power of the analyses.

There were some deaths registered in CPRD that were not registered in ONS (∼10% of deaths). CPRD does not record a cause of death. Therefore, we performed an analysis where we classed those missing causes of death as recorded as due to COVID-19 to assess whether the classification of those deaths would have affected our results.

We repeated the summary-level PBA to simulate differential misclassification of COVID-19 deaths by varying the sensitivity among the treatment groups between 0.6 and 1, assuming that causes of death were more likely to be misclassified among people using triple therapy if they were more likely to die outside of hospital, e.g., in care homes. If the misclassification were differential, we assumed this would affect sensitivity more than specificity (i.e., some cases may be missed), so we used a fixed specificity of 0.97 and varied the sensitivity.

Data was managed using Stata version 17.0[34] and analysis carried out using R (version 4.3.3)[35]. Code lists and data management and analysis code can be found on our GitHub repository (https://github.com/bokern/ics_covid).

## 4. Results

### Description of the study population

The cohort included 161,411 patients with COPD (flowchart in S1.4). Of those, 56,059 (35.37%) were using ICS/LABA at baseline, and 22,319 (13.81%) were using LABA/LAMA. For both outcomes and treatment groups, the median follow-up time was 183 days, the length of the study period. The proportion of patients censored for any reason was 6.45% for COVID-19 hospitalisations and 6.06% for COVID-19 deaths (S1.6).

Cohorts were similar in all measured covariates apart from proportion of people with a history of asthma, and COPD exacerbations in the past year, both of which were higher in the ICS group than the LABA/LAMA group (Table 2). After applying IPTWs, treatment groups were balanced on all covariates.

**Table 2.** Baseline demographic and clinical characteristics of patients in the cohort before and after inverse probability of treatment weighting.

|  | Before weighting |  |  | After inverse probability of treatment weighting |  |  |
| --- | --- | --- | --- | --- | --- | --- |
|  | ICS<br>N = 56,059 | LABA/LAMA<br>N = 22,319 | SMD | ICS<br>N = 56051 | LABA/LAMA<br>N = 22324 | SMD |
| Age |  |  |  |  |  |  |
| Mean (SD) | 71.3 (10.5) | 70.8 (10.2) | 0.0480 | 71.18 (10.46) | 71.19 (10.30) | -0.00170 |
| Median<br>(25%-75%) | 71.7<br>(64.7-78.7) | 71.7<br>(63.7-77.7) |  | 71.67<br>(63.67-78.67) | 71.67<br>(64.67-78.67) |  |
| Gender |  |  |  |  |  |  |
| Male | 29,830 (53%) | 12,245 (55%) |  | 30,094 (54%) | 12,036 (54%) |  |
| Female | 26,229 (47%) | 10,074 (45%) | 0.0166 | 25,957 (46%) | 10,287 (46%) | 0.00226 |
| BMI |  |  |  |  |  |  |
| Underweight<br>(<18.5) | 3,142 (5.6%) | 970 (4.3%) | 0.0126 | 17,928 (32%) | 7,112 (32%) | 0.000344 |
| Normal (18.5-24.9) | 18,163 (32%) | 6,926 (31%) | 0.0136 | 2,936 (5.2%) | 1,162 (5.2%) | 0.00126 |
| Overweight (25-29.9) | 17,349 (31%) | 7,172 (32%) | -0.0118 | 17,529 (31%) | 6,979 (31%) | 9.41E-05 |
| Obese (>=30) | 17,405 (31%) | 7,251 (32%) | -0.0144 | 17,658 (32%) | 7,071 (32%) | -0.00170 |
| Ethnicity |  |  |  |  |  |  |
| White | 49,390 (88%) | 19,584 (88%) | 0.00370 | 49,334 (88%) | 19,648 (88%) | 3.14E-05 |
| South Asian | 742 (1.3%) | 197 (0.9%) | 0.00423 | 665 (1.2%) | 280 (1.3%) | -0.000678 |
| Black | 351 (0.6%) | 130 (0.6%) | 0.000438 | 345 (0.6%) | 140 (0.6%) | -9.71E-05 |
| Mixed | 142 (0.3%) | 49 (0.2%) | 0.000338 | 138 (0.2%) | 61 (0.3%) | -0.000276 |
| Unknown | 5,434 (9.7%) | 2,359 (11%) | -0.00870 | 5,569 (9.9%) | 2,195 (9.8%) | 0.00102 |
| Smoking |  |  |  |  |  |  |
| Current<br>smoking | 22,763 (41%) | 10,073 (45%) |  | 23,489 (42%) | 9,335 (42%) |  |
| Former<br>smoking | 33,296 (59%) | 12,246 (55%) | 0.0452 | 32,562 (58%) | 12,989 (58%) | -0.000910 |
| Index of Multiple<br>Deprivation |  |  |  |  |  |  |
| 1 | 7,178 (13%) | 3,048 (14%) | -0.00846 | 7,310 (13%) | 2,891 (13%) | 0.000934 |
| 2 | 9,213 (16%) | 3,813 (17%) | -0.00649 | 9,321 (17%) | 3,730 (17%) | -0.000787 |
| 3 | 9,971 (18%) | 4,077 (18%) | -0.00480 | 10,051 (18%) | 4,022 (18%) | -0.000835 |
| 4 | 12,722 (23%) | 5,012 (22%) | 0.00235 | 12,680 (23%) | 5,061 (23%) | -0.000502 |
| 5 | 16,945 (30%) | 6,357 (28%) | 0.0174 | 16,659 (30%) | 6,609 (30%) | 0.00114 |
| Missing | 30 (<0.1%) | 12 (<0.1%) | -2.44E-06 | 29 (<0.1%) | 11 (<0.1%) | 5.40E-05 |
| Diabetes | 14,081 (25%) | 5,517 (25%) | 0.00397 | 14,021 (25%) | 5,617 (25%) | -0.00146 |
| Hypertension | 28,603 (51%) | 11,319 (51%) | 0.00311 | 28,563 (51%) | 11,415 (51%) | -0.00173 |
| Cardiovascular<br>disease | 16,772 (30%) | 6,540 (29%) | 0.00618 | 16,682 (30%) | 6,665 (30%) | -0.000911 |
| Cancer | 10,543 (19%) | 4,416 (20%) | -0.00974 | 10,697 (19%) | 4,270 (19%) | -0.000435 |
| Past asthma | 15,346 (27%) | 2,665 (12%) | 0.154 | 12,875 (23%) | 5,141 (23%) | -0.000602 |
| Kidney impairment | 16,724 (30%) | 6,738 (30%) | -0.00352 | 16,800 (30%) | 6,758 (30%) | -0.00300 |
| Immunosuppression | 665 (1.2%) | 277 (1.2%) | -0.000547 | 674 (1.2%) | 268 (1.2%) | 1.33E-06 |
| Influenza vaccine | 44,905 (80%) | 17,962 (80%) | -0.00373 | 44,954 (80%) | 17,898 (80%) | 0.000252 |
| Pneumococcal vaccine | 5,985 (11%) | 3,237 (15%) | -0.0382 | 6,598 (12%) | 2,636 (12%) | -0.000376 |
| COPD exacerbation in past 12 months | 22,809 (41%) | 6,222 (28%) | 0.128 | 20,759 (37%) | 8,290 (37%) | -0.00100 |
| BMI = Body Mass Index, ICS = Inhaled corticosteroid, LABA = Long-acting β-agonist SD = Standard Deviation, SMD = Standardised Mean Difference | | | | | | |

### Association between ICS and clinical outcomes

662 COVID-19 hospitalisations and 366 COVID-19 deaths occurred in the study period Table *3*). In unadjusted models, people using ICS were at increased risk of all outcomes compared with people using LABA/LAMA (OR for COVID-19 hospitalisation 1.59 (95% CI 1.31 – 1.92), OR for COVID-19 death 1.63 (95% CI 1.26 – 2.11)) (**Fig. *1***). After IPTW, ORs shifted slightly towards the null (OR for COVID-19 hospitalisation 1.46 (95% CI 1.21 – 1.76), OR for COVID-19 death 1.42 (95% CI 1.11 – 1.82)). We also observed an increased risk of all-cause death among ICS users (OR 1.38 (95% CI 1.26 – 1.52)), which attenuated towards the null after IPTW (OR 1.23 (95% CI (1.12 – 1.34)) (results not shown).

**Fig. 1.**
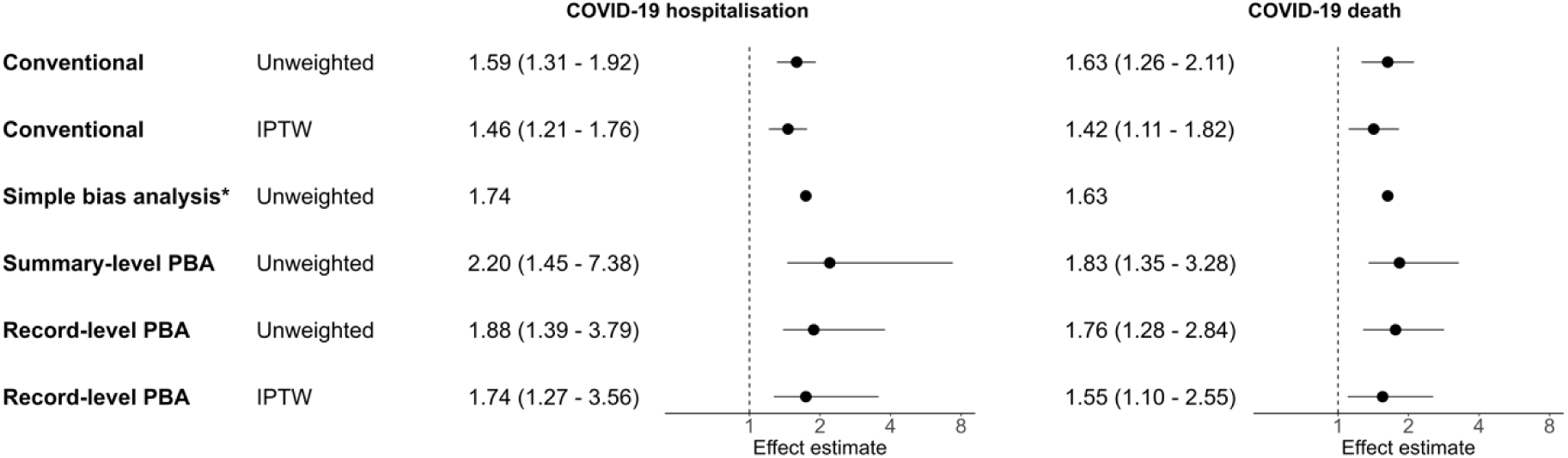
Forest plot of odds ratios and 95% confidence or simulation intervals for COVID-19 hospitalisations and deaths, comparing ICS/LABA (+/- LAMA) users to LABA/LAMA users. Effect estimates >1 indicate an increased risk in the ICS group. Conventional analysis refers to logistic regression without quantitative bias analysis. IPTW = Inverse probability of treatment weighting, PBA = probabilistic bias analysis. * Simple bias analysis only results in a point estimate

**Fig. 2.**
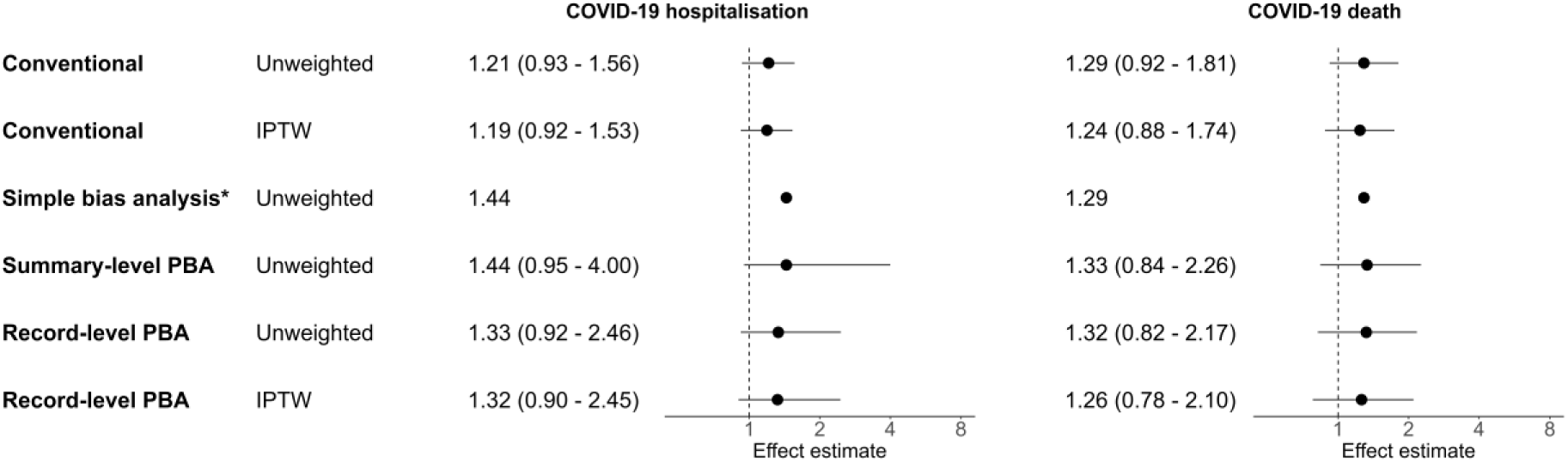
Forest plot of odds ratios and 95% confidence or simulation intervals for COVID-19 hospitalisations and deaths, comparing ICS/LABA users to LABA/LAMA users, excluding triple therapy users. Effect estimates >1 indicate an increased risk in the ICS group. Conventional analysis refers to logistic regression without quantitative bias analysis. IPTW = Inverse probability of treatment weighting, PBA = probabilistic bias analysis. *Simple bias analysis only results in a point estimate

**Table 3.**
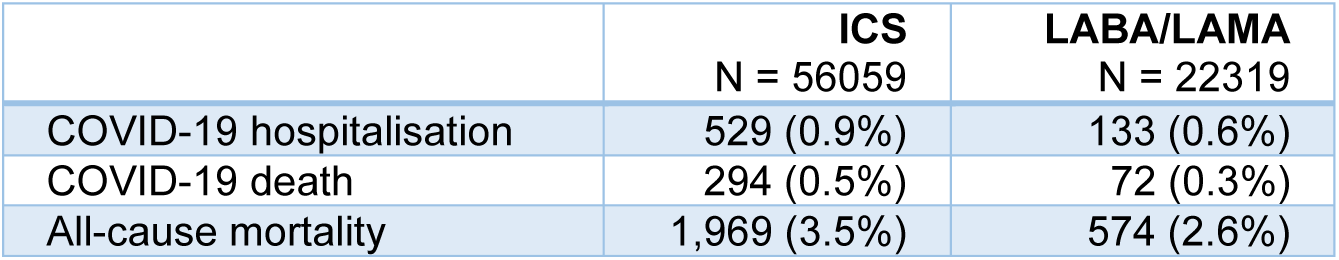
Observed outcomes by treatment group.

### QBA results for hospitalisations

SBA resulted in an OR = 1.75. Summary-level PBA shifted ORs away from the null (OR 2.20 (95% SI 1.45 – 7.38)). Record-level PBA with subsequent logistic regression gave a median adjusted OR of 1.88 (95% SI 1.39 – 3.79), and an IPT-weighted OR 1.74 (95% SI 1.27 – 3.56). No iterations were discarded due to negative cell counts.

### QBA results for deaths

SBA for outcome misclassification of deaths gave an OR = 1.63. Summary-level PBA of deaths resulted in an OR of 1.83 (95% SI 1.35 – 3.28). Record-level PBA with subsequent logistic regression gave a median unweighted OR of 1.76 (95% SI 1.28 – 2.84) and IPT-weighted OR of 1.55 (95% SI 1.10 – 2.55). 0.13% of the iterations resulted in negative cell counts and were therefore discarded.

### Analysis excluding people using triple therapy

Excluding people using triple therapy from the ICS group gave ORs of 1.21 (95% CI 0.93 – 1.56) for COVID-19 hospitalisations and 1.29 (95% CI 0.92 – 1.81) for COVID-19 deaths. IPTW shifted the results towards the null (OR 1.19 (95% CI 0.92 – 1.53) for COVID-19 hospitalisations, OR 1.24 (95% CI 0.88 – 1.74) for COVID-19 deaths). Summary-level QBA for COVID-19 hospitalisations resulted in an OR 1.44 (95% SI 0.95 – 4.00). Record-level PBA with subsequent logistic regression resulted in an unweighted OR 1.88 (95% SI 0.82 – 2.17) and a IPT-weighted OR 1.26 (95% SI 0.78 – 2.10).

Summary level QBA for COVID-19 deaths resulted in an OR 1.33 (95% SI 0.84 – 2.26). Record-level PBA with subsequent logistic regression resulted in an unweighted OR 1.32 (95% SI 0.82 – 2.17) and a IPT-weighted OR 1.26 (95% SI 0.78 – 2.10).

### Other Sensitivity Analyses

Including people with asthma or other chronic respiratory diseases in the cohort resulted in more than doubling in size of the ICS group, whereas the LABA/LAMA comparator group only increased in size by 23%. The effect estimates were closer to the null (OR for COVID-19 hospitalisation 1.55 (95% CI 1.32-1.81), OR for COVID-19 death 1.40 (95% CI 1.13-1.73), IPT-weighted OR for COVID-19 hospitalisation 1.42 (95% CI 1.22-1.65), IPT-weighted OR for COVID-19 death 1.17 (95% CI 0.96-1.42)) (S6.1).

We repeated the analysis using an exposure period defined as 6 months after the last prescription issue date. The results of the logistic regression did not change substantially compared to the analysis using an exposure defined as 60 days after the calculated end of the exposure period (S6.2). Coding all deaths missing in ONS as COVID-19 resulted in effect estimates closer to the null (S6.3).

Simulation of differential sensitivities of the classification of COVID-19 deaths between the treatment groups showed that sensitivity among the ICS group would have had to have been considerably higher than in the LABA/LAMA group in order to shift the effect estimates in such a way that the SI crosses 1 (S6.4)

## 5. Discussion

We found that among people using ICS, the risk of being hospitalised or dying with a record of COVID-19 was higher than among those using LABA/LAMA. However, the effect estimates attenuated towards the null after IPTW and after excluding people using triple therapy at baseline. This is likely explained by the fact that the ICS group included people using triple therapy who have more severe COPD and may be generally sicker than people using LABA/LAMA. Accounting for outcome misclassification using QBA shifted the effect estimates away from the null and increased the uncertainty around the point estimates, but did not change the conclusions of the analyses.

### Comparison to other studies

Two studies published early in the pandemic used UK population-level EHR data and the ONS death registry to investigate the association between ICS and COVID-19 deaths and hospitalisations. The adjusted estimate in our study for COVID-19 death is almost identical to the effect estimate in the OpenSAFELY study (aHR for COVID-19 death 1.39 (1.10–1.76)), possibly due to similarities in study design[5]. A study using the QResearch database[8] found a more modest risk of severe COVID-19 and COVID-19 death associated with ICS use, independent of underlying respiratory disease (COVID-19 hospitalisation aHR 1.13 (1.03-1.23), COVID-19 death aHR 1.15 (1.01-1.31)). Both studies addressed confounding as a potential source of bias and discussed potential exposure misclassification. There were several other studies that investigated inhaled corticosteroids and COVID-19 outcomes, but in different populations, e.g., in patients with COVID-19 infection or patients hospitalised with COVID-19[6,7,9]. Despite concerns that COVID-19 outcomes may have been misclassified, we are not aware of any other pharmacoepidemiological studies which have attempted to quantitatively correct for such outcome misclassification of COVID-19 outcomes.

### Correcting for potential outcome misclassification

All implementations of QBA shifted the effect estimates away from the null, in line with the heuristic that non-differential misclassification biases results towards the null on average. This suggests that conventional estimates underestimate the association between ICS and severe COVID-19. However, what our assessment adds is confirmation that a realistic extent of misclassification would not have resulted in a shift away from the null substantial enough to have warranted a change in conclusions of the study. Additionally, the uncertainty around the point estimates increased in PBA because the SI takes into account more sources of random error than the conventional CI[36]. As expected, the QBA methods which additionally allowed us to control for confounding gave ORs closer to the null. In sensitivity analyses looking at differential misclassification, we found that differential misclassification would have had to have been substantial, with sensitivity of >90% in the ICS arm and <60% in the LABA/LAMA arm, to have concealed a protective effect. We also found that even relatively small decreases in specificity resulted in substantial shifts in the effect estimate, while changes in sensitivity were less consequential.

One challenge we encountered was negative cell values, with the initially specified distributions for hospitalisations resulting in a large proportion of simulations with negative cell values, driven by false-positives (i.e., low specificity) in the LABA/LAMA group. This indicated that the chosen distributions, and consequently sampled combinations of sensitivity and specificity, were incompatible with the data, and has been known to happen when outcomes are very rare[36]. As a result, we increased the lower bound of the specificity distribution to 0.97 to obtain plausible cell counts.

There are many different tutorials and guides to the different forms of QBA used here[36,37], and each method has strengths and limitations. SBA, while easily implemented using available tools[38], is limited in its value as it requires the bias parameters to be known with relative certainty and does not give a measure of uncertainty of the results. Summary-level PBA is more time-intensive to implement but offers the advantage of generating a SI around the point estimate. Both approaches are limited in their analysis, however, as the calculation of the effect estimates is based on 2x2 tables rather than individual records. Record-level PBA is more sophisticated in this respect. As each row of data is corrected for misclassification, the analyst has the same amount of analytical flexibility, e.g., regarding methods to account for confounding, as in a conventional analysis without QBA. However, record-level PBA requires substantial computational power which may limit its application to large datasets. All methods of QBA performed similarly in this setting, but ultimately, the choice of QBA method depends on the degree of concern around severity and impact of outcome misclassification and the analytical and computational resources available.

### Strengths and limitations

Strengths of this study include the size of CPRD Aurum, which has been shown to be representative of the UK population. HES data captured the majority of COVID-19 hospitalisations in England[39,40], and ONS death data is the most reliable source of data on deaths in the UK. We used sensitivity analyses to assess different definitions of exposure, potential residual confounding by underlying disease and the impact of exclusion criteria.

Due to the short follow-up period, we could not use a new-user design, and assumed that any causal effect of ICS on the outcomes would be independent of treatment history and length. Therefore, the study population is potentially heterogeneous in terms of disease stage and treatment history. By excluding people using triple therapy in a sensitivity analysis, we made the treatment groups more comparable. This resulted in effect estimates closer to the null, and the CI indicated weak evidence of an increased risk in people using ICS. Even after excluding patients using triple therapy, patients in the ICS group may be different to those using LABA/LAMA, as ICS are recommended for patients with high eosinophil counts and frequent exacerbations.[41] The covariates did not include symptoms of COPD or measures of lung function, which could have additionally controlled for confounding by disease severity. The analysis excluding triple therapy users may have been underpowered to detect an increased risk in the ICS users, but suggests their inclusion in the study led to confounding, highlighting the trade-off of power and improved comparability of the treatment groups.

Although outcome misclassification was thought to be the most impactful source of misclassification in this study and was therefore the focus of the QBA, exposure misclassification may also have affected this study. Previous work has shown a spike in prescriptions in the UK in March 2020, meaning that estimated exposure durations may be less reliable than in non-pandemic times[42]. Additionally, this study did not capture in-hospital prescriptions as HES does not hold this information.

Our approach to QBA required several strong assumptions, which are largely untestable and likely imperfect. However, the alternative of not doing QBA assumes that outcome sensitivity and specificity were both 100%, which is an even stronger and less plausible position.

Particularly in the early months of the pandemic, treatments and the healthcare system’s response to COVID-19 were rapidly evolving, and it is therefore likely that sensitivity and specificity of recording of COVID-19 outcomes changed over time. The values and distributions chosen in this analysis represent best estimates. For excess deaths, the chosen values were based on an assumption that all excess deaths were true COVID-19 deaths. While this is a strong assumption, we made it based on data on deaths and excess deaths reported by the ONS and COVID-19 death counts reported to the UK government[31,32]. The values for excess deaths were based on available data from March to May 2020[31]. It is likely that sensitivity increased with time, but there were few COVID-19 deaths in the summer months of 2020 (June-August). For hospitalisations, we informed our estimates of sensitivity and specificity using validation studies conducted in North America, and the bounds given by the data. While coding practices between countries and healthcare systems may differ, studies from the UK were not available at the time of writing, so these values were taken as the best available estimates.

As methods of QBA are not yet well established for time-to-event outcomes, we assumed the recorded dates and the occurrence of hospitalisations and deaths to be accurate, and only attempted to correct for misclassified causes.

### Conclusions

In conclusion, we observed increased risks of COVID-19 hospitalisation and death among ICS-users compared to LABA/LAMA users. However, taken together with the results of the analysis excluding triple therapy users, the observed increase in risk may be attributed to residual confounding, even after IPTW. Quantitative bias analysis showed that outcome misclassification was unlikely to have substantially changed the results of the analysis, despite concerns around the classification of COVID-19 deaths early during the pandemic.

## Data Availability

No additional data available. Data management and analysis code and all code lists are available on our GitHub repository.

https://github.com/bokern/ics_covid

## Acknowledgments and funding

This study is based in part on data from the Clinical Practice Research Datalink obtained under licence from the UK Medicines and Healthcare products Regulatory Agency. The data is provided by patients and collected by the NHS as part of their care and support. The interpretation and conclusions contained in this study are those of the authors alone. The study was approved by the Independent Scientific Advisory Committee (approval number: 22_001876).

MPB is funded by a GSK PhD studentship to investigate the application of quantitative bias analysis in observational studies of COVID-19. IJD has unrestricted grants from and shares in GSK. AS is employed by LSHTM on a fellowship funded by GSK. JH was employed by GSK and owned stock in GSK during the conduct of this work. CTR and JQ report no conflicts of interest.

## Statements and Declarations

### Funding

MB is funded by a GSK PhD studentship to undertake this work.

### Author contributions

MB, AS, CTR, IJD contributed to the study design. MB conducted the data management and analysis, and drafted the manuscript. All authors were involved in design and conceptual development, and reviewed, edited and approved the final manuscript.

### Ethics Approval

The study was approved by the London School of Hygiene and Tropical Medicine Research Ethics Committee (Reference: 27896) and the Independent Scientific Advisory Committee of the UK Medicines and Healthcare Products Regulatory Agency (approval number: 22_001876).

## Notes

### Clinical Protocols

https://catalogues.ema.europa.eu/node/3194/administrative-details

https://www.cprd.com/approved-studies/investigating-biases-observational-studies-inhaled-corticosteroids-and-risk-covid

