## Supplement for "Using quantitative bias analysis to adjust for misclassification of COVID-19 outcomes: An applied example of inhaled corticosteroids and COVID-19 outcomes"

S 2.3. Plot of absolute standardised mean differences in the unweighted cohort, and after average treatment effect in the population (ATE) and average treatment effect in the treated (ATT) weighting 16

**S 3. Logistic regression to estimate the association between ICS and COVID-19 outcomes** 18

#### S 1. Study cohort

#### S 1.1. RECORD-PE checklist

| The RECORD statement for pharmacoepidemiology (RECORD-PE) checklist of items, extended from the STROBE and RECORD statements, which should be reported in non-interventional pharmacoepidemiological studies using routinely collected health data.[1] | | | | |
|---|---|---|---|---|
| Item No | STROBE items | RECORD items | RECORD-PE items | Page No / section |
| Title and abstract | | | | |
| 1 | (a) Indicate the study's design with a commonly used term in the title or the abstract.<br>(b) Provide in the abstract an informative and balanced summary of what was done and what was found. | 1.1: The type of data used should be specified in the title or abstract. When possible, the name of the databases used should be included.<br>1.2: If applicable, the geographical region and timeframe within which the study took place should be reported in the title or abstract.<br>1.3: If linkage between databases was conducted for the study, this should be clearly stated in the title or abstract. | --- | Abstract |
| **Introduction** | | | | |
| Background rationale | | | | |
| 2 | Explain the scientific background and rationale for the investigation being reported. | --- | --- | Introduction |
| Objectives | | | | |
| 3 | State specific objectives, including any prespecified hypotheses. | --- | --- | Introduction |
| **Methods** | | | | |
| Study design | | | | |
| 4 | Present key elements of study design early in the paper. | --- | 4.a: Include details of the specific study design (and its features) and report the use of multiple designs if used.<br>4.b: The use of a diagram(s) is recommended to illustrate key aspects of the study design(s), including exposure, washout, lag and observation periods, and covariate definitions as relevant. | Methods – Study design, S 1.2 |
| Setting | | | | |
| 5 | Describe the setting, locations, and relevant dates, including periods of recruitment, exposure, follow-up, and data collection. | --- | --- | Methods – Study design, S1.2 |
| Participants | | | | |
| | (a) Cohort study—give the eligibility criteria, and the sources and methods of selection of participants. Describe methods of follow-up. Case-control study—give the eligibility criteria, and | 6.1: The methods of study population selection (such as codes or algorithms used to identify participants) should be | --- | Methods – Study |

| | | | | |
|---|---|---|---|---|
| | the sources and methods of case ascertainment and control selection. Give the rationale for the choice of cases and controls.<br>Cross sectional study—give the eligibility criteria, and the sources and methods of selection of participants.<br>(b) Cohort study—for matched studies, give matching criteria and number of exposed and unexposed. Case-control study—for matched studies, give matching criteria and the number of controls per case. | listed in detail. If this is not possible, an explanation should be provided.<br>6.2: Any validation studies of the codes or algorithms used to select the population should be referenced. If validation was conducted for this study and not published elsewhere, detailed methods and results should be provided.<br>6.3: If the study involved linkage of databases, consider use of a flow diagram or other graphical display to demonstrate the data linkage process, including the number of individuals with linked data at each stage. | | population, S1.2, S1.4 |
| Variables | | | | |
| | Clearly define all outcomes, exposures, predictors, potential confounders, and effect modifiers. Give diagnostic criteria, if applicable. | 7.1: A complete list of codes and algorithms used to classify exposures, outcomes, confounders, and effect modifiers should be provided. If these cannot be reported, an explanation should be provided. | 7.1.a: Describe how the drug exposure definition was developed.<br>7.1.b: Specify the data sources from which drug exposure information for individuals was obtained.<br>7.1.c: Describe the time window(s) during which an individual is considered exposed to the drug(s). The rationale for selecting a particular time window should be provided. The extent of potential left truncation or left censoring should be specified.<br>7.1.d: Justify how events are attributed to current, prior, ever, or cumulative drug exposure.<br>7.1.e: When examining drug dose and risk attribution, describe how current, historical or time on therapy are considered.<br>7.1.f: Use of any comparator groups should be outlined and justified.<br>7.1.g: Outline the approach used to handle individuals with more than one relevant drug exposure during the study period. | Methods – Exposure, S1.3, Methods - outcome, covariates. Code available on GitHub |
| Data sources/measurement | | | | |
| | For each variable of interest, give sources of — data and details of methods of assessment (measurement). Describe comparability of assessment methods if there is more than one group. | --- | 8.a: Describe the healthcare system and mechanisms for generating the drug exposure records. Specify the care setting in which the drug(s) of interest was prescribed. | Methods – Data source, exposure |
| Bias | | | | |
| 9 | Describe any efforts to address potential sources of bias. | --- | --- | Methods - Statistical analyses, Quantitative bias analysis |
| Study size | | | | |

| | | | | |
|---|---|---|---|---|
| 10 | Explain how the study size was arrived at. | --- | --- | Cohort flowchart (S1.4) |
| Quantitative variables | | | | |
| 11 | Explain how quantitative variables were handled in the analyses. If applicable, describe which groupings were chosen, and why. | --- | --- | Methods - Covariates |
| Statistical methods | | | | |
| 12 | (a) Describe all statistical methods, including — those used to control for confounding.<br>(b) Describe any methods used to examine subgroups and interactions.<br>(c) Explain how missing data were addressed.<br>(d) Cohort study—if applicable, explain how loss to follow-up was addressed. Case-control study—if applicable, explain how matching of cases and controls was ad-dressed. Cross-sectional study—if applicable, describe analytical methods taking account of sampling strategy.<br>(e) Describe any sensitivity analyses. | --- | 12.1.a: Describe the methods used to evaluate whether the assumptions have been met.<br>12.1.b: Describe and justify the use of multiple designs, design features, or analytical approaches. | Methods- statistical analysis, S1.6, S3 |
| Data access and cleaning methods | | | | |
| 12 | --- | 12.1: Authors should describe the extent to which the investigators had access to the database population used to create the study population.<br>12.2: Authors should provide information on the data cleaning methods used in the study. | --- | Methods- Data source |
| Linkage | | | | |
| | --- | 12.3: State whether the study included person level, institutional level, or other data linkage across two or more databases. The methods of linkage and methods of linkage quality evaluation should be provided. | --- | Methods – Data source |
| Results | | | | |
| Participants | | | | |
| 13 | (a) Report the numbers of individuals at each stage of the study (eg, numbers potentially eligible, examined for eligibility, confirmed eligible, included in the study, completing follow-up, and analysed).<br>(b) Give reasons for non-participation at each stage.<br>(c) Consider use of a flow diagram | 13.1: Describe in detail the selection of the individuals included in the study (that is, study population selection) including filtering based on data quality, data availability, and linkage. The selection of included individuals can be described in the text or by means of the study flow diagram. | --- | Flow chart in S1., Methods- study population |
| Descriptive data | | | | |
| 14 | (a) Give characteristics of study participants (eg, demographic, clinical, social) and information on exposures and potential confounders.<br>(b) Indicate the number of participants with missing data for each variable of interest. | --- | --- | Table 2, S 1.6 |

| | | | | |
|---|---|---|---|---|
| | (c) Cohort study—summarise follow-up time (eg, average and total amount). | | | |
| Outcome data | | | | |
| 15 | Cohort study—report numbers of outcome events or summary measures over time. Case-control study—report numbers in each exposure category, or summary measures of exposure. Cross sectional study—report numbers of outcome events or summary measures. | --- | --- | Table 3 |
| Main results | | | | |
| 16 | (a) Give unadjusted estimates and, if applicable, confounder adjusted estimates and their precision (eg, 95% confidence intervals). Make clear which confounders were adjusted for and why they were included.<br>(b) Report category boundaries when continuous variables are categorised.<br>(c) If relevant, consider translating estimates of relative risk into absolute risk for a meaningful time period. | --- | --- | Methods – covariates, Figure 1 and 2 |
| Other analyses | | | | |
| 17 | Report other analyses done—eg, analyses of subgroups and interactions, and sensitivity analyses. | --- | --- | Methods – sensitivity analyses, Results – sensitivity analyses, S6 |
| Discussion | | | | |
| Key results | | | | |
| 18 | Summarise key results with reference to study objectives. | --- | --- | Discussion, paragraph 1 |
| Limitations | | | | |
| 19 | Discuss limitations of the study, taking into account sources of potential bias or imprecision. Discuss both direction and magnitude of any potential bias. | 19.1: Discuss the implications of using data that were not created or collected to answer the specific research question(s). Include discussion of misclassification bias, unmeasured confounding, missing data, and changing eligibility over time, as they pertain to the study being reported. | 19.1.a: Describe the degree to which the chosen database(s) adequately captures the drug exposure(s) of interest. | Discussion - Strengths and limitations |
| Interpretation | | | | |
| 20 | Give a cautious overall interpretation of — results considering objectives, limitations, multiplicity of analyses, results from similar studies, and other relevant evidence. | --- | 20.a: Discuss the potential for confounding by indication, contraindication or disease severity or selection bias (healthy adherer/sick stopper) as alternative explanations for the study findings when relevant. | Discussion |
| Generalisability | | | | |
| 21 | Discuss the generalisability (external validity) of the study results. | --- | --- | Strengths and limitations |
| Other information | | | | |

| | | | | |
|---|---|---|---|---|
| Funding | | | | |
| 22 | Give the source of funding and the role of the funders for the present study and, if applicable, for the original study on which the present article is based. | --- | --- | Funding, competing interests |
| Accessibility of protocol, raw data, and programming code | | | | |
| 22 | --- | 22.1: Authors should provide information on how to access any supplemental information such as the study protocol, raw data, or programming code. | --- | Methods, first sentence and last paragraph |
| RECORD=reporting of studies conducted using observational routinely collected data; RECORD-PE=RECORD for pharmacoepidemiological research; STROBE=strengthening the reporting of observational studies in epidemiology. This checklist has been duplicated from table 1 in *BMJ* 2018;363:k3532, as a standalone document for readers to print out or fill in electronically. | | | | |

#### S 1.2. Study diagram

Study diagram depicting inclusion and exclusion criteria, covariate assessment periods, exposure assessment, and follow-up times

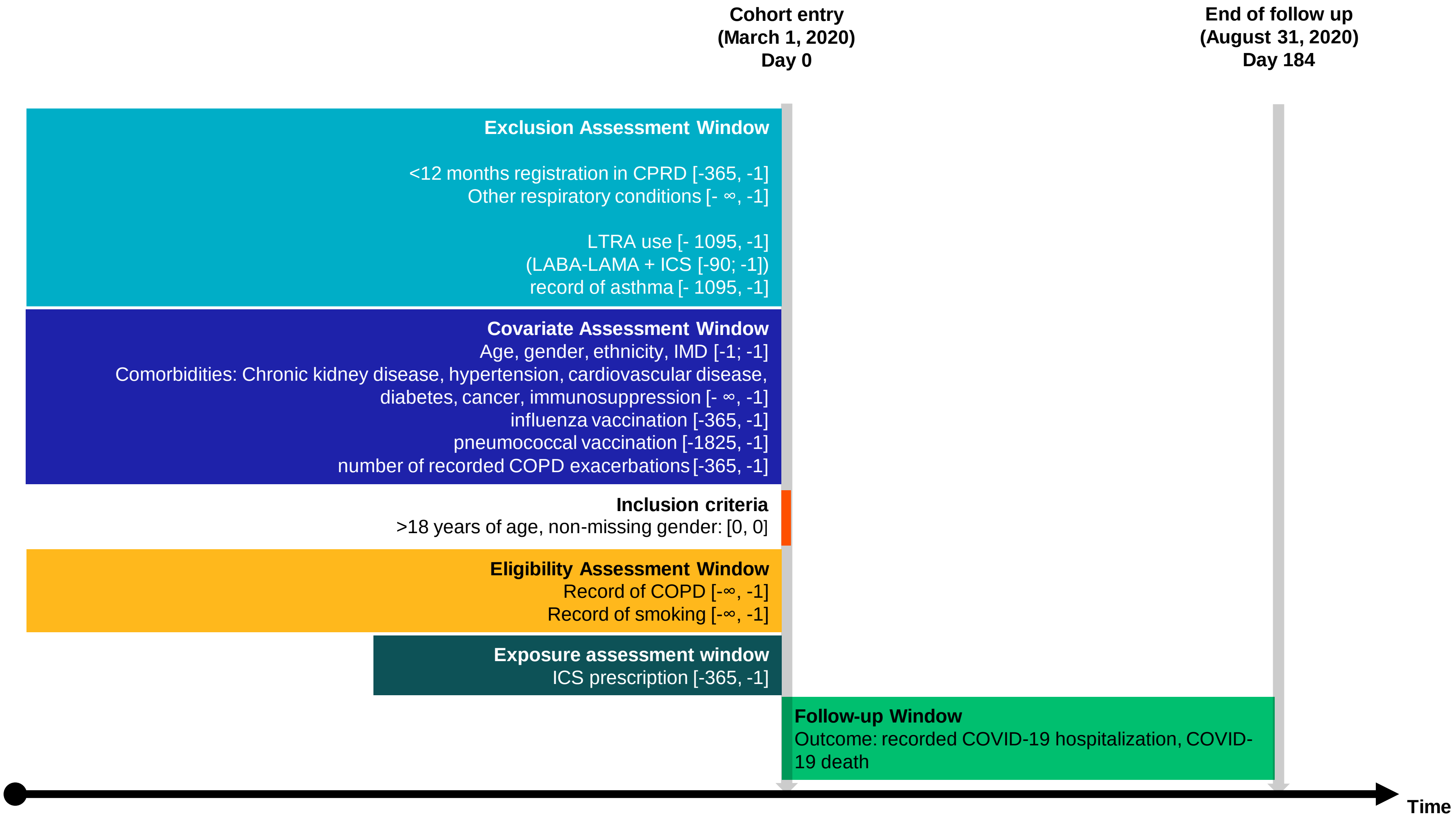

##### S 1.3. Treatment episodes

Information contained in the quantity variable was treated as the number of doses per prescription. The estimated number of days' drug supply was calculated by dividing the quantity by the number of daily doses. Where the GP-entered duration was plausible (>7 days and <100 days), that value was used as the exposure duration. Where this was considered implausible, the calculated $\frac{quantity}{daily\ dose}$ was used. Finally, if both were unavailable, the median entered prescription duration for that drug was used. Quantities <10 and >1000 were considered implausible and therefore as missing.

If a new prescription was issued before the end of the estimated exposure period of the preceding prescription, the overlapping days were "snowballed" and added on to the end of the calculated exposure period. However, the allowable overlap was capped at 90 days. When a prescription was issued within a 60-day grace period (two times the median duration) of the calculated exposure end of the preceding prescription of the same drug class, these prescriptions were considered as belonging to the same treatment episode and the patient was considered to be using their medication continuously. Discontinuations were thus defined as no new ICS prescription within 60 days of the calculated exposure end date. The discontinuation date was then the end date of the 60-day grace period.

#### S 1.4. Cohort selection flowchart

Flowchart depicting cohort selection criteria

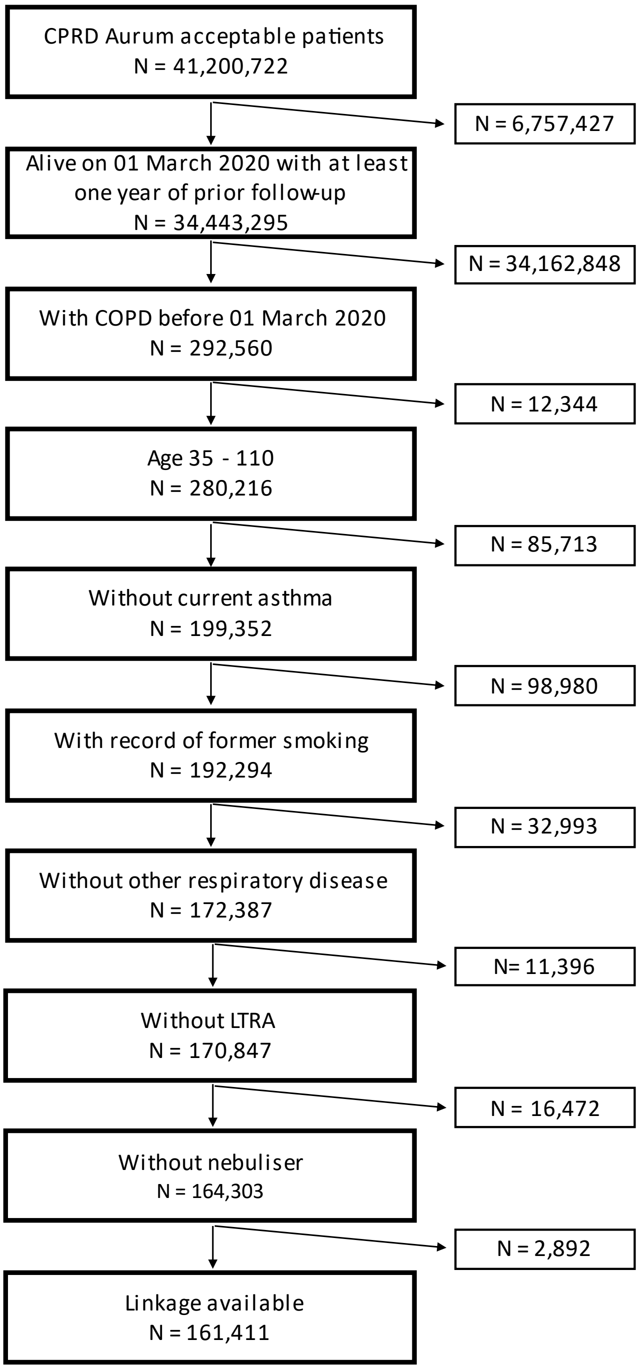

#### S 1.5. Baseline table including people who ever used either treatment during the study period

Baseline table including people who ever used either treatment during the study period (i.e., patients could be included in both groups)

| | **ICS/LABA N = 65205[1]** | **LABA/LAMA N = 29634[1]** |
|---|---|---|
| Age | | |
| *Mean (SD)* | 70.98 (10.64) | 70.44 (10.40) |
| *Median (25%-75%)* | 71.67 (63.67-78.67) | 70.67 (63.67-77.67) |
| Gender | | |
| *Male* | 34,701 (53%) | 16,199 (55%) |
| *Female* | 30,504 (47%) | 13,435 (45%) |
| BMI | | |
| *Underweight (<18.5)* | 3,619 (5.6%) | 1,339 (4.5%) |
| *Normal (18.5-24.9)* | 21,056 (32%) | 9,227 (31%) |
| *Overweight (25-29.9)* | 20,219 (31%) | 9,482 (32%) |
| *Obese (>=30)* | 20,311 (31%) | 9,586 (32%) |
| Index of Multiple Deprivation | | |
| *1* | 8,458 (13%) | 3,966 (13%) |
| *2* | 10,678 (16%) | 5,037 (17%) |
| *3* | 11,624 (18%) | 5,460 (18%) |
| *4* | 14,786 (23%) | 6,707 (23%) |
| *5* | 19,620 (30%) | 8,445 (28%) |
| *Missing* | 39 (<0.1%) | 19 (<0.1%) |
| Ethnicity | | |
| *White* | 57,315 (88%) | 25,935 (88%) |
| *South Asian* | 872 (1.3%) | 281 (0.9%) |
| *Black* | 422 (0.6%) | 193 (0.7%) |
| *Mixed* | 176 (0.3%) | 74 (0.2%) |
| *Unknown* | 6,420 (9.8%) | 3,151 (11%) |
| Smoking | | |
| *Current smoking* | 27,024 (41%) | 13,592 (46%) |
| *Former smoking* | 38,181 (59%) | 16,042 (54%) |
| Diabetes | 16,228 (25%) | 7,244 (24%) |
| Hypertension | 32,824 (50%) | 14,811 (50%) |
| Cardiovascular disease | 19,351 (30%) | 8,680 (29%) |
| Cancer | 12,160 (19%) | 5,758 (19%) |
| Past asthma | 17,811 (27%) | 3,794 (13%) |
| Kidney impairment | 19,269 (30%) | 8,798 (30%) |
| Any exacerbation in past 12 months | 25,999 (40%) | 8,196 (28%) |
| [1]n (%) | | |

#### S 1.6. Censoring

Proportion of patients censored for each analysis and median follow-up times and interquartile range (days)

| | ICS | LABA/LAMA | Total |
|---|---|---|---|
| % censored COVID-19 hospitalisation | 6.8 | 5.6 | 6.4 |
| % censored COVID-19 death | 6.3 | 5.3 | 6.1 |
| % censored all-cause death | 6.3 | 5.3 | 6.1 |
| COVID-19 hospitalisation | 183 (183-183) | 183 (183-183) | 183 (183-183) |
| COVID-19 death | 183 (183-183) | 183 (183-183) | 183 (183-183) |
| All-cause death | 183 (183-183) | 183 (183-183) | 183 (183-183) |

#### S 2. IPTW diagnostics

##### S 2.1. Unweighted propensity score distribution

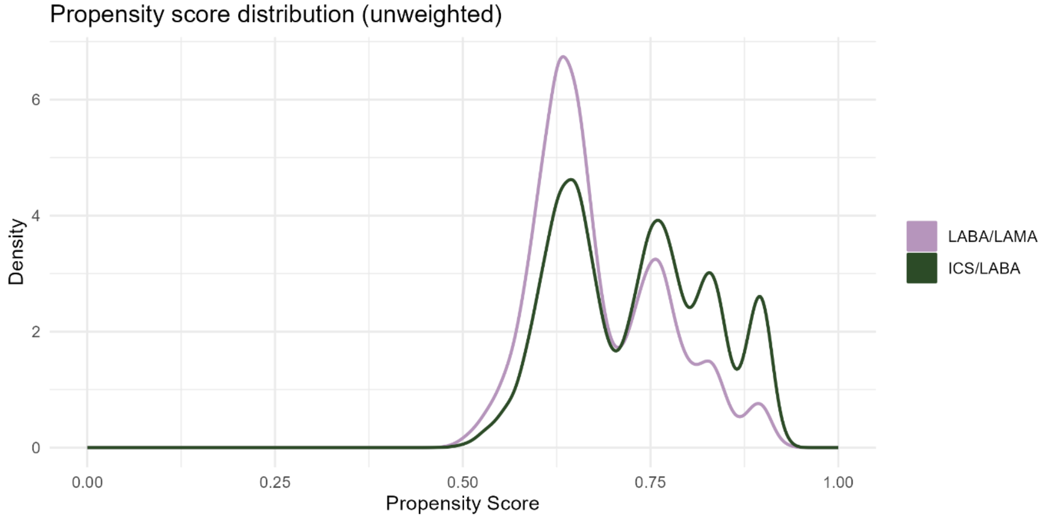

##### S 2.2. Standardised mean differences before and after weighting

| Variable | SMD (unweighted) | SMD (ATT unstabilised) | SMD (ATE unstabilised) | SMD (ATE stabilised) |
|---|---|---|---|---|
| **COPD exacerbation (past 12 months)** | 0.12798066 | -0.00143753 | -0.00100067 | -0.00100067 |
| **Pneumococcal Vaccine (past 5 years)** | -0.03821351 | -0.00058872 | -0.00037632 | -0.00037632 |
| **Influenza Vaccine (past 12 months)** | -0.00372733 | 0.00048466 | 0.00025228 | 0.00025228 |
| **Immunosuppression** | -0.00054689 | -8.1727E-06 | 1.3272E-06 | 1.3272E-06 |
| **Chronic Kidney Disease** | -0.00351788 | -0.00473349 | -0.00300453 | -0.00300453 |
| **Past asthma** | 0.15420739 | -0.00081798 | -0.00060229 | -0.00060229 |
| **Cancer** | -0.00973696 | -0.0005955 | -0.00043496 | -0.00043496 |
| **CVD** | 0.0061746 | -0.00158202 | -0.00091079 | -0.00091079 |
| **Hypertension** | 0.00310782 | -0.00276413 | -0.00172944 | -0.00172944 |
| **Diabetes** | 0.00397351 | -0.00218175 | -0.00146134 | -0.00146134 |
| **Former smoking** | 0.04518598 | -0.00114842 | -0.00091015 | -0.00091015 |
| **IMD 1** | -0.00846006 | 0.00138366 | 0.00093393 | 0.00093393 |
| **IMD 2** | -0.00649245 | -0.00122699 | -0.00078688 | -0.00078688 |
| **IMD 3** | -0.00479759 | -0.00128112 | -0.00083475 | -0.00083475 |
| **IMD 4** | 0.00235435 | -0.00065291 | -0.00050256 | -0.00050256 |
| **IMD 5** | 0.01739819 | 0.00168791 | 0.00113627 | 0.00113627 |
| **IMD: Missing** | -2.4366E-06 | 8.9448E-05 | 5.3987E-05 | 5.3987E-05 |
| **Ethnicity: White** | 0.00369526 | 1.3184E-05 | 3.1374E-05 | 3.1374E-05 |
| **Ethnicity: South Asian** | 0.00423305 | -0.00095781 | -0.00067803 | -0.00067803 |
| **Ethnicity: Black** | 0.00043748 | -0.00016263 | -9.7062E-05 | -9.7062E-05 |
| **Ethnicity: Mixed** | 0.00033796 | -0.00042465 | -0.00027614 | -0.00027614 |

| | | | | |
|---|---|---|---|---|
| **Ethnicity: Unknown** | -0.00870375 | 0.00153191 | 0.00101985 | 0.00101985 |
| **BMI: Normal (18.5-24.9)** | 0.01358054 | 0.00202498 | 0.00126309 | 0.00126309 |
| **BMI: Underweight (< 18.5)** | 0.01255973 | 0.00058224 | 0.00034402 | 0.00034402 |
| **BMI: Overweight (25-29.9)** | -0.01177742 | 0.00034835 | 9.4051E-05 | 9.4051E-05 |
| **BMI: Obese (>=30)** | -0.01436285 | -0.00295557 | -0.00170115 | -0.00170115 |
| **Gender: Female** | 0.01656316 | 0.00336453 | 0.00226494 | 0.00226494 |
| **Age at baseline** | 0.04803999 | -0.00219494 | -0.00169935 | -0.00169935 |

#### S 2.3. Plot of absolute standardised mean differences in the unweighted cohort, and after average treatment effect in the population (ATE) and average treatment effect in the treated (ATT) weighting

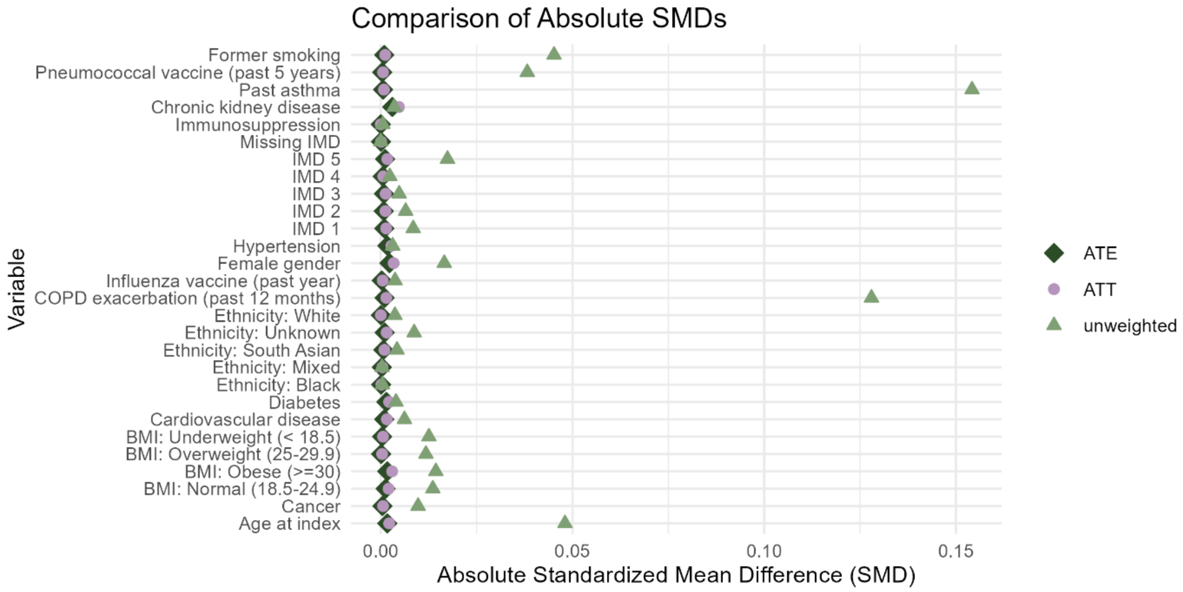

#### S 2.4. Kernel Density Plot of propensity score by treatment, after weighting (ATE)

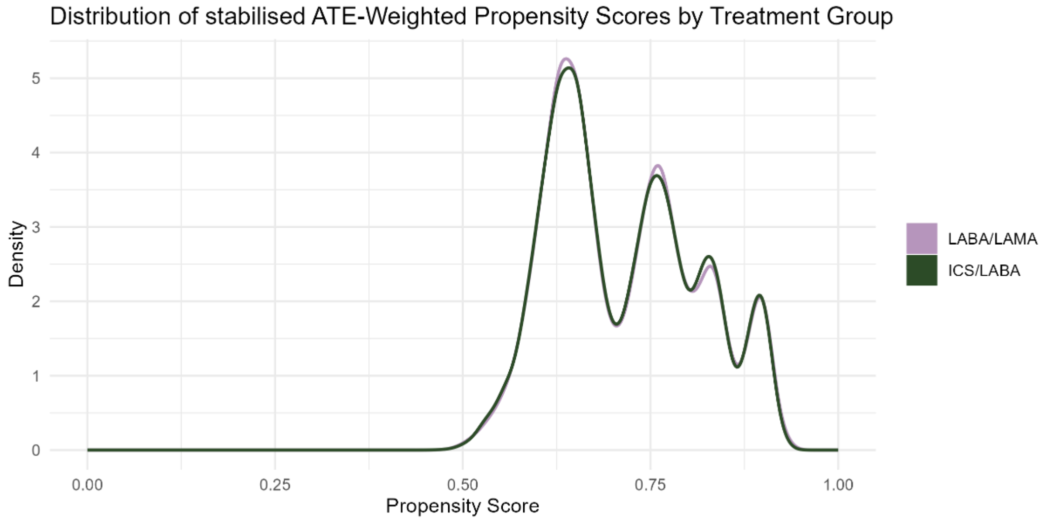

#### S 2.5. Kernel Density Plot of propensity score by treatment, after weighting (ATT)

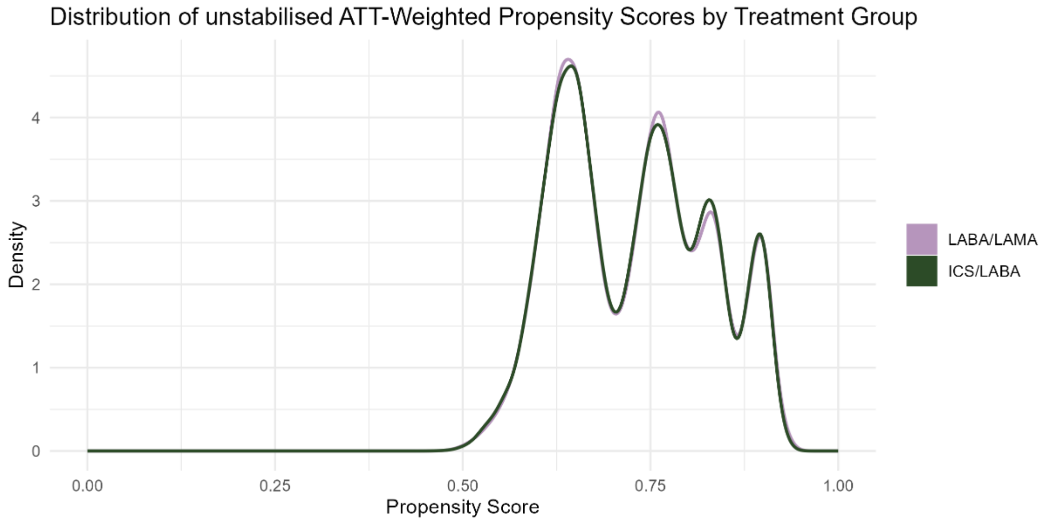

#### S 3. Logistic regression to estimate the association between ICS and COVID-19 outcomes

##### S 3.1. Log residuals

Log residuals for COVID-19 hospitalisation (unadjusted)

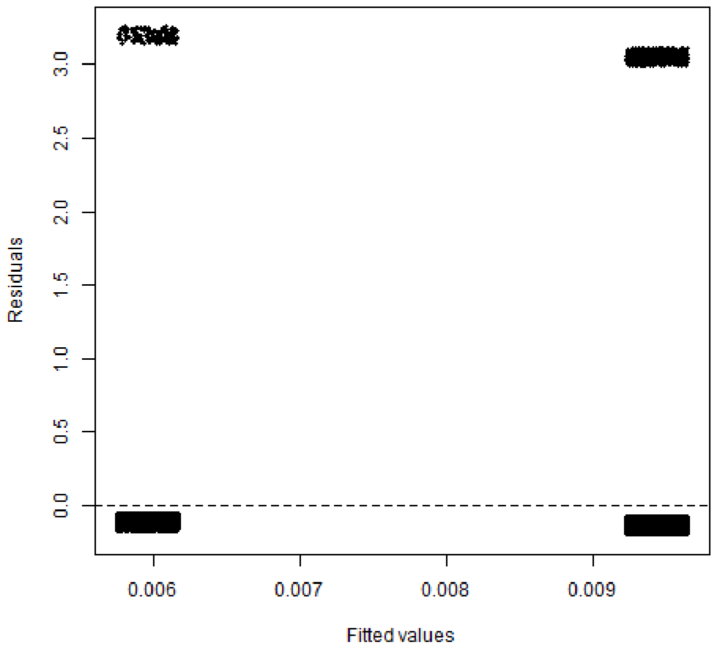

Log residuals for COVID-19 hospitalisation using stabilised ATE weights

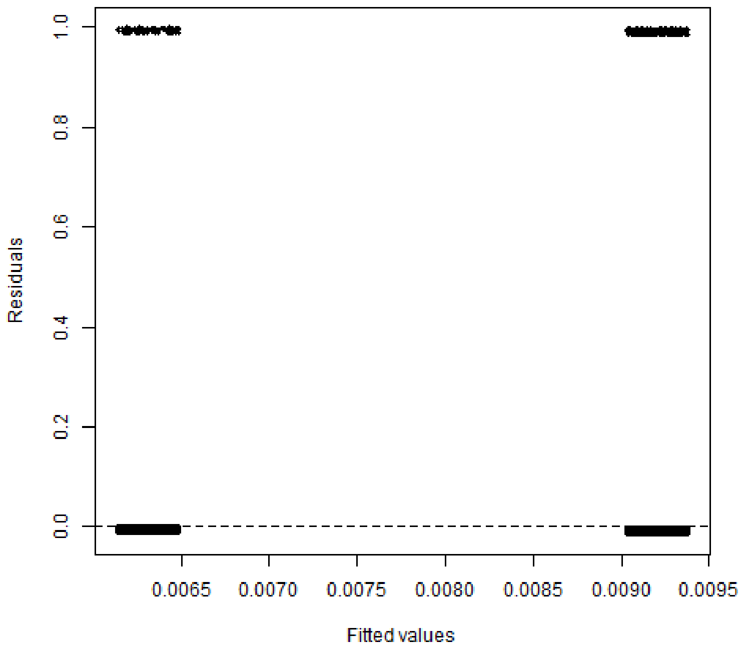

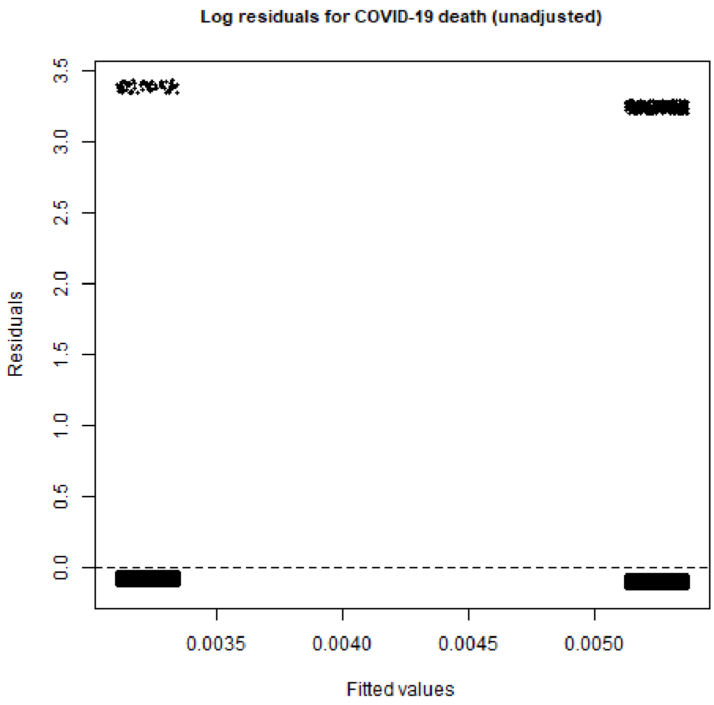
Log residuals for COVID-19 death (unadjusted)
Residuals
3.5
3.0
2.5
2.0
1.5
1.0
0.5
0.0
0.0035
0.0040
0.0045
0.0050
Fitted values

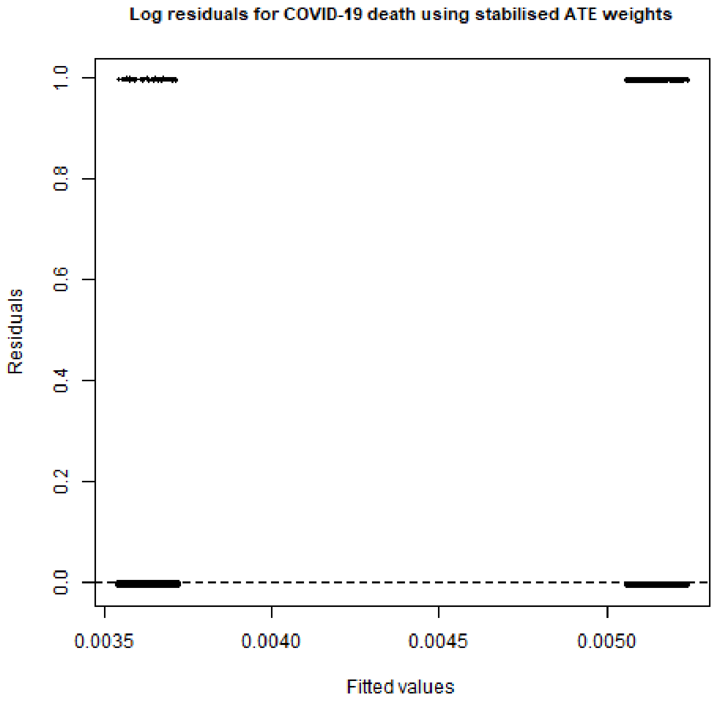
Log residuals for COVID-19 death using stabilised ATE weights
Residuals
1.0
0.8
0.6
0.4
0.2
0.0
0.0035
0.0040
0.0045
0.0050
Fitted values

**Log residuals for all-cause mortality (unadjusted)**

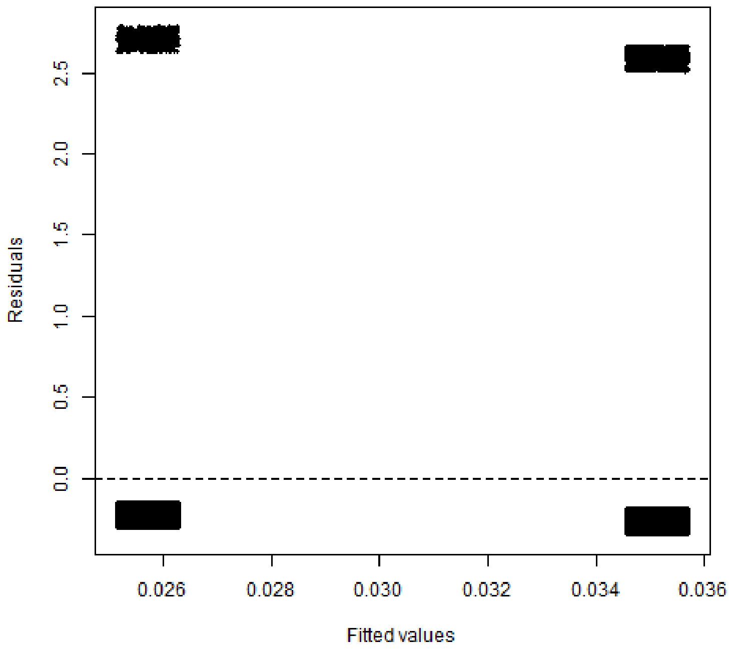

**Log residuals for COVID-19 death using stabilised ATE weights**

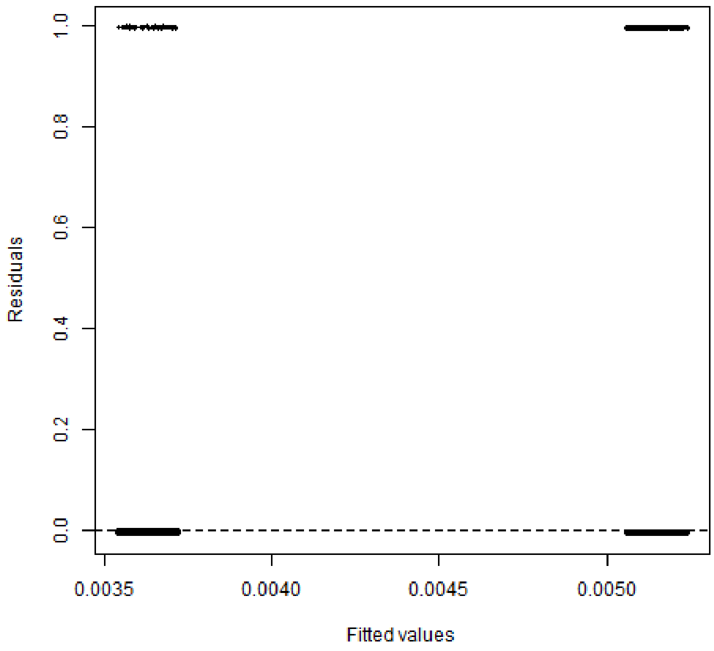

#### S 4. Quantitative bias analysis

##### S 4.1. Simple bias analysis

We conducted simple bias analysis using the following values:

Table S4.1.1 Sensitivity and specificity values used in simple bias analysis for the outcomes COVID-19 hospitalisation and deaths

| | Hospitalisation | Deaths |
|---|---|---|
| Sensitivity | 0.905 | 0.722 |
| Specificity | 0.953 | 0.966 |

Table S4.1.2 Equations to correct 2x2 table for outcome misclassification

| | | Exposure | |
|---|---|---|---|
| | | ICS | LABA/LAMA |
| Outcome | Outcome | $A = \frac{a - (a + c)(1 - Sp)}{Se - (1 - Sp)}$ | $B = \frac{b - (b + d)(1 - Sp)}{Se - (1 - Sp)}$ |
| | No outcome | $C = (a + c) - A$ | $D = (b + d) - B$ |
| | | a + c = A + C | b + d = B + D |

Table S4.1.3 Cell counts for COVID-19 hospitalisations by treatment group after adjustment for misclassification, among those with hospitalisation for any cause. Red indicates that the cell count was negative, indicating incompatibility of the bias parameters with the data.

| | Observed results | | | Adjusted results | | |
|---|---|---|---|---|---|---|
| | Exposure | | | Exposure | | |
| | ICS | LABA/LAMA | | ICS | LABA/LAMA | |
| COVID-19 hospitalisation | 529 | 133 | 662 | 81.4 | **-37.2** | 44.2 |
| Hospitalisation (other cause) | 9240 | 3376 | 12616 | 9687.6 | 3546.2 | 13233.8 |
| | 9769 | 3509 | 13279 | 9769 | 3509 | |

For deaths, quantitative bias analysis was used to correct for cause of death among people who died of any cause.

Table S4.1.4 Cell counts for the outcome COVID-19 death by treatment group after adjustment for misclassification, among those with hospitalisation for any cause.

| | Observed results | | | Adjusted results | | |
|---|---|---|---|---|---|---|
| | Exposure | | | Exposure | | |
| | ICS | LABA/LAMA | | ICS | LABA/LAMA | |
| COVID-19 death | 294 | 72 | 366 | 330.3 | 76.5 | 406.8 |
| Death (other cause) | 1675 | 502 | 2177 | 1491.7 | 450.5 | 1942.8 |
| Total deaths | 1969 | 574 | | 1822 | 527 | |

Subsequently, people who did not die during the study period were included to calculate effect estimates.

Table S4.1.5 Cell counts for COVID-19 death by treatment group among the entire cohort

| | Observed results | | | Adjusted results | | |
|---|---|---|---|---|---|---|
| | Exposure | | | Exposure | | |
| | ICS | LABA/LAMA | | ICS | LABA/LAMA | |
| COVID-19 death | 294 | 72 | 366 | 330.3 | 76.5 | 406.8 |
| No COVID-19 death | 55765 | 22247 | 78012 | 55728.7 | 22242.5 | 77971.2 |
| Total | 56059 | 22319 | | 56059 | 22319 | |

$$OR_{unadj} = \frac{294 * 22247}{72 * 55765} = 1.629$$

$$OR_{QBA} = \frac{330.3 * 22242.5}{76.5 * 55728.7} = 1.723$$

#### S 4.2. Bias parameters: sources and calculation

##### S 4.2.1. Summary of validation studies for hospitalisations

| **Study** | **Country** | **Time period** | **Data sources/setting** | **Hospitalisation due to COVID-19** |
|---|---|---|---|---|
| Wu et al.[2] | Canada | 1 March 2020 - 28 February 2021 | Compares the use of U07.1 in hospital data to the data held by Public Health Laboratory database, which captures SARS-CoV-2 laboratory PCR test results. U07.2 is not assessed. | ICD-10 code U07.1:<br>Sensitivity: 82.5% (81.8%–83.2%),<br>PPV = 93.1% (92.6%–93.6%).<br><br>combination of U07.1 and U07.3 (multisystem inflammatory syndrome associated with COVID-19):<br>sensitivity: 82.5% (81.9%–83.2%)<br>PPV: 92.9% (92.4%–93.4%) |
| Kadri et al.[3] | USA | 01 April - 31 May 2020 | Hospital diagnoses from Premier Healthcare Database. SARS-CoV-2 PCR test results from TheraDoc clinical surveillance system. A positive SARS-CoV-2 PCR test result during or up to 4 weeks prior to the hospitalization was used as the reference standard. | code U07.1:<br>Sensitivity: 98.01% (97.62%-98.39%),<br>Specificity: 99.04% (98.95% - 99.13%)<br>PPV: 91.52% (90.77%-92.27%)<br>NPV: 99.79% (99.75%-99.83%) |
| Kluberg et al.[4] | USA | 20 February –17 October 2020, Stratified by time period: February 20–March 31 (Time A), April 1–30 (Time B), May 1–October 17 (Time C) | Hospital claims data and laboratory test results from national laboratories that primarily process outpatient tests. This paper provides results stratified by time periods, and uses several different algorithms. | Overall PPV of code U07.1: 84.7% (84.0 – 85.4%) |
| Lynch et al.[5] | USA | April 1, 2020 - March 31 2021 | Records of ICD-10 code U07.1 from inpatient, outpatient, and | PPV of U07.1 in inpatient settings: 93.8% (91.8–95.6) |

| | | | | |
|---|---|---|---|---|
| | | | emergency care settings were extracted from VA medical record data. A weighted, random sample of 1500 records from each quarter of the study period was reviewed by study personnel to confirm active COVID-19 infection at the time of diagnosis and classify reasons for false positive records. | |
| Bodilsen et al.[6] | Denmark | 27 February – 4 May 2020 | Validates ICD-10 codes against medical records, looking at records of positive PCR test and clinical presentation. | PPV for COVID-19 was 99% (95% CI 99–100) compared with 99% (95% CI 98–100) when using definite cases only. |
| Bhatt et al.[7] | USA | April 1 - July 31 2020 | Identified inpatient encounters with ≥ 1 SARSCoV-2 RT-PCR in the Mass General Brigham health system. The agreement between COVID-19 positivity (RT-PCR) and primary or secondary ICD-10 coding of U07.1 was determined. This study splits data by month. | Sensitivity of U07.1 compared to positive PCR: 49.2% (47.1– 51.3) |
| Slater et al.[8] | UK | 23 March - 28 April 2020 | Data from Leeds Teaching Hospitals NHS Trust. | 162 patients with a positive SARS-CoV-2 PCR died. COVID-19 infection was recorded as the direct cause of death in 150 (93%) (sensitivity). Review of the records revealed 92% of patients had pulmonary infiltrates on chest radiography, and 97% required oxygen therapy → majority of hospitalised patients with positive SARS-CoV-2 PCR died as a direct consequence of COVID-19. |

S 4.2.2. Calculations of sensitivity and specificity for COVID-19 deaths

To inform the choice of values for the bias parameters, we used published information on excess deaths in England and Wales. There were 130,009 total deaths registered in England and Wales between 07 March 2020 and 01 May 2020. Of these, 46,380 were deemed to be excess deaths and 12,900 of these did not have COVID-19 recorded anywhere on the death certificate.[9] In the same time period, 36,323 COVID-19 deaths were reported to the UK government.[10] These include people who died from COVID-19, as decided by the clinician registering the death, and they were considered to be the total number of observed COVID-19 deaths. Based on the total number of excess deaths (46,380) and the number of excess deaths that did not have COVID-19 recorded as a cause of death on the death certificate (12,900), we calculated the number of deaths that were deemed to be true COVID-19 deaths that were also observed as COVID-19 death ($46{,}380 - 12{,}900 = 33{,}480$). We calculated the number of true non-COVID-19 deaths that were observed as COVID-19 (false positives): $36{,}323 - 33{,}480 = 2{,}843$.

Table S4.2.2 Data to inform the choice of bias parameters for misclassification of deaths, assuming that all observed excess deaths were true COVID-19 deaths.

| | | True | | |
|---|---|---|---|---|
| | | Non-COVID-19 death | COVID-19 death | |
| **Observed** | Non-COVID-19 death | 80,786 | **12,900** | 93,686 |
| | COVID-19 death | 2,843 | 33,480 | **36,323** |
| | | 83,629 | **46,380** | **130,009** |

*Numbers in **bold** were drawn from the literature. All other numbers were calculated from those.

Based on these numbers, estimates for sensitivity and specificity were calculated.

$$Se = \frac{33{,}480}{46{,}380} = 72.19\ \%$$

$$Sp = \frac{80{,}786}{83{,}629} = 96.60\ \%$$

#### S 4.3. Bias parameter sampling distributions

Sampled sensitivity for summary-level probabilistic bias analysis of outcome misclassification for COVID-19 hospitalisations

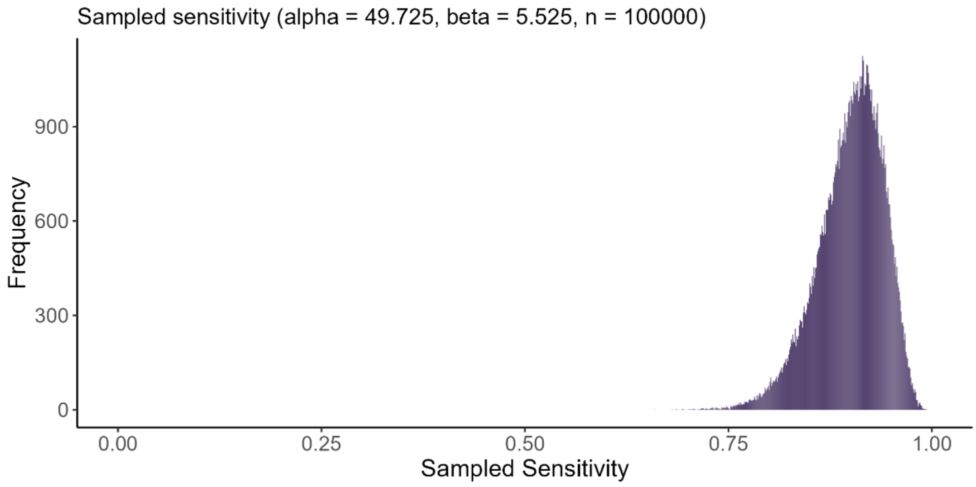

Sampled specificity for summary-level probabilistic bias analysis of outcome misclassification for COVID-19 hospitalisations

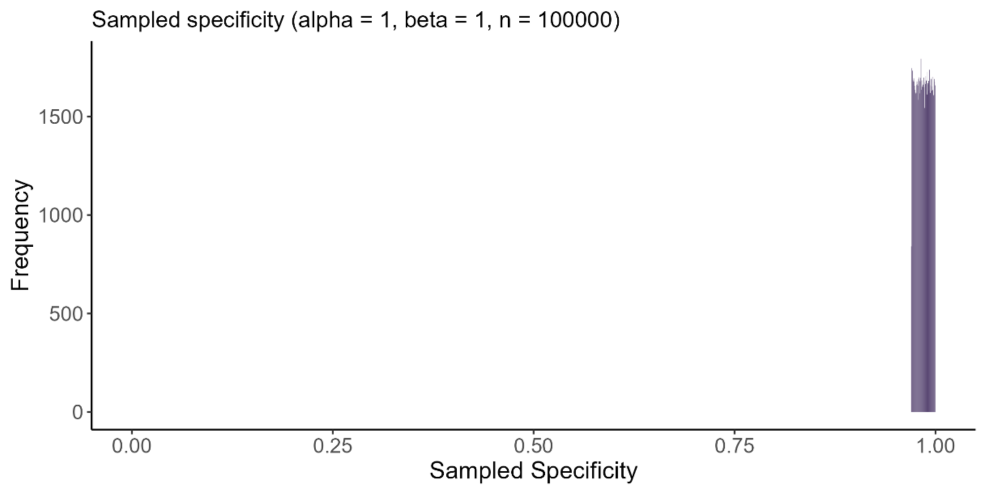

Sampled sensitivity for record-level probabilistic bias analysis of outcome misclassification for COVID-19 hospitalisations

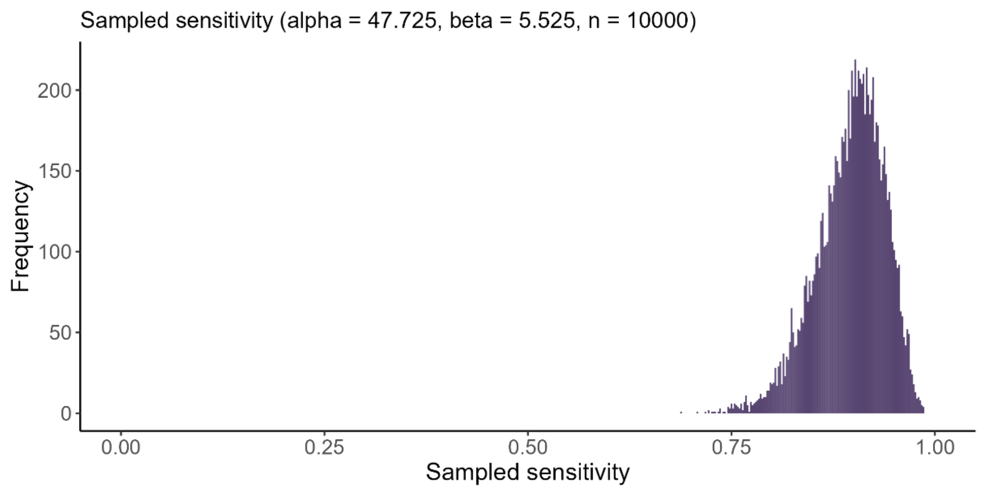

Sampled specificity for record-level probabilistic bias analysis of outcome misclassification for COVID-19 hospitalisations

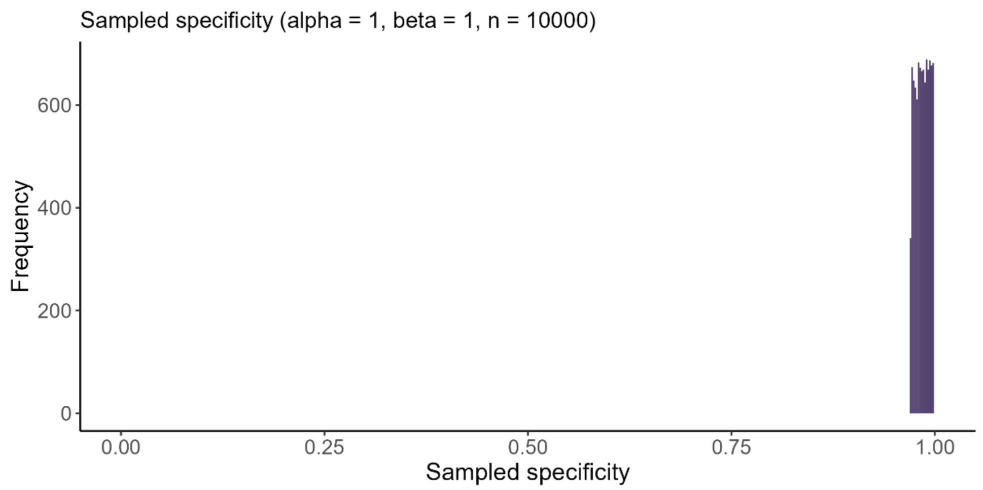

Sampled sensitivity for summary-level probabilistic bias analysis of outcome misclassification for COVID-19 deaths

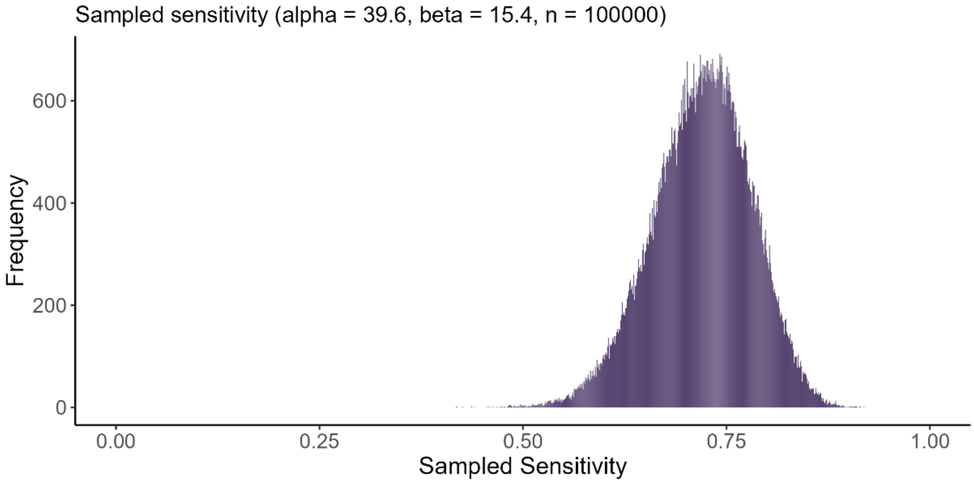

Sampled specificity for summary-level probabilistic bias analysis of outcome misclassification for COVID-19 deaths

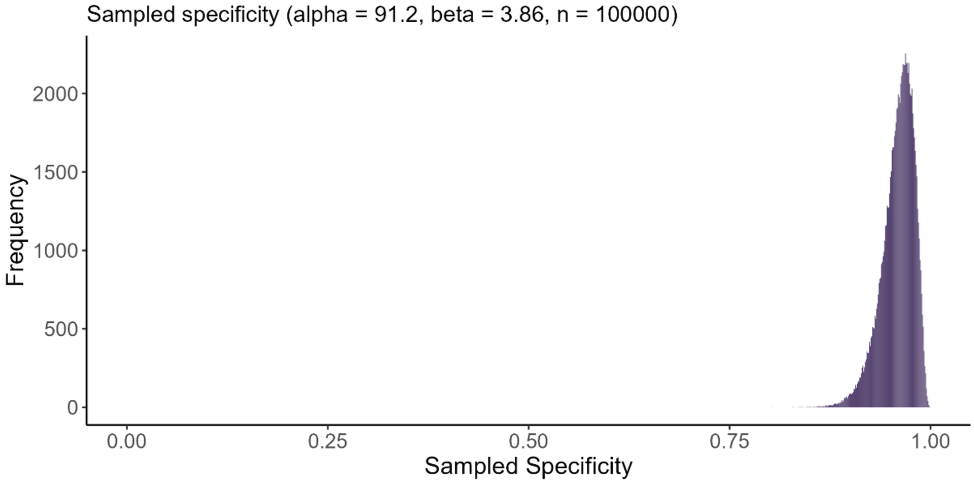

Sampled sensitivity for record-level probabilistic bias analysis of outcome misclassification for COVID-19 deaths

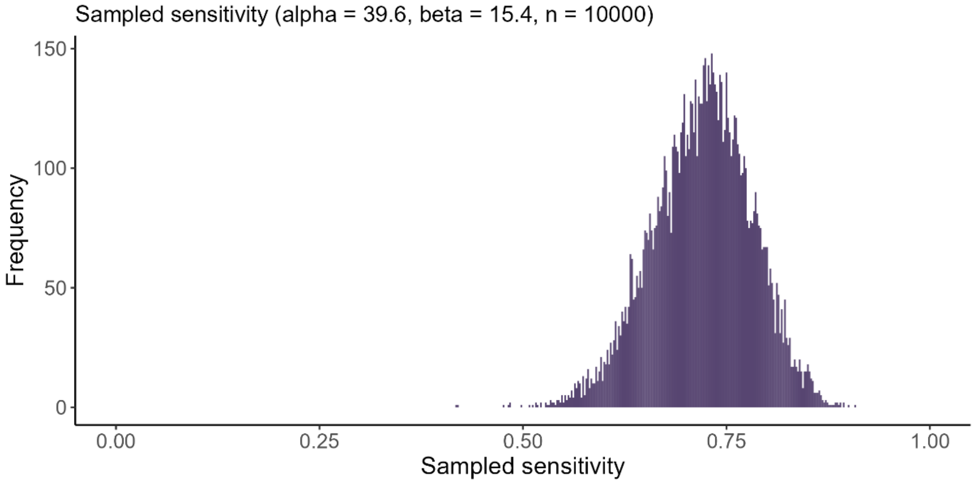

Sampled specificity for record-level probabilistic bias analysis of outcome misclassification for COVID-19 deaths

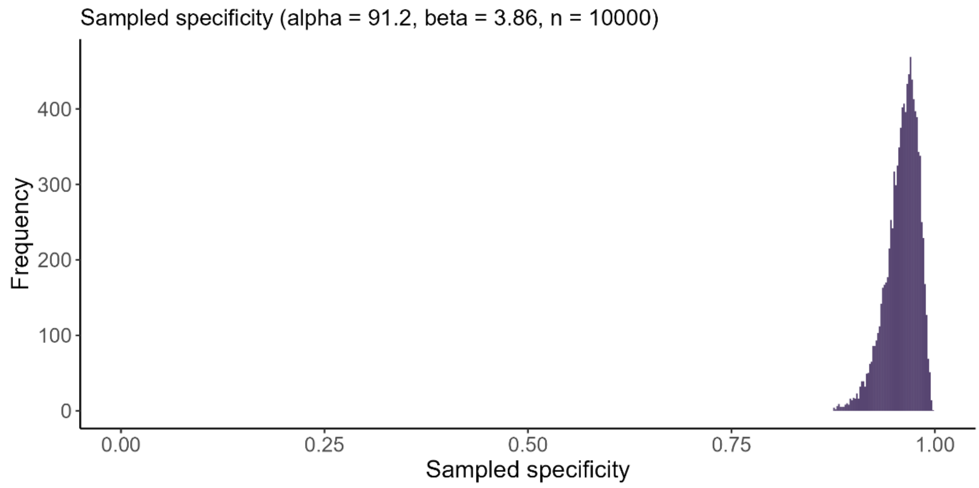

#### S 4.4. Method of summary-level bias analysis

##### 1) Choose bias parameters and specify their distributions

For misclassification, bias parameters are either sensitivity and specificity or positive and negative predictive values.

Distributions are assigned to bias parameters to reflect the fact that we do not know the value of the bias parameters with certainty.

##### 2) Draw values from the prespecified distributions using Monte-Carlo sampling.

##### 3) Apply the drawn values to the 2x2 table

Table S4.4.1 Correction of observed values for sampled sensitivity and specificity

| | Observed | | Bias-adjusted | |
|---|---|---|---|---|
| | ICS | Control | ICS | Control |
| (+) COVID-19 | $a$ | $b$ | $A = \frac{a - (a+c)*(1-SP_{E1})}{SE_{E1} - (1-SP_{E1})}$ | $B = \frac{b - (b+d)*(1-SP_{E0})}{SE_{E0} - (1-SP_{E0})}$ |
| (-) COVID-19 | $c$ | $d$ | $C = (a+c) - A$ | $D = (b+d) - B$ |
| | $a+c$ | $b+d$ | | |

The sampled values are used to correct the 2x2 table using the equations in Table (adapted from Fox et al.[11]).

##### 4) Estimate the prevalence of the outcome in each exposure group.

$$PrevD_{exp} = \frac{A}{A+C}$$

$$PrevD_{unexp} = \frac{B}{B + D}$$

The prevalence of the outcome necessarily needs to be calculated separately for each exposure group, as not doing so would imply that the null hypothesis of no difference between the treatment groups holds.[11]

In practice, instead of calculating the outcome prevalences directly, we sample from a beta distribution to reflect the random sampling error. As the mean of a beta distribution is $E(X) = \frac{\alpha}{\alpha+\beta}$, we can use the values A and C (or B and D, respectively) as alpha and beta parameters.

$$PrevD_{exp} \sim beta(A, C)$$
$$PrevD_{unexp} \sim beta(B, D)$$

5) **Use the sampled prevalence and the values for sensitivity and specificity to estimate predictive values.**

$$PPV_{exp} = \frac{Se * PrevD_{exp}}{(Se * PrevD_{exp}) + (1 - Sp) * (1 - PrevD_{exp})}$$

$$NPV_{exp} = \frac{Sp * (1 - PrevD_{exp})}{(1 - Se) * PrevD_{exp} + Sp * (1 - PrevD_{exp})}$$

$$PPV_{unexp} = \frac{Se * PrevD_{unexp}}{(Se * PrevD_{unexp}) + (1 - Sp) * (1 - PrevD_{unexp})}$$

$$NPV_{unexp} = \frac{Sp * (1 - PrevD_{unexp})}{(1 - Se) * PrevD_{unexp} + Sp * (1 - PrevD_{unexp})}$$

6) **Apply PPV and NPV values to 2x2 table**

Binomial trials are simulated to correct for the 2x2 using the predictive values

Table S4.1.2 Correcting the 2x2 using binomial trials and the predictive values

| | Observed | | Bias-adjusted | |
|---|---|---|---|---|
| | ICS | Control | ICS | Control |
| (+) COVID-19 | a | b | $A^* = rbinom(A, PPV_{exp}) + rbinom(C, 1 - NPV_{exp})$ | $B^* = rbinom(B1, PPV_{unexp}) + rbinom(D, 1 - NPV_{unexp})$ |
| (-) COVID-19 | c | d | $C^* = n_{exp} - A^*$ | $D^* = n_{unexp} - B^*$ |
| | a + c | b + c | | |

* A, B, C and D are rounded to integer values in order to conduct binomial trials.

By conducting binomial trials, we incorporate random error arising during the misclassification process.

7) **Calculate OR or RR from corrected table**

$$\text{OR} = \frac{A^* * D^*}{B^* * C^*}$$

$$\text{RR} = \frac{\frac{A^*}{A^* + C^*}}{\frac{B^*}{B^* + D^*}} = \frac{A^* * (B^* + D^*)}{B^* * (A^* + C^*)}$$

The random error is incorporated by multiplying a draw from the standard normal distribution with the standard error.

$$estimate_{total} = estimate_{adj} - z_i * SE_i^{adj}$$

8) **Repeat steps 2 – 6 a prespecified number of times (e.g. 100,000) to generate a distribution of effect estimates**
9) **Calculate median effect estimate and 95% simulation interval from the distribution of effect estimates**

#### S 4.5. Method of record-level bias analysis

Record-level probabilistic bias analysis was conducted similarly to summary-level bias analysis as described in Section **Error! Reference source not found.**. However, instead of conducting Bernoulli t rials on the summary-level 2x2 table in step 6, Bernoulli trials are conducted for each individual patient in order to reclassify them. By doing this, regression models can be fitted to each simulated dataset, and can take into account confounder adjustment or inverse probability of treatment weighting (IPTW).

For each individual row of data, the probability of having the outcome is calculated using the following equation:

$$\begin{aligned} p_i = {} & E_i * d_i * PPV_{E1} \\ & + E_i * (1 - d_i) * (1 - NPV_{E1}) \\ & + (1 - E_i) * d_i * PPV_{E0} \\ & + (1 - E_i) * (1 - d_i) * (1 - NPV_{E0}) \end{aligned}$$

where E = 1 is the exposed group, E = 0 is the unexposed group, d = 1 are people with the observed outcome, and d = 0 are people without the outcome of interest observed.

Based on the probability of having the outcome p, Bernoulli trials are conducted for each patient to adjust the outcome classification and create a new, simulated, binary outcome variable d:

$$d_i \sim rbinom(1, 1, p_i)$$

Regression models can be fit using the new, reclassified outcome variable d. Steps 7 and 8 are the same as for summary-level probabilistic bias analysis, as described in S 4.4.

#### S 4.6. Initial probabilistic bias analysis for hospitalisations with high proportion of negative cell counts (summary-level)

Sampled sensitivity for summary-level probabilistic bias analysis of outcome misclassification for COVID-19 hospitalisations (initially specified distribution)

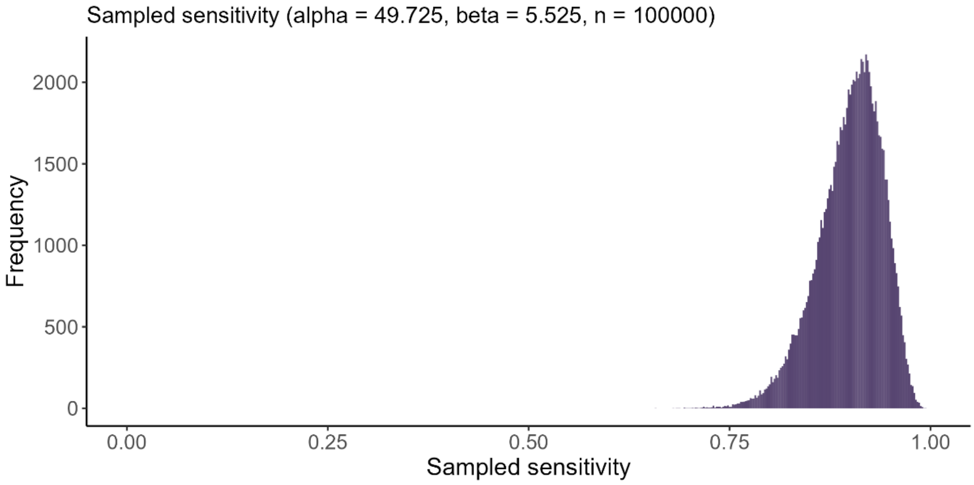

Sampled specificity for summary-level probabilistic bias analysis of outcome misclassification for COVID-19 hospitalisations (initially specified distribution)

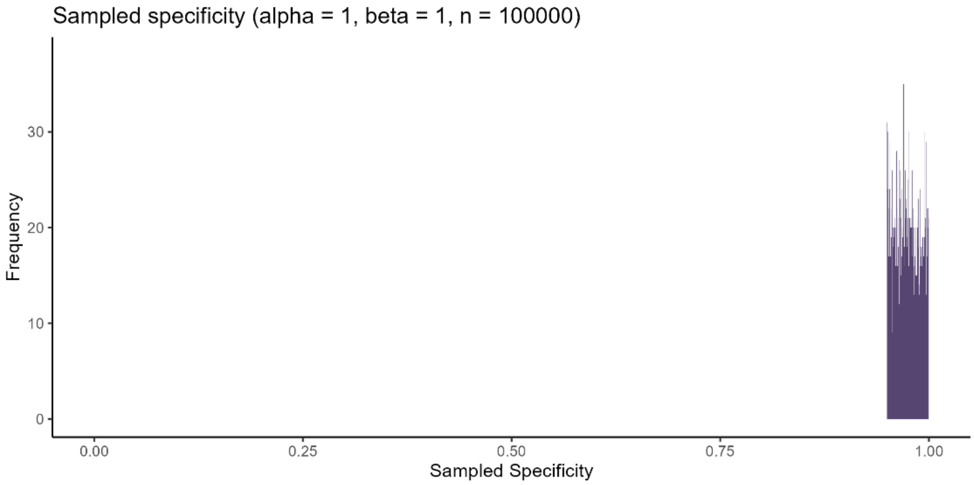

Sampled sensitivity values for summary-level probabilistic bias analysis of outcome misclassification for COVID-19 hospitalisations that result in plausible cell counts (initially specified distribution)

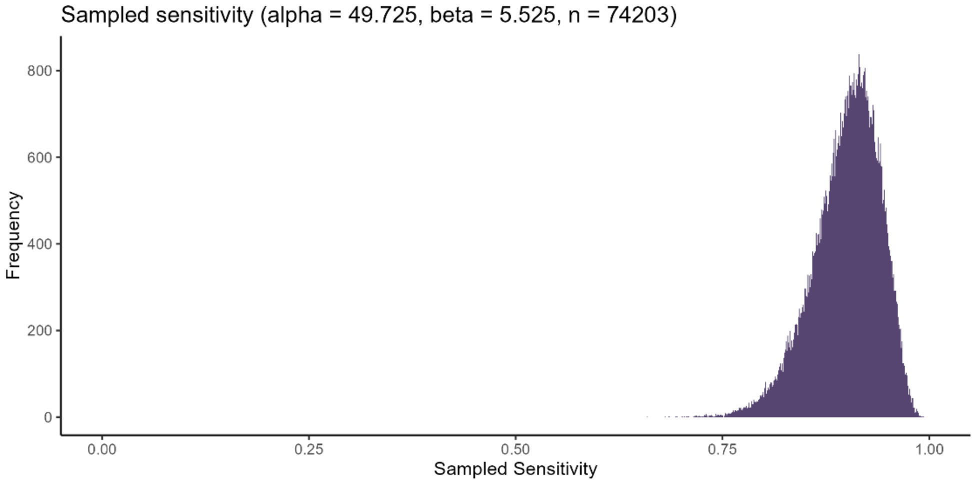

Sampled specificity values for summary-level probabilistic bias analysis of outcome misclassification for COVID-19 hospitalisations that result in plausible cell counts (initially specified distribution)

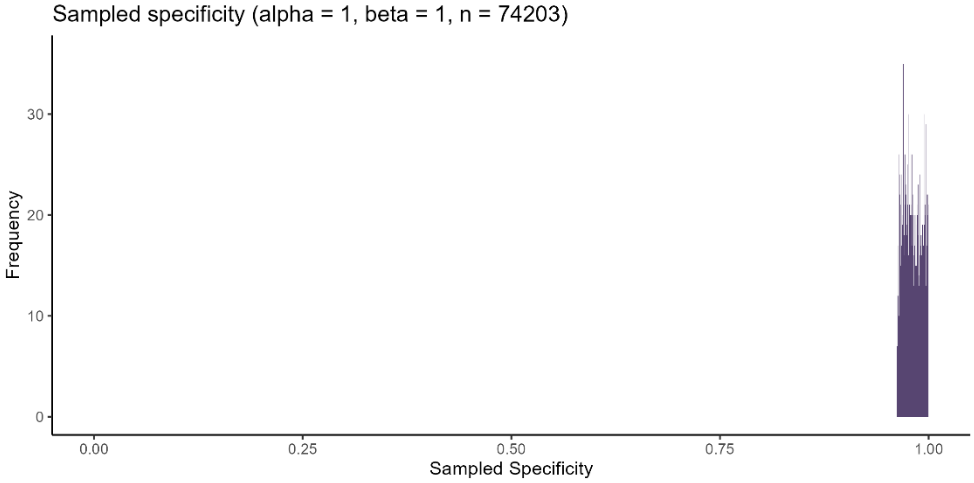

Sampled outcome prevalence by treatment group for summary-level probabilistic bias analysis for COVID-19 hospitalisations (initially specified distribution)

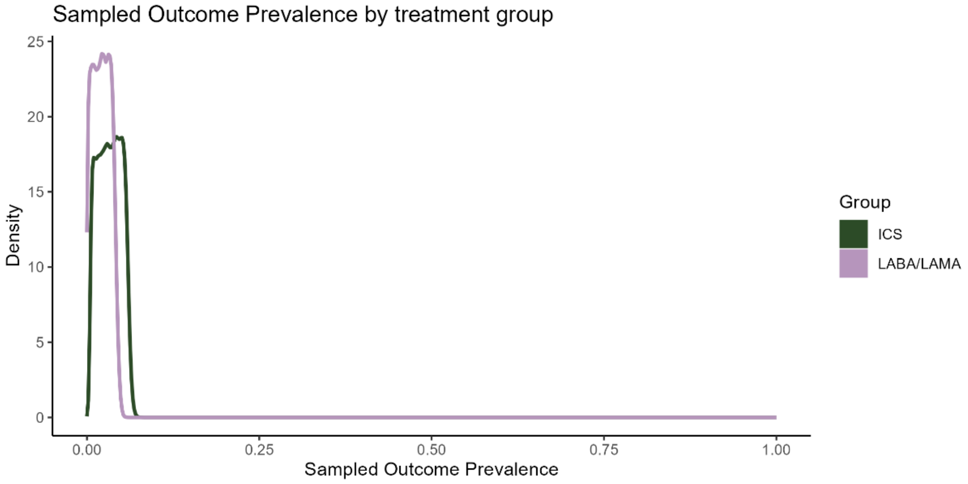

Sampled positive predictive values by treatment group for summary-level probabilistic bias analysis for COVID-19 hospitalisations (initially specified distribution)

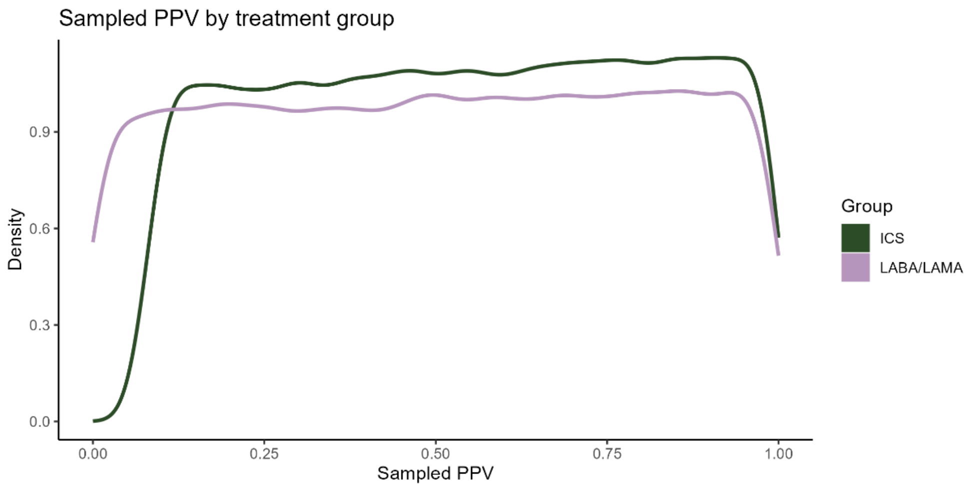

Sampled negative predictive values by treatment group for summary-level probabilistic bias analysis for COVID-19 hospitalisations (initially specified distribution)

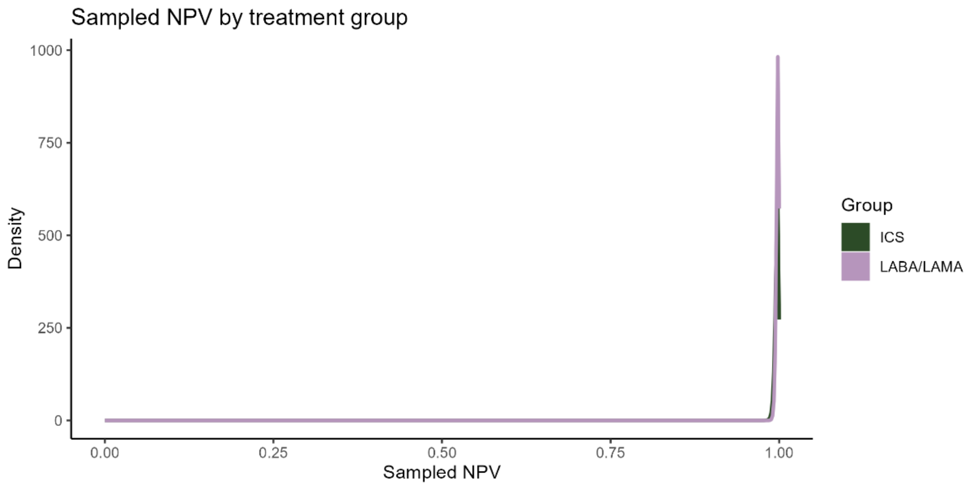

Distribution of risk ratios after adjusting for outcome misclassification for summary-level probabilistic bias analysis for COVID-19 hospitalisations (initially specified distribution)

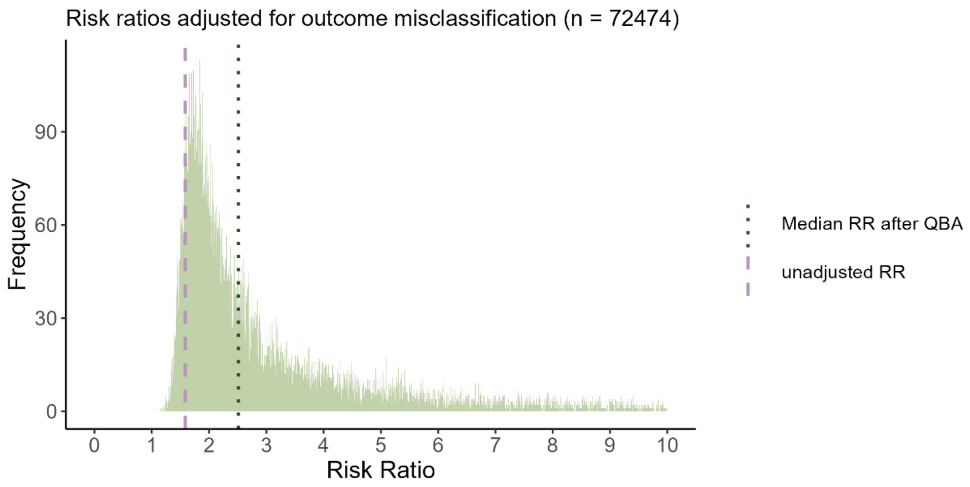

Distribution of odds ratios after adjusting for outcome misclassification for summary-level probabilistic bias analysis for COVID-19 hospitalisations (initially specified distribution)

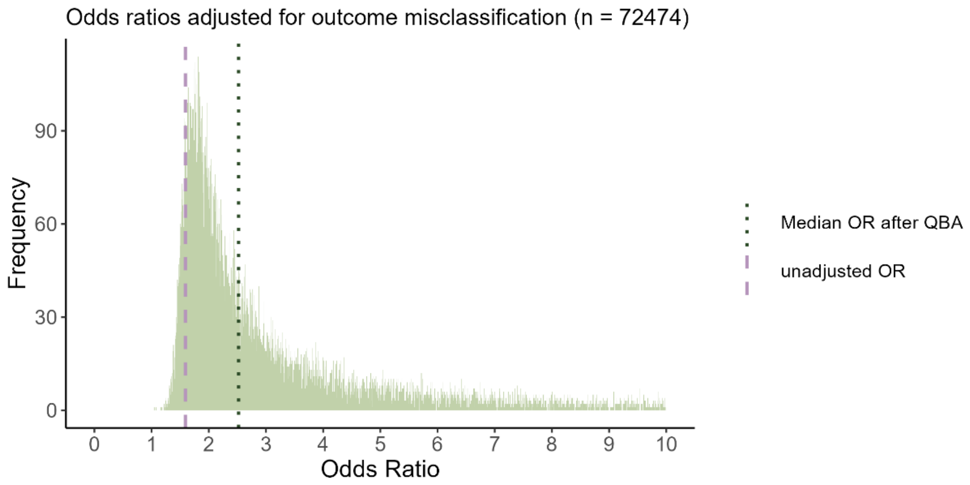

Scatter plot depicting which combination of sensitivity and specificity values result in possible and impossible cell counts.

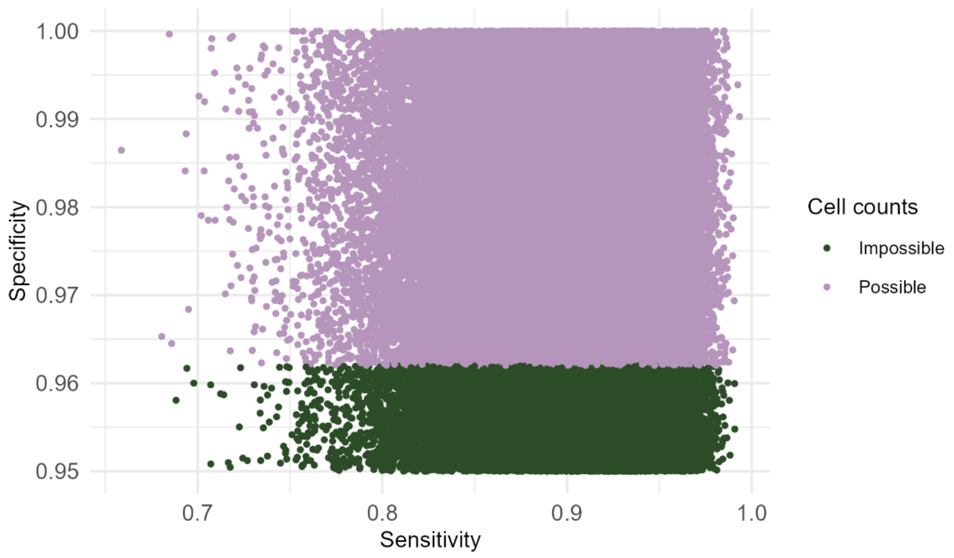

1.00
0.99
0.98
0.97
0.96
0.95
Specificity
0.7
0.8
0.9
1.0
Sensitivity
Cell counts
Impossible
Possible

#### S 4.7. Summary-level probabilistic bias analysis for COVID-19 hospitalisations (main analysis)

Sampled outcome prevalence by treatment group for summary-level probabilistic bias analysis for COVID-19 hospitalisations

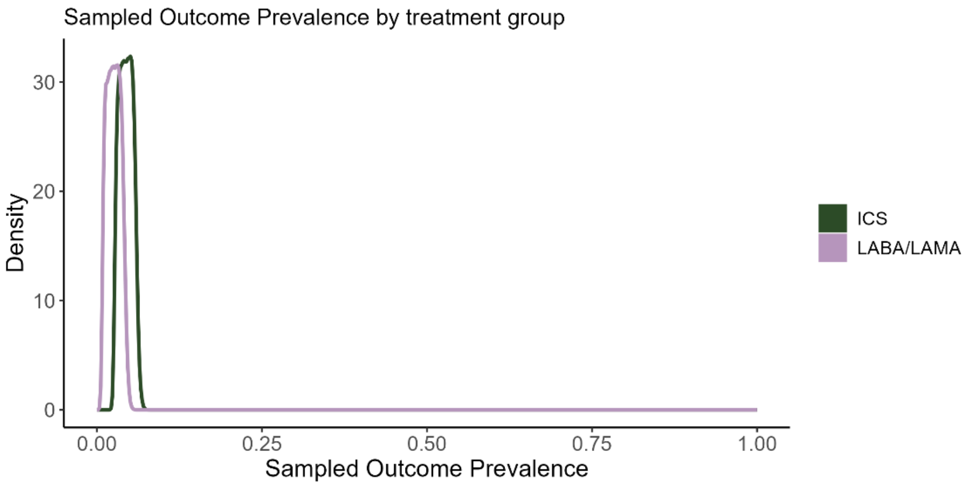

Sampled positive predictive values by treatment group for summary-level probabilistic bias analysis for COVID-19 hospitalisations

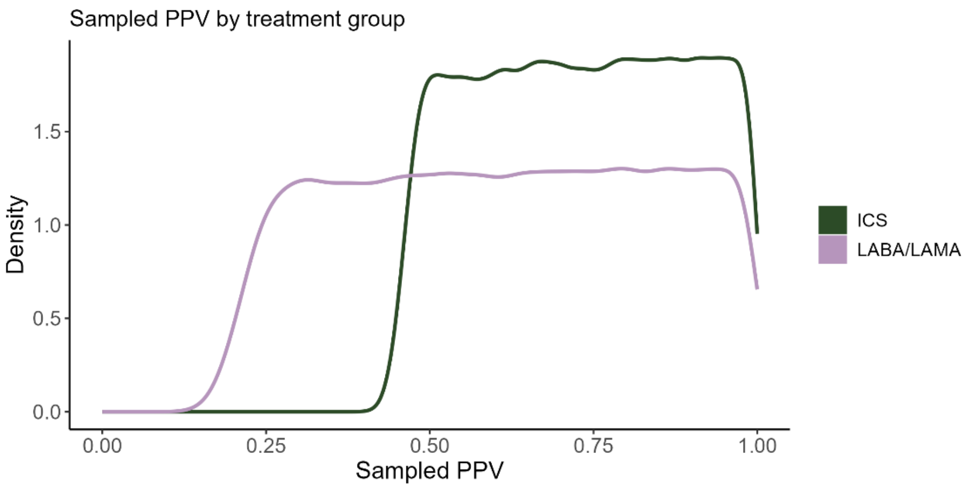

Sampled negative predictive values by treatment group for summary-level probabilistic bias analysis for COVID-19 hospitalisations

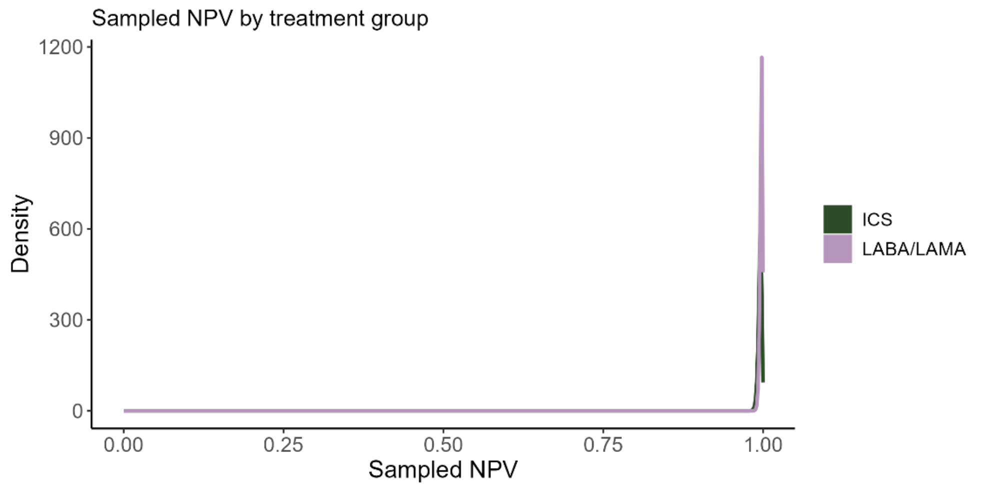

Distribution of risk ratios after adjusting for outcome misclassification using summary-level probabilistic bias analysis for COVID-19 hospitalisations

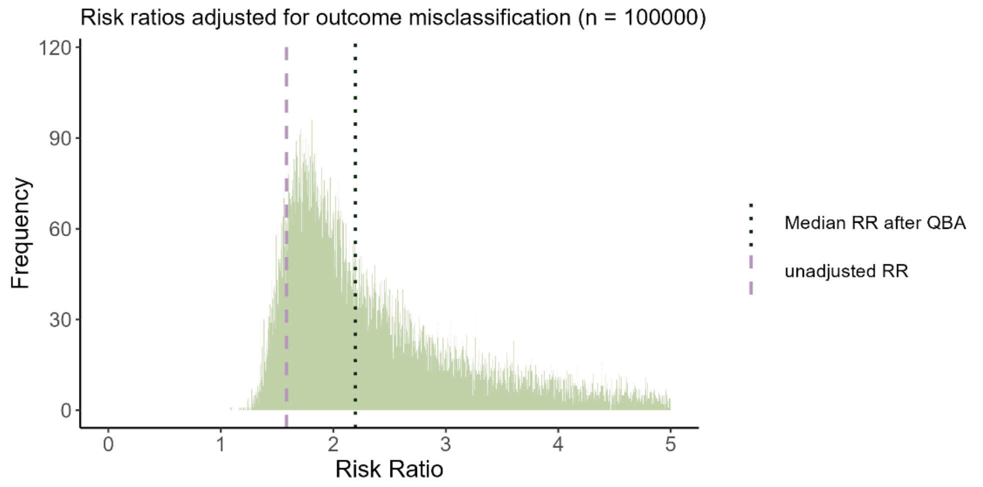

Distribution of odds ratios after adjusting for outcome misclassification using summary-level probabilistic bias analysis for COVID-19 hospitalisations

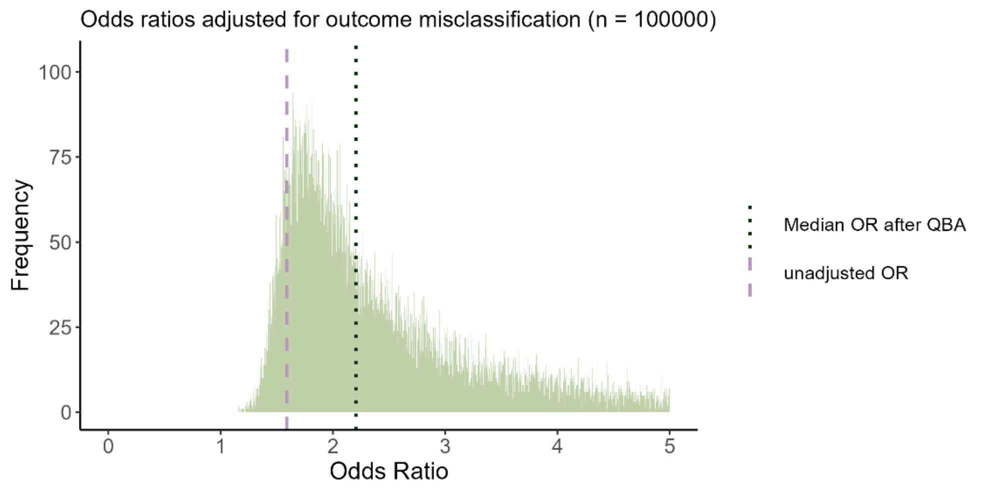

#### S 4.8. Record-level probabilistic bias analysis for COVID-19 hospitalisations (main analysis)

Sampled outcome prevalence by treatment group for record-level probabilistic bias analysis for COVID-19 hospitalisations

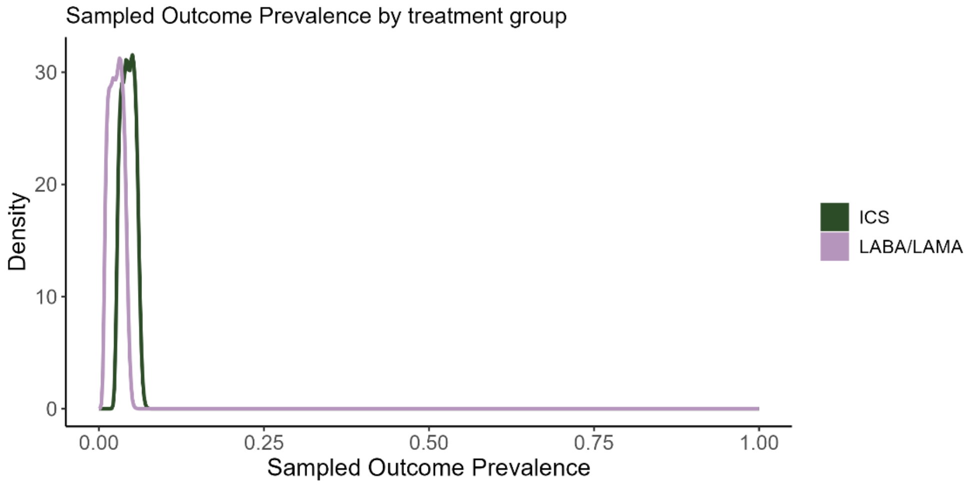

Sampled positive predictive values by treatment group for record-level probabilistic bias analysis for COVID-19 hospitalisations

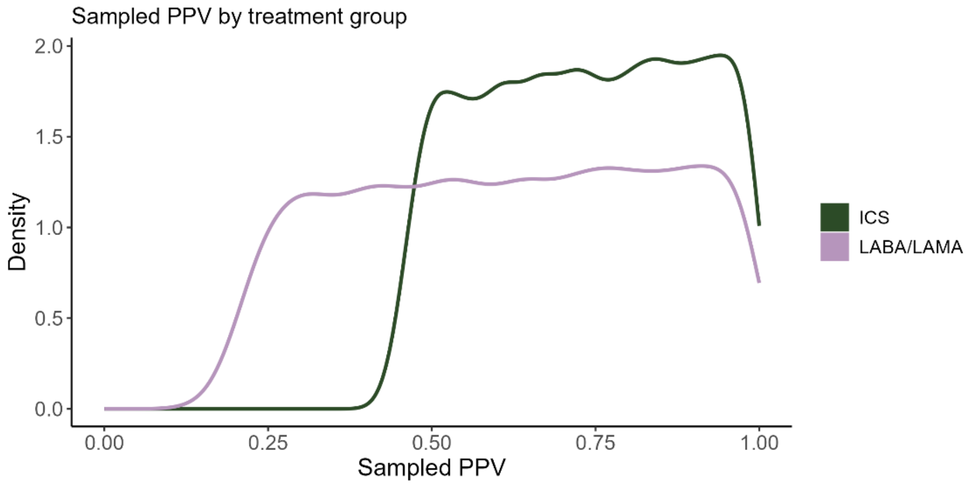

Sampled negative predictive values by treatment group for record-level probabilistic bias analysis for COVID-19 hospitalisations

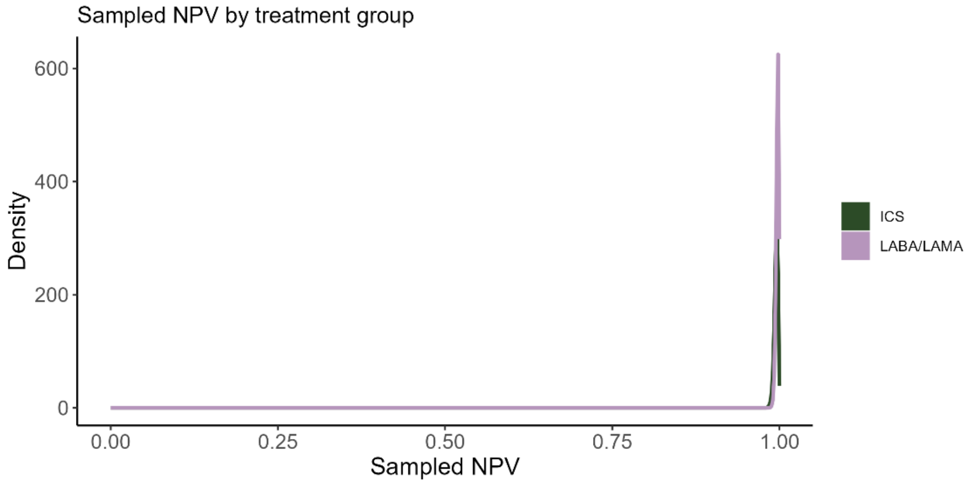

Distribution of odds ratios after adjusting for outcome misclassification using record-level probabilistic bias analysis for COVID-19 hospitalisations, unweighted

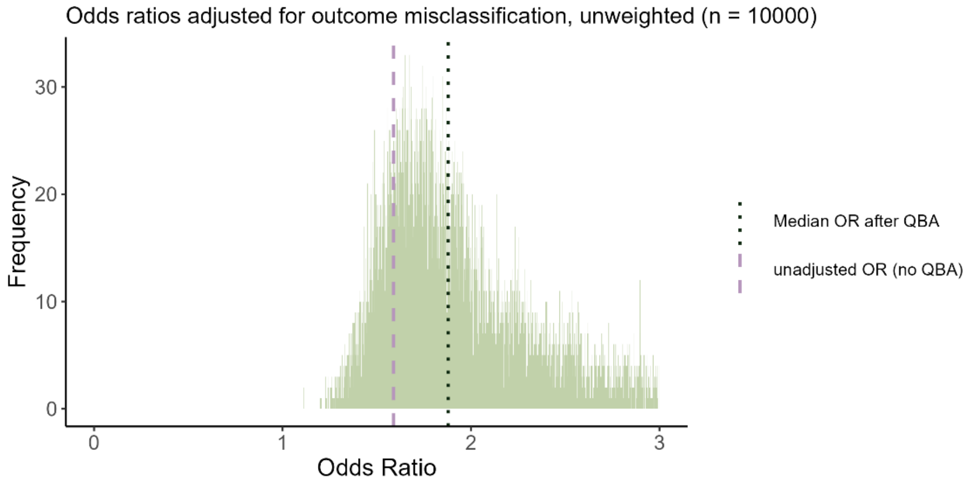

Distribution of odds ratios after adjusting for outcome misclassification using record-level probabilistic bias analysis for COVID-19 hospitalisations, inverse probability of treatment weighted

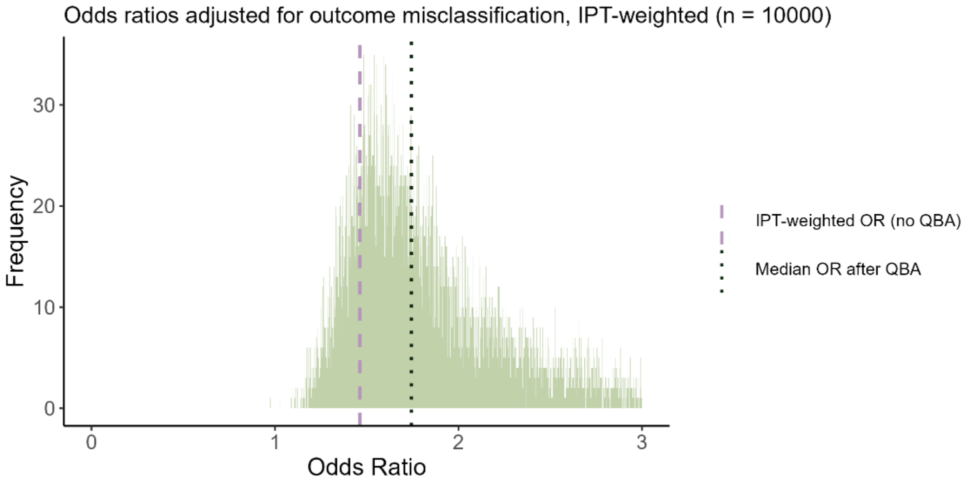

#### S 4.9.	Summary-level probabilistic bias analysis for deaths (main analysis)

Sampled outcome prevalence by treatment group for summary-level probabilistic bias analysis for COVID-19 deaths

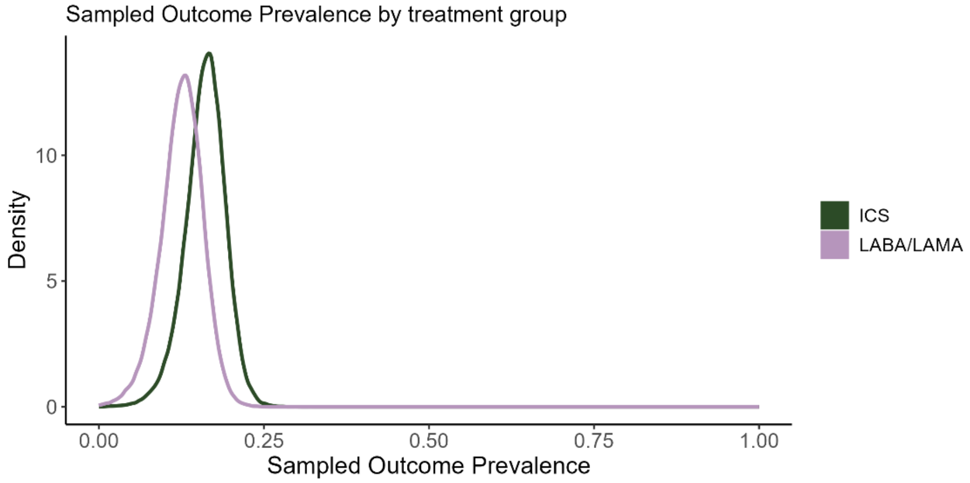

Sampled positive predictive values by treatment group for summary-level probabilistic bias analysis for COVID-19 deaths

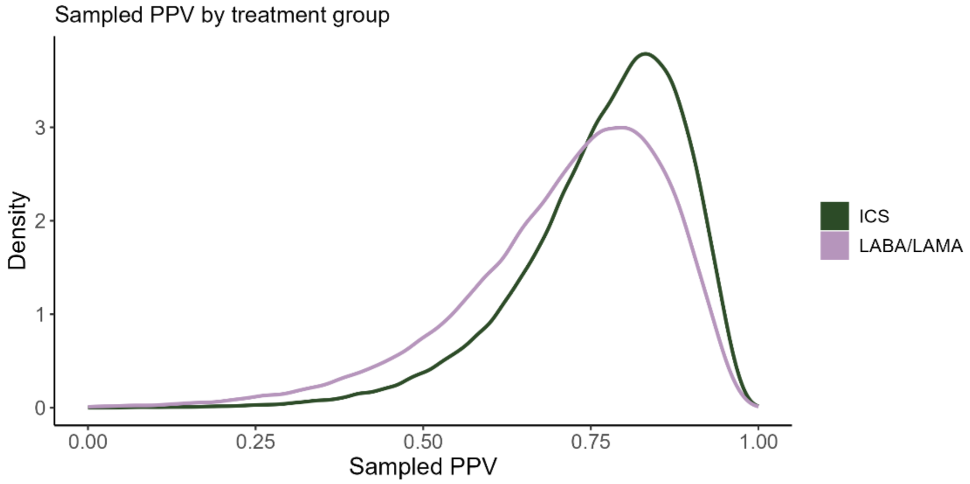

Sampled negative predictive values by treatment group for summary-level probabilistic bias analysis for COVID-19 deaths

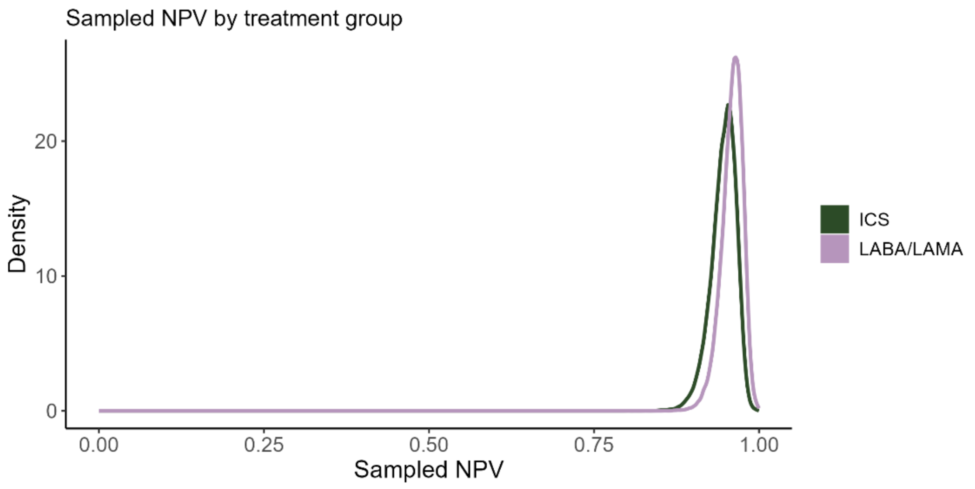

Distribution of risk ratios after adjusting for outcome misclassification using summary-level probabilistic bias analysis for COVID-19 deaths

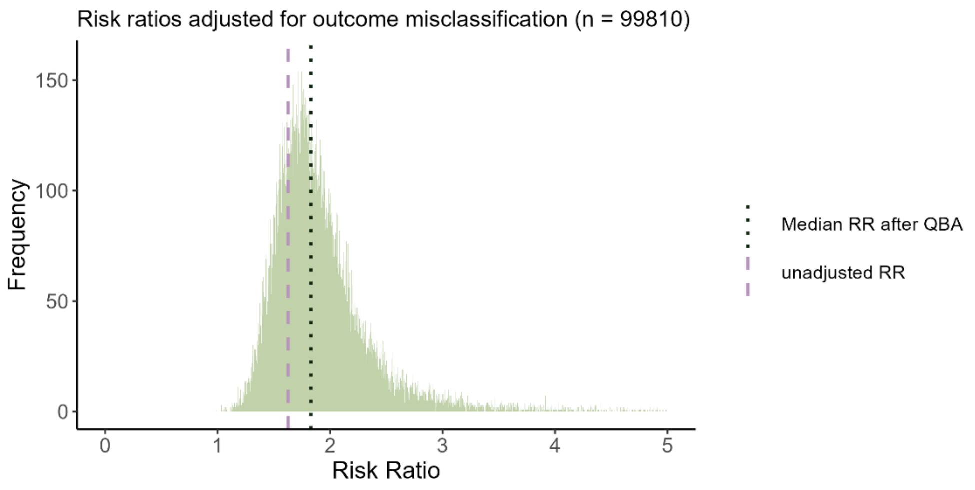

Distribution of odds ratios after adjusting for outcome misclassification using summary-level probabilistic bias analysis for COVID-19 deaths

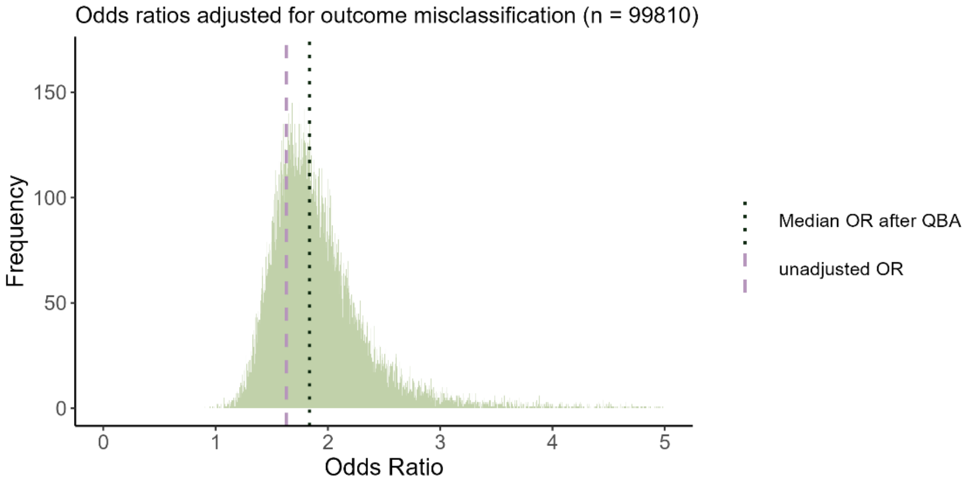

#### S 4.10. Record-level probabilistic bias analysis for COVID-19 deaths (main analysis)

Sampled outcome prevalence by treatment group for record-level probabilistic bias analysis for COVID-19 deaths

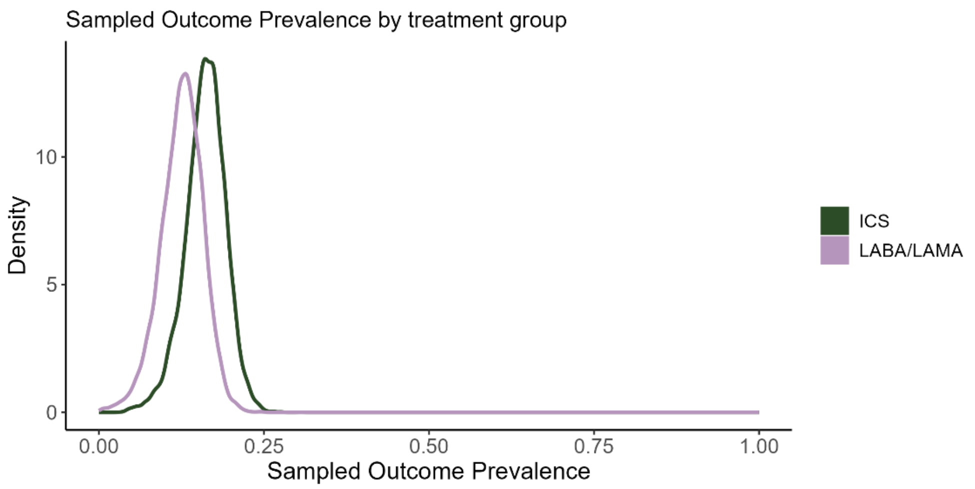

Sampled positive predictive values by treatment group for record-level probabilistic bias analysis for COVID-19 deaths

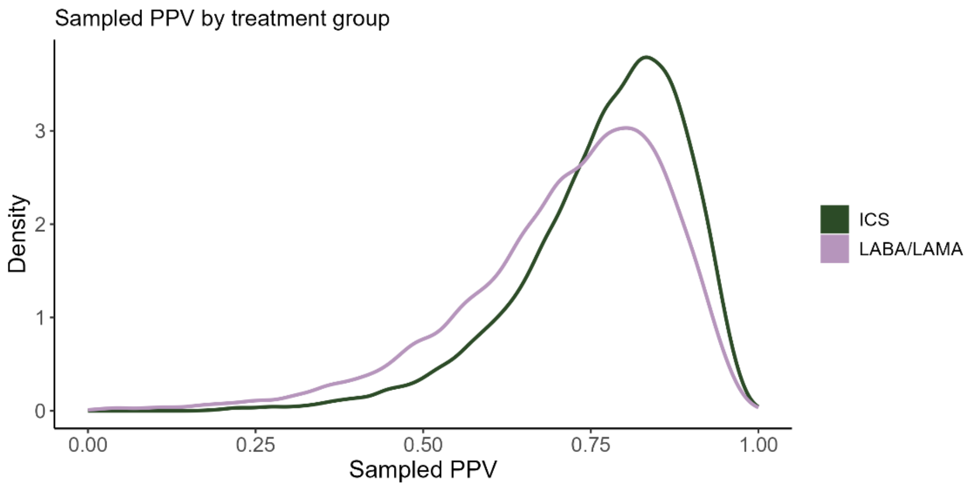

Sampled negative predictive values by treatment group for record-level probabilistic bias analysis for COVID-19 deaths

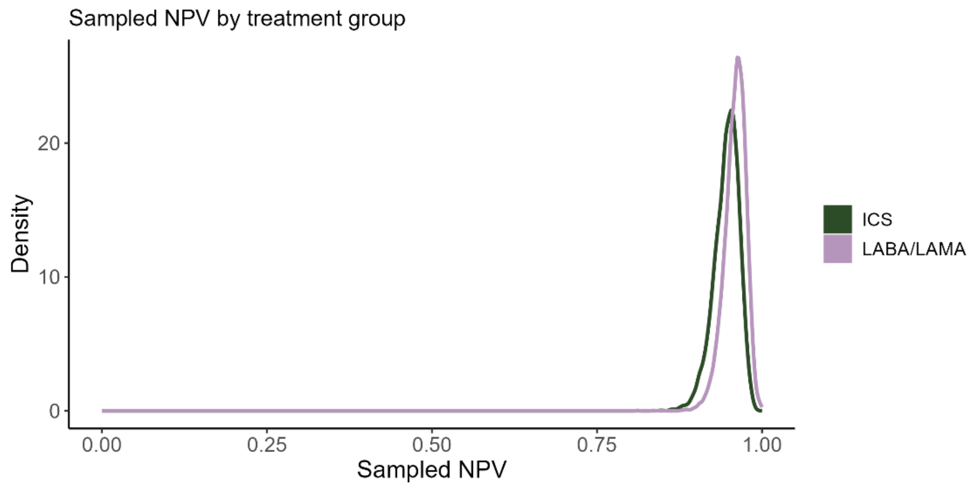

Distribution of odds ratios after adjusting for outcome misclassification using record-level probabilistic bias analysis for COVID-19 deaths, unweighted

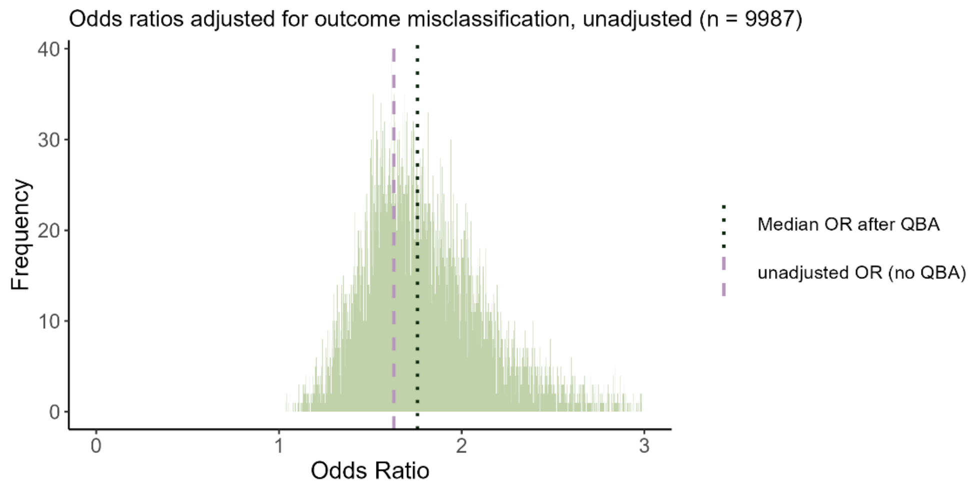

Distribution of odds ratios after adjusting for outcome misclassification using record-level probabilistic bias analysis for COVID-19 deaths, inverse probability of treatment weighted

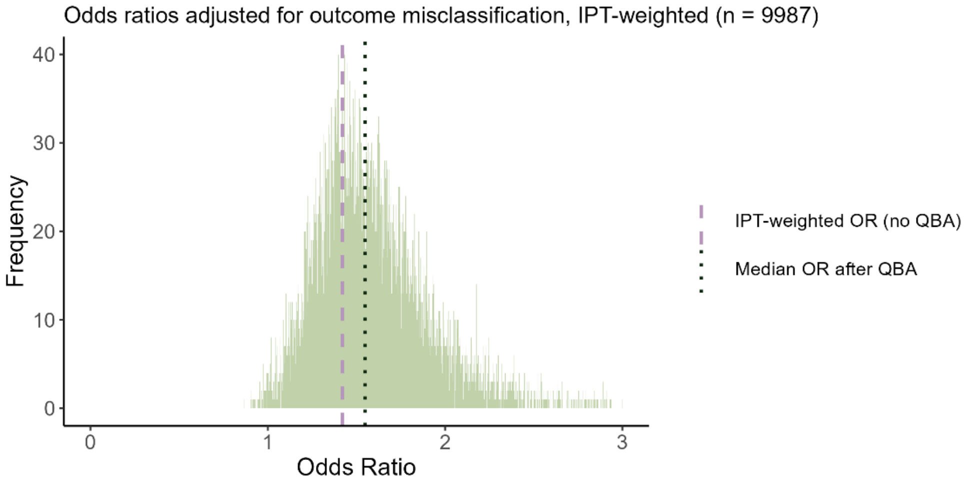

#### S 5. Sensitivity analysis excluding patients using triple therapy at baseline

##### S 5.1. Baseline characteristics

Table S5.1.1 Baseline characteristics of the cohort excluding people using triple therapy at baseline by treatment group, before and after IPT-weighting

| | **Unweighted** | | **After IPT-weighting** | |
|---|---|---|---|---|
| | ICS **N = 14906[1]** | LABA/LAMA **N = 22319[1]** | ICS **N = 14906[1]** | LABA/LAMA **N = 22310[1]** |
| Age | | | | |
| *Mean (SD)* | 71.37 (11.29) | 70.82 (10.23) | 71.02 (11.25) | 71.04 (10.26) |
| *Median (25%-75%)* | 71.67 (63.67-79.67) | 71.67 (63.67-77.67) | 71.67 (63.67-78.67) | 71.67 (63.67-77.67) |
| Gender | | | | |
| *Male* | 7,817 (52%) | 12,245 (55%) | 8,030 (54%) | 12,030 (54%) |
| *Female* | 7,089 (48%) | 10,074 (45%) | 6,876 (46%) | 10,280 (46%) |
| BMI | | | | |
| *Underweight (<18.5)* | 619 (4.2%) | 970 (4.3%) | 4,614 (31%) | 6,906 (31%) |
| *Normal (18.5-24.9)* | 4,629 (31%) | 6,926 (31%) | 632 (4.2%) | 950 (4.3%) |
| *Overweight (25-29.9)* | 5,021 (34%) | 7,172 (32%) | 4,877 (33%) | 7,306 (33%) |
| *Obese (>=30)* | 4,637 (31%) | 7,251 (32%) | 4,783 (32%) | 7,148 (32%) |
| Ethnicity | | | | |
| *White* | 12,891 (86%) | 19,584 (88%) | 13,012 (87%) | 19,469 (87%) |
| *South Asian* | 292 (2.0%) | 197 (0.9%) | 196 (1.3%) | 299 (1.3%) |
| *Black* | 138 (0.9%) | 130 (0.6%) | 110 (0.7%) | 166 (0.7%) |
| *Mixed* | 44 (0.3%) | 49 (0.2%) | 40 (0.3%) | 59 (0.3%) |
| *Unknown* | 1,541 (10%) | 2,359 (11%) | 1,548 (10%) | 2,317 (10%) |
| Smoking | | | | |
| *Current smoking* | 5,964 (40%) | 10,073 (45%) | 2,027 (14%) | 3,028 (14%) |
| *Former smoking* | 8,942 (60%) | 12,246 (55%) | 2,565 (17%) | 3,829 (17%) |
| Index of Multiple Deprivation | | | 2,742 (18%) | 4,105 (18%) |
| *1* | 2,021 (14%) | 3,048 (14%) | 3,393 (23%) | 5,088 (23%) |
| *2* | 2,562 (17%) | 3,813 (17%) | 4,179 (28%) | 6,258 (28%) |
| *3* | 2,759 (19%) | 4,077 (18%) | 1 (<0.1%) | 1 (<0.1%) |
| *4* | 3,473 (23%) | 5,012 (22%) | 3,677 (25%) | 5,515 (25%) |
| *5* | 4,090 (27%) | 6,357 (28%) | 7,626 (51%) | 11,406 (51%) |
| *Missing* | 1 (<0.1%) | 12 (<0.1%) | 4,379 (29%) | 6,555 (29%) |
| Diabetes | 3,685 (25%) | 5,517 (25%) | 2,899 (19%) | 4,337 (19%) |
| Hypertension | 7,689 (52%) | 11,319 (51%) | 2,723 (18%) | 4,080 (18%) |
| Cardiovascular disease | 4,388 (29%) | 6,540 (29%) | 4,547 (31%) | 6,798 (30%) |
| Cancer | 2,827 (19%) | 4,416 (20%) | 186 (1.2%) | 278 (1.2%) |
| Past asthma | 4,135 (28%) | 2,665 (12%) | 11,761 (79%) | 17,607 (79%) |
| Kidney impairment | 4,567 (31%) | 6,738 (30%) | 1,900 (13%) | 2,842 (13%) |
| Immunosuppression | 188 (1.3%) | 277 (1.2%) | 4,368 (29%) | 6,541 (29%) |
| Influenza vaccine | 11,390 (76%) | 17,962 (80%) | | |
| Pneumococcal vaccine | 1,497 (10%) | 3,237 (15%) | 6,438 (43%) | 9,628 (43%) |
| Any exacerbation in past 12 months | 4,638 (31%) | 6,222 (28%) | 8,469 (57%) | 12,681 (57%) |
| [1]n (%) | | | | |

#### S 5.2. Observed outcomes by treatment group, excluding patients using triple therapy at baseline

Table S5.2.1 Observed outcomes by treatment group, excluding patients using triple therapy at baseline.

| | **ICS**<br>N = 14906[1] | **LABA/LAMA**<br>N = 22319[1] |
|---|---|---|
| COVID-19 hospitalisation | 107 (0.7%) | 133 (0.6%) |
| COVID-19 death | 62 (0.4%) | 72 (0.3%) |
| All-cause mortality | 473 (3.2%) | 574 (2.6%) |
| [1]n (%) | | |

#### S 5.3. Log residuals (excluding triple therapy users)

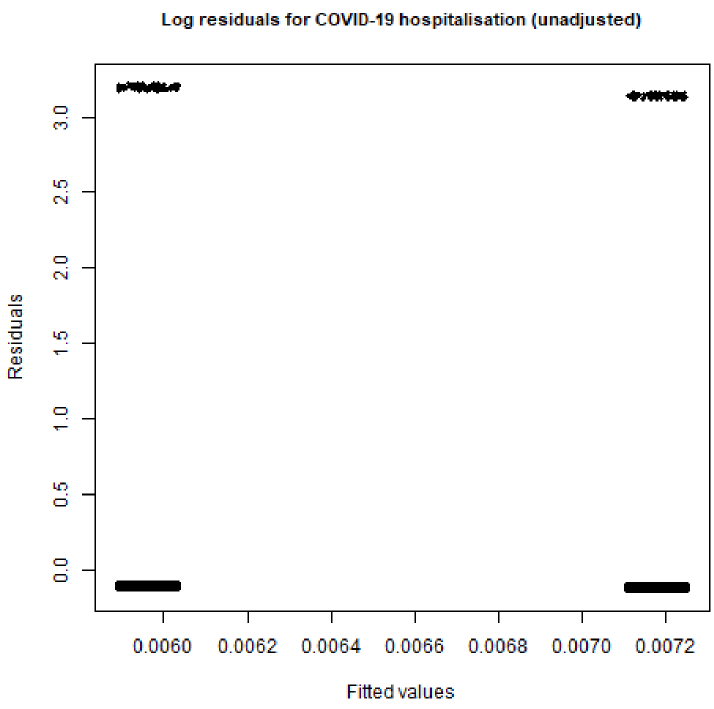

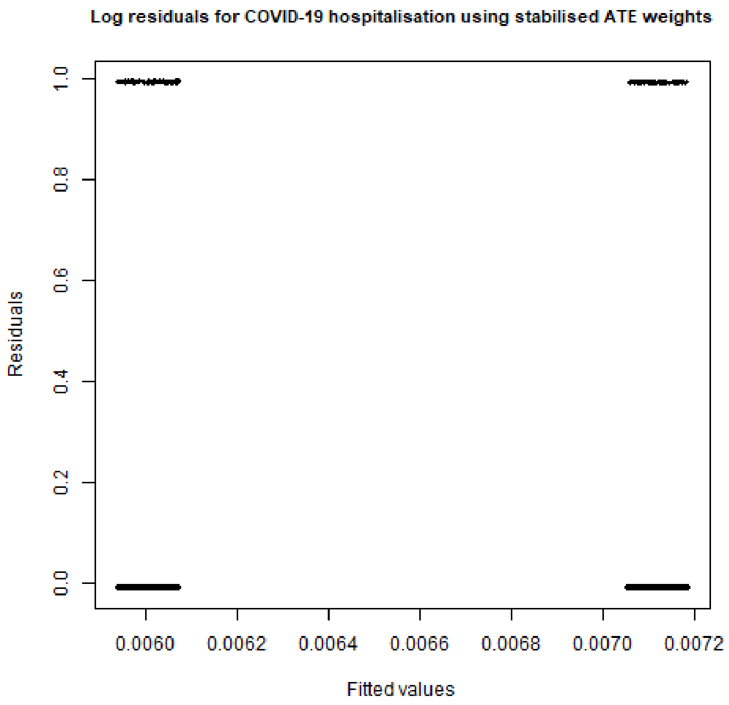
Log residuals for COVID-19 hospitalisation using stabilised ATE weights
Residuals
1.0
0.8
0.6
0.4
0.2
0.0
0.0060
0.0062
0.0064
0.0066
0.0068
0.0070
0.0072
Fitted values

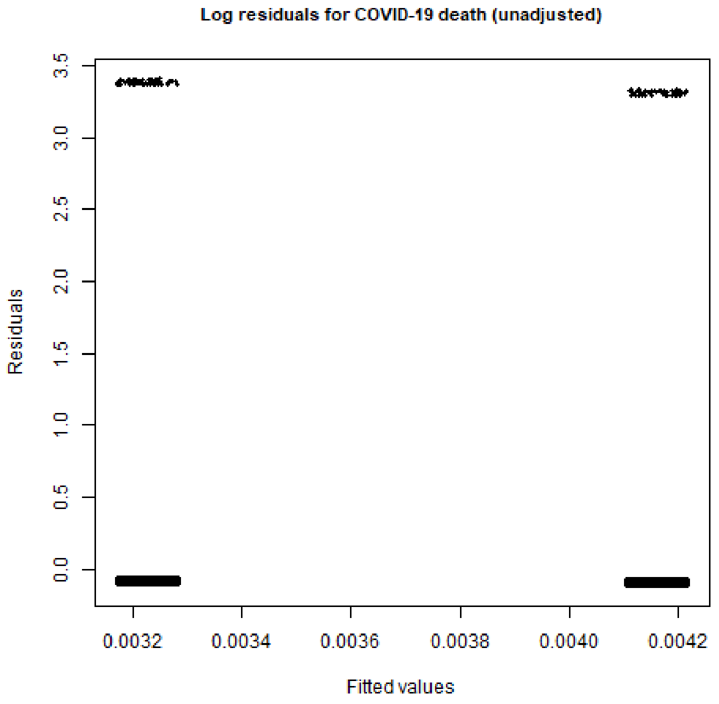
Log residuals for COVID-19 death (unadjusted)
Residuals
3.5
3.0
2.5
2.0
1.5
1.0
0.5
0.0
0.0032
0.0034
0.0036
0.0038
0.0040
0.0042
Fitted values

Log residuals for COVID-19 death using stabilised ATE weights

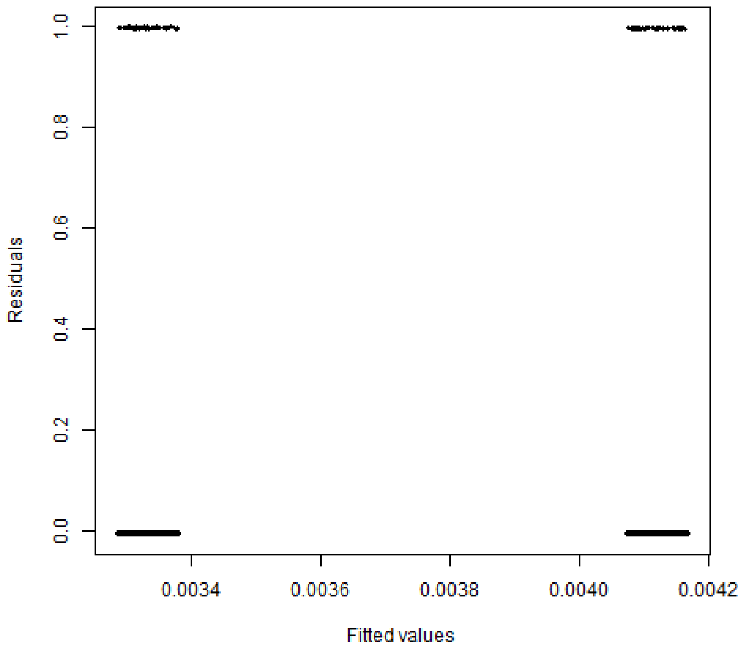

Log residuals for all-cause mortality (unadjusted)

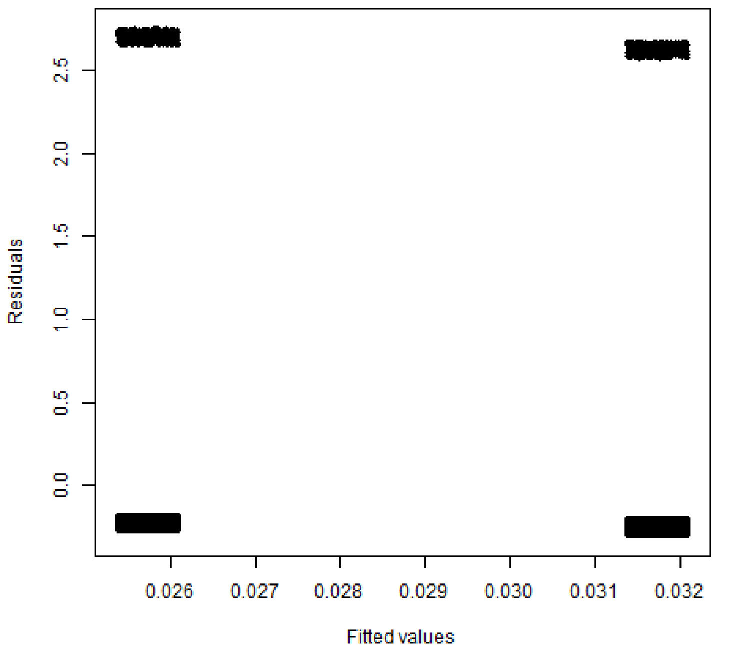

**Log residuals for all-cause mortality using stabilised ATE weights**

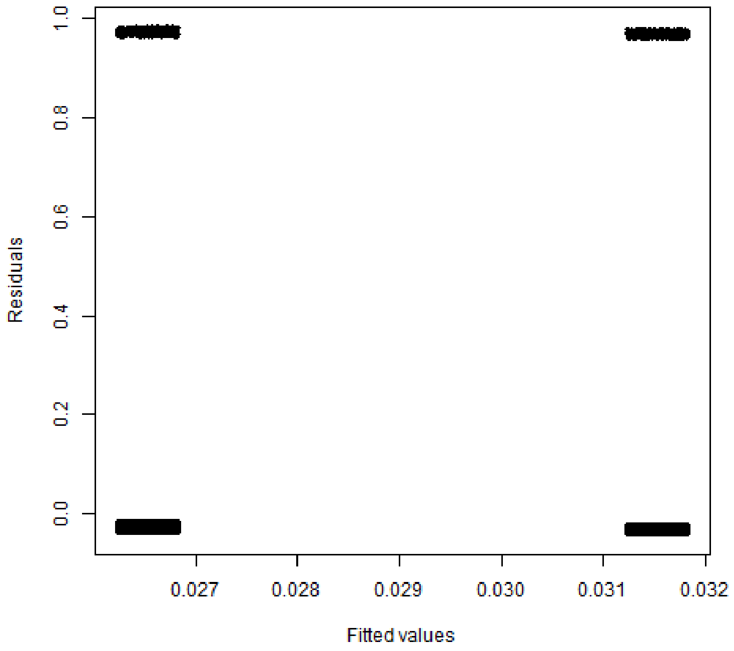

#### S 5.4. Summary-level probabilistic bias analysis for COVID-19 hospitalisations (excluding triple therapy users)

Sampled outcome prevalence by treatment group for summary-level probabilistic bias analysis for COVID-19 hospitalisations, excluding patients using triple therapy at baseline

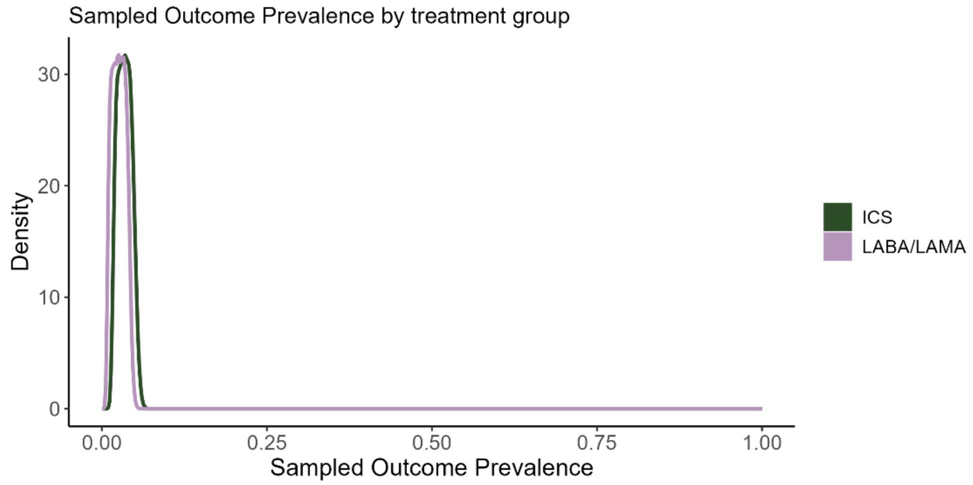

Sampled positive predictive values by treatment group for summary-level probabilistic bias analysis for COVID-19 hospitalisations, excluding patients using triple therapy at baseline

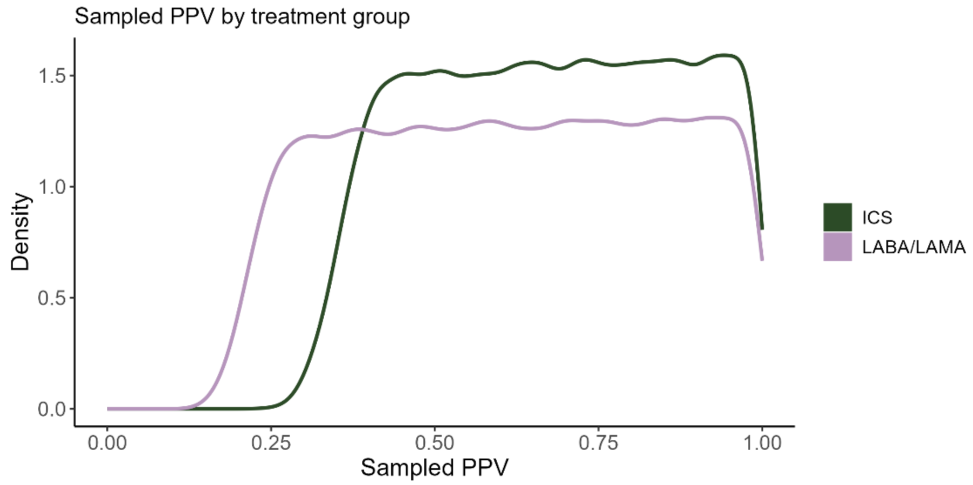

Sampled negative predictive values by treatment group for summary-level probabilistic bias analysis for COVID-19 hospitalisations, excluding patients using triple therapy at baseline

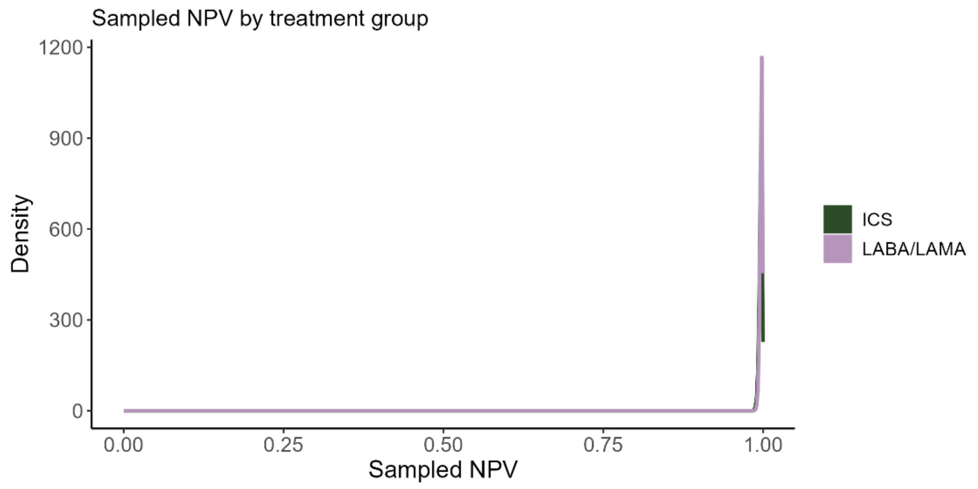

Distribution of risk ratios after adjusting for outcome misclassification using summary-level probabilistic bias analysis for COVID-19 hospitalisations, excluding patients using triple therapy at baseline

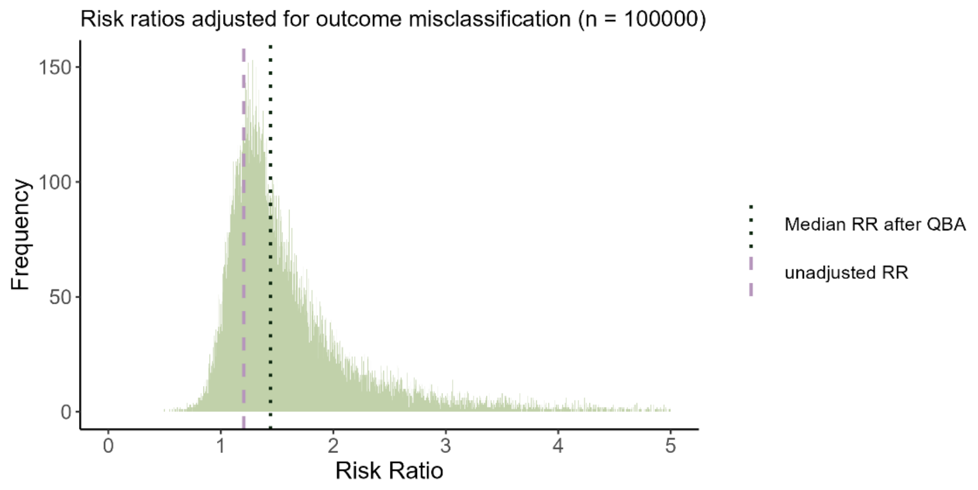

Distribution of odds ratios after adjusting for outcome misclassification using summary-level probabilistic bias analysis for COVID-19 hospitalisations, excluding patients using triple therapy at baseline

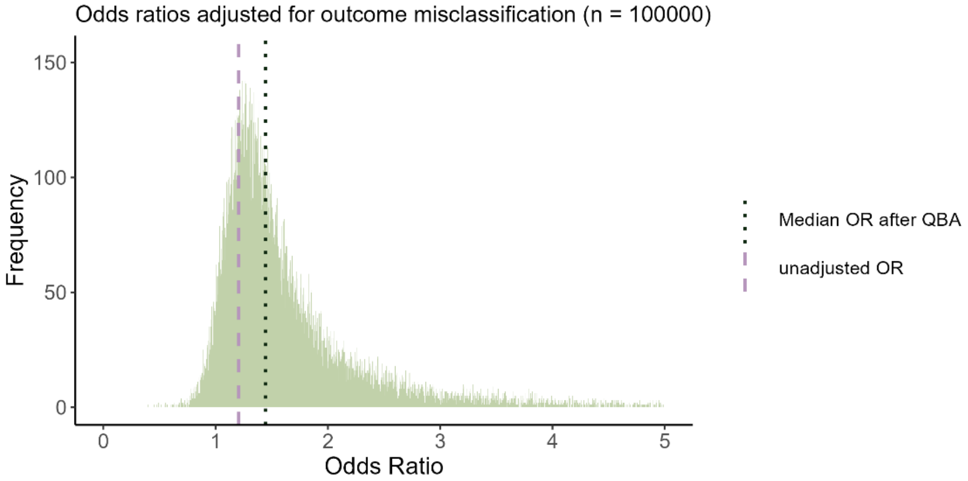

#### S 5.5. Record-level probabilistic bias analysis for hospitalisations (excluding triple therapy users)

Sampled outcome prevalence by treatment group for record-level probabilistic bias analysis for COVID-19 hospitalisations, excluding patients using triple therapy at baseline

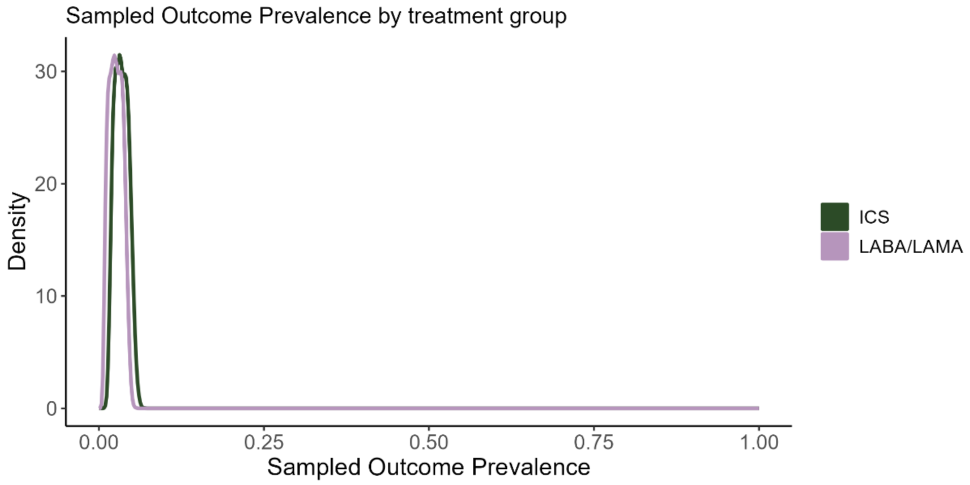

Sampled positive predictive values by treatment group for record-level probabilistic bias analysis for COVID-19 hospitalisations, excluding patients using triple therapy at baseline

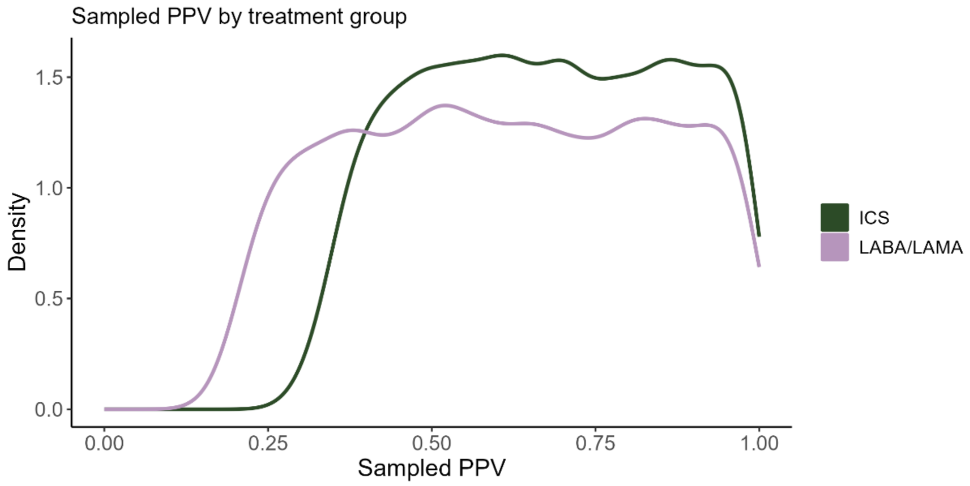

Sampled negative predictive values by treatment group for record-level probabilistic bias analysis for COVID-19 hospitalisations, excluding patients using triple therapy at baseline

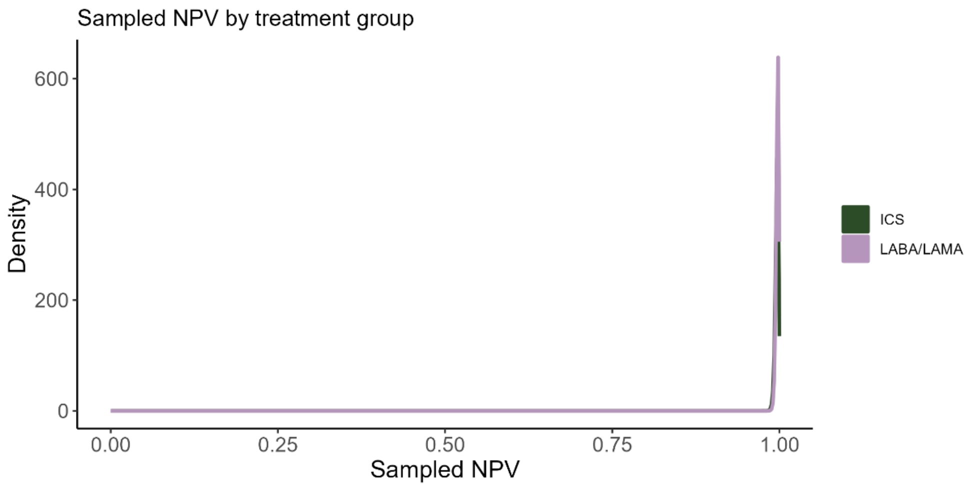

Distribution of odds ratios after adjusting for outcome misclassification using record-level probabilistic bias analysis for COVID-19 hospitalisations, excluding patients using triple therapy at baseline, unweighted

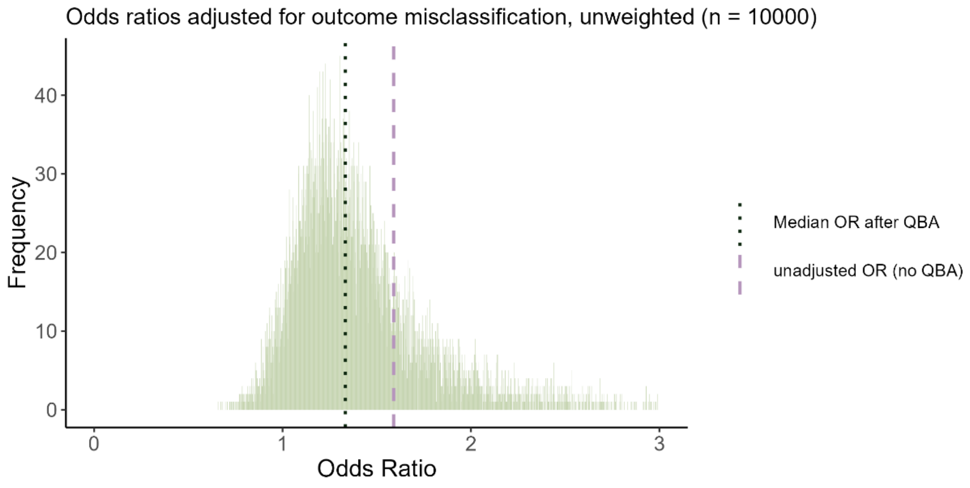

Distribution of odds ratios after adjusting for outcome misclassification using record-level probabilistic bias analysis for COVID-19 hospitalisations, excluding patients using triple therapy at baseline, inverse probability of treatment weighted

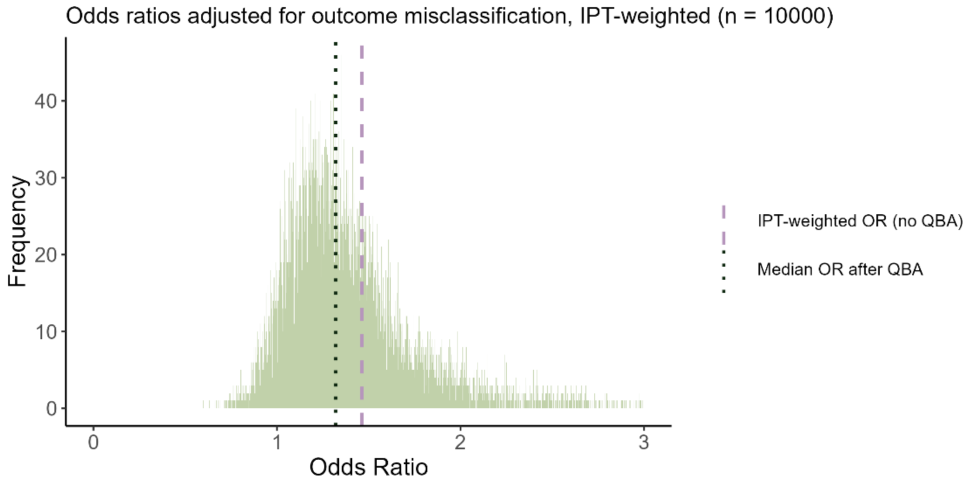

#### S 5.6. Summary-level probabilistic bias analysis for COVID-19 deaths (excluding triple therapy users)

Sampled outcome prevalence by treatment group for summary-level probabilistic bias analysis for COVID-19 deaths, excluding patients using triple therapy at baseline

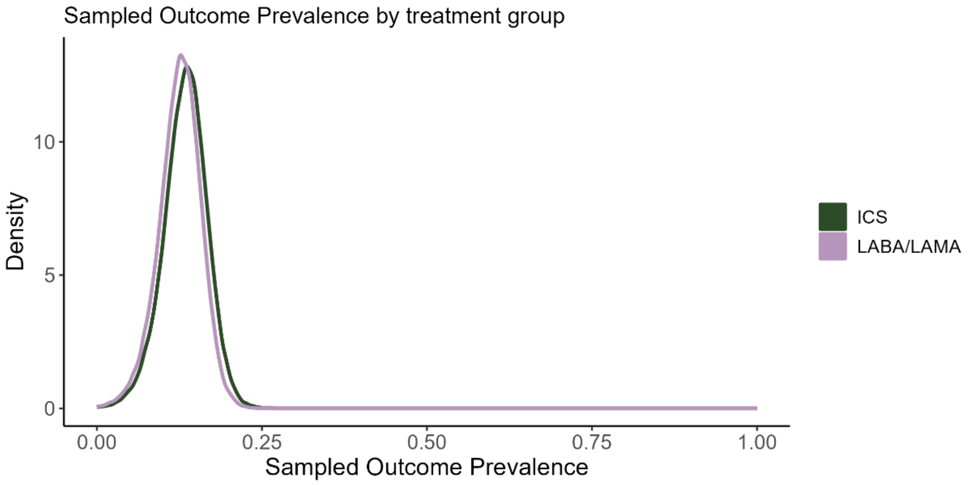

Sampled positive predictive values by treatment group for summary-level probabilistic bias analysis for COVID-19 deaths, excluding patients using triple therapy at baseline

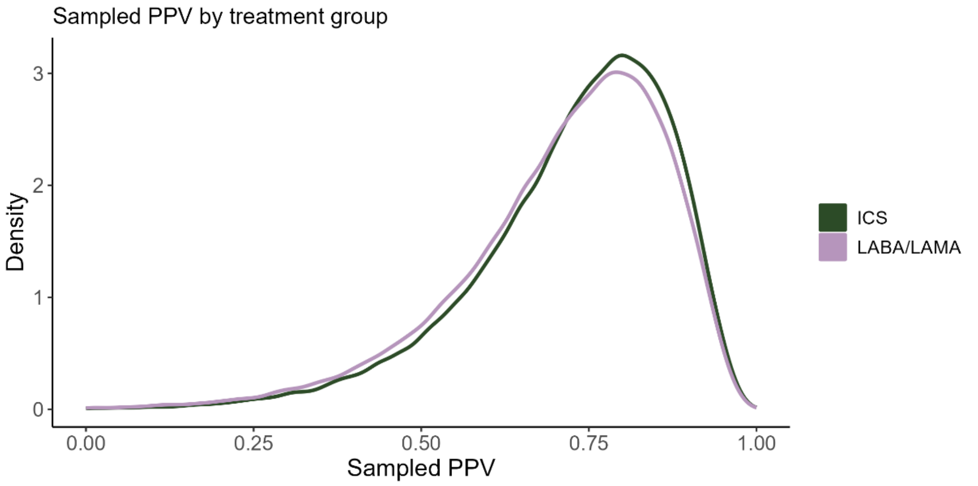

Sampled negative predictive values by treatment group for summary-level probabilistic bias analysis for COVID-19 deaths, excluding patients using triple therapy at baseline

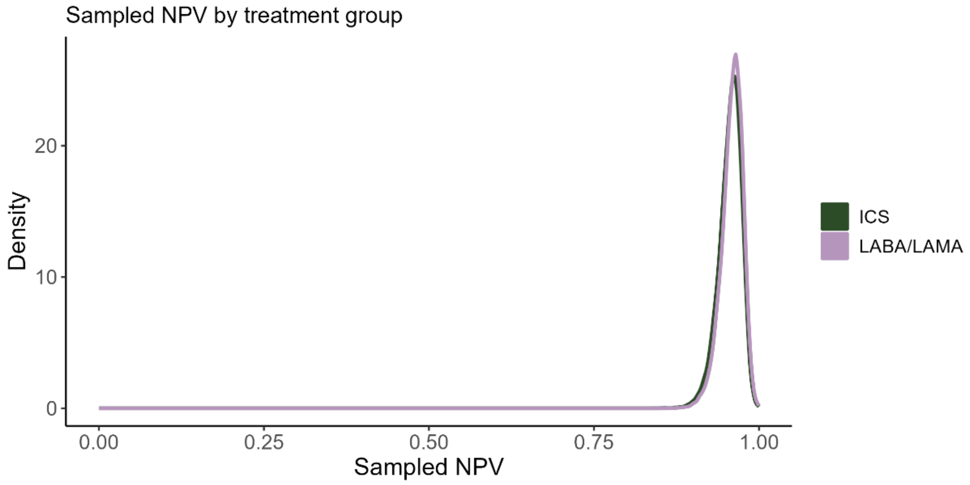

Distribution of risk ratios after adjusting for outcome misclassification using summary-level probabilistic bias analysis for COVID-19 deaths, excluding patients using triple therapy at baseline

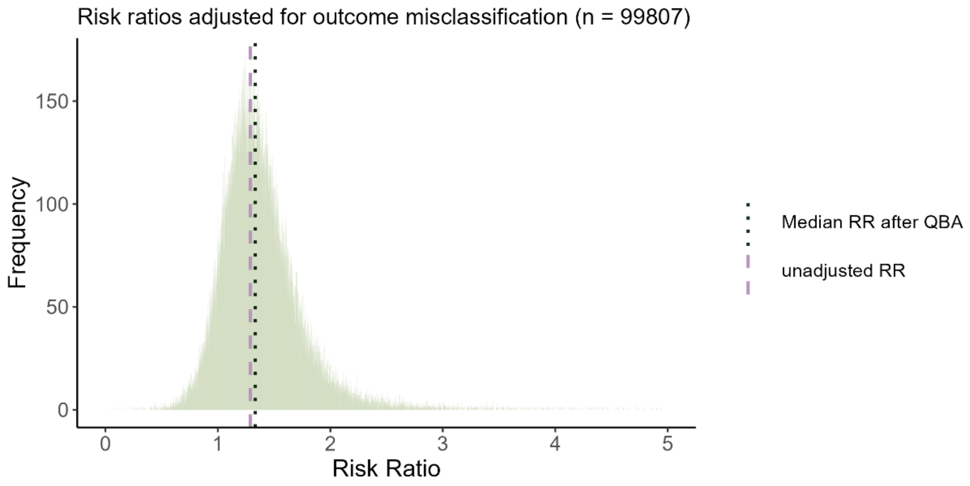

Distribution of odds ratios after adjusting for outcome misclassification using summary-level probabilistic bias analysis for COVID-19 deaths, excluding patients using triple therapy at baseline

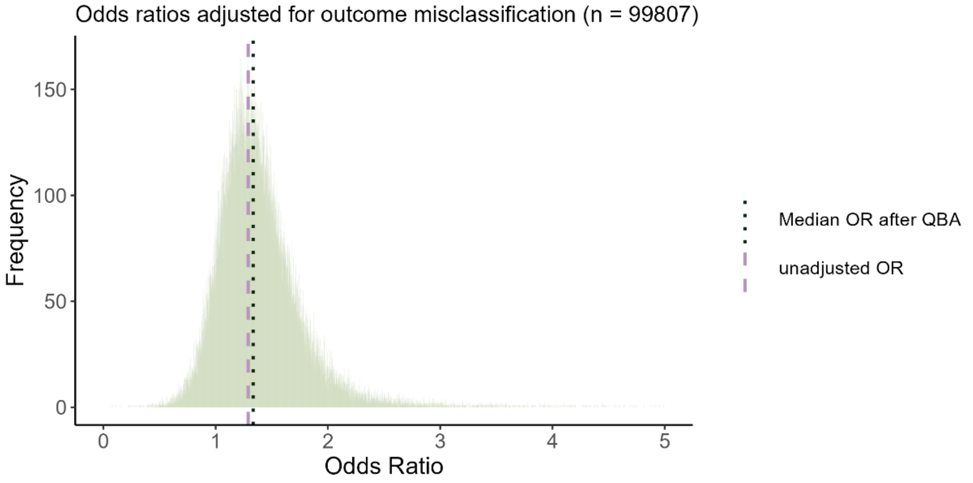

#### S 5.7. Record-level probabilistic bias analysis for COVID-19 deaths (excluding triple therapy users)

Sampled outcome prevalence by treatment group for record-level probabilistic bias analysis for COVID-19 deaths, excluding patients using triple therapy at baseline

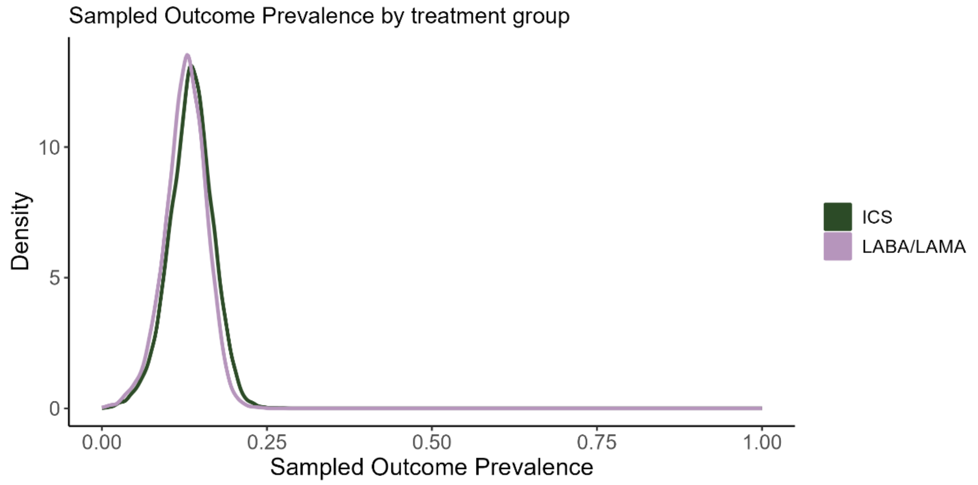

Sampled positive predictive values by treatment group for record-level probabilistic bias analysis for COVID-19 deaths, excluding patients using triple therapy at baseline

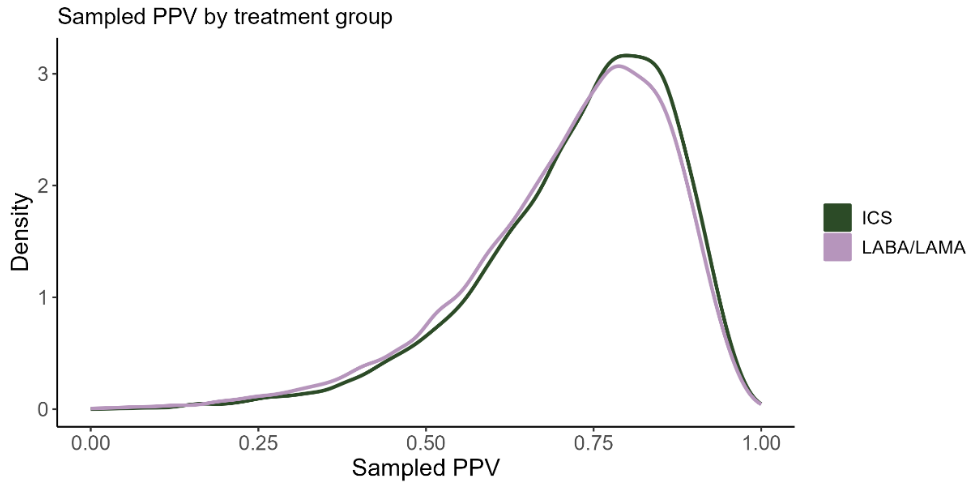

Sampled negative predictive values by treatment group for record-level probabilistic bias analysis for COVID-19 deaths, excluding patients using triple therapy at baseline

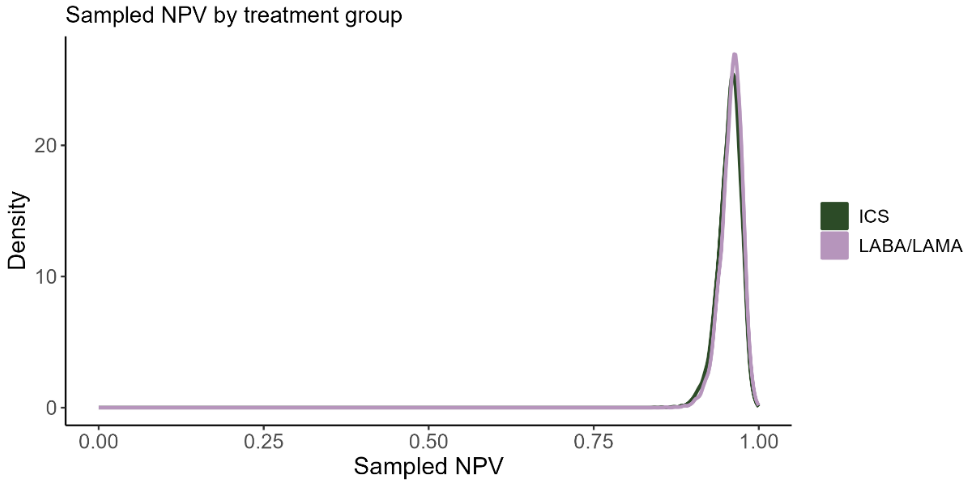

Distribution of odds ratios after adjusting for outcome misclassification using record-level probabilistic bias analysis for COVID-19 deaths, excluding patients using triple therapy at baseline, unweighted

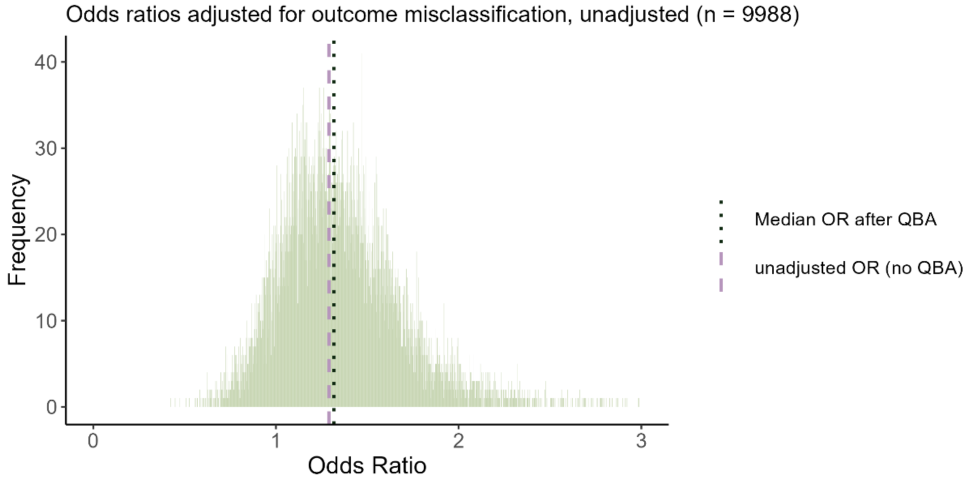

Distribution of odds ratios after adjusting for outcome misclassification using record-level probabilistic bias analysis for COVID-19 deaths, excluding patients using triple therapy at baseline, inverse probability of treatment weighted

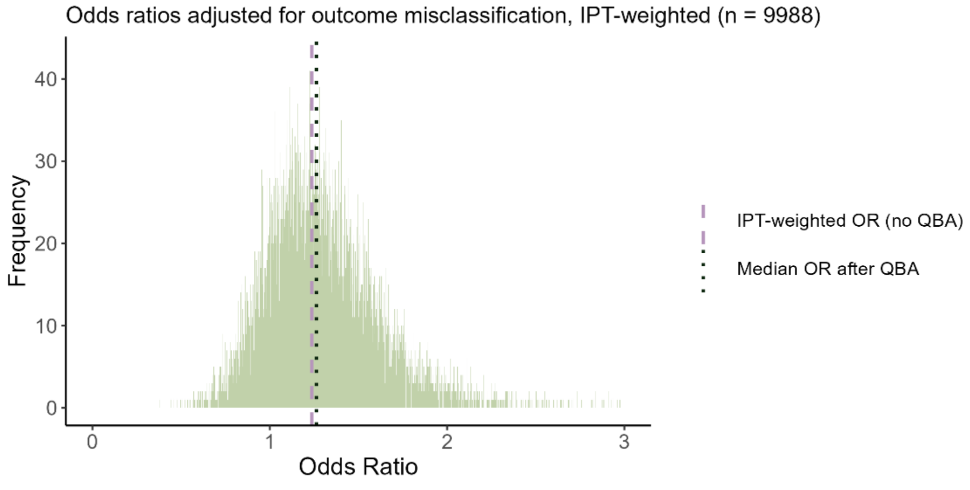

### S 6. Other sensitivity analyses

#### S 6.1. Including people with asthma or other respiratory disease

Baseline characteristics of the cohort, including people with asthma or other chronic respiratory disease

| | **ICS**<br>N = 123697[1] | **LABA/LAMA**<br>N = 27580[1] |
|---|---|---|
| Age | | |
| *Mean (SD)* | 70.39 (11.03) | 71.06 (10.28) |
| *Median (25%-75%)* | 71.67 (62.67-78.67) | 71.67 (64.67-78.67) |
| Gender | | |
| *Male* | 61,538 (50%) | 15,161 (55%) |
| *Female* | 62,159 (50%) | 12,419 (45%) |
| BMI | | |
| *Underweight (<18.5)* | 6,000 (4.9%) | 1,214 (4.4%) |
| *Normal (18.5-24.9)* | 38,042 (31%) | 8,581 (31%) |
| *Overweight (25-29.9)* | 38,565 (31%) | 8,905 (32%) |
| *Obese (>=30)* | 41,090 (33%) | 8,880 (32%) |
| Ethnicity | | |
| *White* | 108,615 (88%) | 24,172 (88%) |
| *South Asian* | 2,625 (2.1%) | 310 (1.1%) |
| *Black* | 1,132 (0.9%) | 183 (0.7%) |
| *Mixed* | 391 (0.3%) | 61 (0.2%) |
| *Unknown* | 10,934 (8.8%) | 2,854 (10%) |
| Smoking | | |
| *Current smoking* | 45,361 (37%) | 12,002 (44%) |
| *Former smoking* | 78,336 (63%) | 15,578 (56%) |
| Index of Multiple Deprivation | | |
| *1* | 16,108 (13%) | 3,748 (14%) |
| *2* | 20,459 (17%) | 4,680 (17%) |
| *3* | 22,322 (18%) | 5,061 (18%) |
| *4* | 27,654 (22%) | 6,201 (22%) |
| *5* | 37,073 (30%) | 7,873 (29%) |
| *Missing* | 81 (<0.1%) | 17 (<0.1%) |
| Diabetes | 32,544 (26%) | 7,007 (25%) |
| Hypertension | 62,479 (51%) | 14,072 (51%) |
| Cardiovascular disease | 36,617 (30%) | 8,366 (30%) |
| Cancer | 24,946 (20%) | 6,179 (22%) |
| Past asthma | 67,719 (55%) | 4,906 (18%) |
| Current asthma | 51,864 (42%) | 2,534 (9.2%) |
| Kidney impairment | 36,177 (29%) | 8,541 (31%) |
| Immunosuppression | 1,627 (1.3%) | 373 (1.4%) |
| Influenza vaccine | 60,557 (49%) | 19,293 (70%) |
| Pneumococcal vaccine | 5,985 (4.8%) | 3,237 (12%) |
| Any exacerbation in past 12 months | 54,349 (44%) | 8,147 (30%) |

[1]n (%)

Observed outcomes by treatment group, including people with asthma or other chronic respiratory disease

| | ICS N = 123697[1] | LABA/LAMA N = 27580[1] |
|---|---|---|
| COVID-19 hospitalisation | 1,211 (1.0%) | 175 (0.6%) |
| COVID-19 death | 628 (0.5%) | 100 (0.4%) |
| All-cause mortality | 4,297 (3.5%) | 844 (3.1%) |

Forest plot of results of logistic regression for COVID-19 hospitalisation, COVID-19 death, using a cohort including people with asthma or other chronic respiratory diseases. Effect estimates > 1 indicate an increased risk in the ICS group compared to the LABA/LAMA group. IPTW = inverse probability of treatment weighting, in this case using average treatment effect in the population weights.

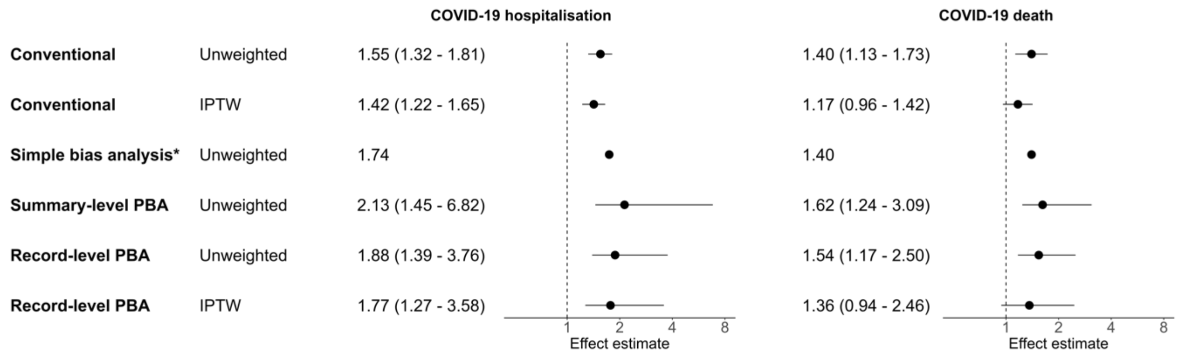

#### S 6.2. Using “6 months post last prescription date” - exposure definition

Baseline characteristics before weighting, defined by a “6 months post-last prescription issue date” exposure definition

| | **ICS**<br>N = 59130[1] | **LABA/LAMA**<br>N = 23201[1] |
|---|---|---|
| Age | | |
| *Mean (SD)* | 71.25 (10.54) | 70.77 (10.29) |
| *Median (25%-75%)* | 71.67 (63.67-78.67) | 71.67 (63.67-77.67) |
| Gender | | |
| *Male* | 31,464 (53%) | 12,717 (55%) |
| *Female* | 27,666 (47%) | 10,484 (45%) |
| BMI | | |
| *Underweight (<18.5)* | 3,298 (5.6%) | 1,014 (4.4%) |
| *Normal (18.5-24.9)* | 19,098 (32%) | 7,198 (31%) |
| *Overweight (25-29.9)* | 18,335 (31%) | 7,454 (32%) |
| *Obese (>=30)* | 18,399 (31%) | 7,535 (32%) |
| Ethnicity | | |
| *White* | 52,053 (88%) | 20,334 (88%) |
| *South Asian* | 799 (1.4%) | 219 (0.9%) |
| *Black* | 384 (0.6%) | 131 (0.6%) |
| *Mixed* | 161 (0.3%) | 50 (0.2%) |
| *Unknown* | 5,733 (9.7%) | 2,467 (11%) |
| Smoking | | |
| *Current smoking* | 24,078 (41%) | 10,562 (46%) |
| *Former smoking* | 35,052 (59%) | 12,639 (54%) |
| Index of Multiple Deprivation | | |
| *1* | 7,608 (13%) | 3,150 (14%) |
| *2* | 9,727 (16%) | 3,926 (17%) |
| *3* | 10,510 (18%) | 4,237 (18%) |
| *4* | 13,446 (23%) | 5,235 (23%) |
| *5* | 17,805 (30%) | 6,639 (29%) |
| *Missing* | 34 (<0.1%) | 14 (<0.1%) |
| Diabetes | 14,908 (25%) | 5,758 (25%) |
| Hypertension | 30,027 (51%) | 11,766 (51%) |
| Cardiovascular disease | 17,681 (30%) | 6,834 (29%) |
| Cancer | 11,141 (19%) | 4,575 (20%) |
| Past asthma | 16,090 (27%) | 2,756 (12%) |
| Kidney impairment | 17,659 (30%) | 7,018 (30%) |
| Immunosuppression | 708 (1.2%) | 290 (1.2%) |
| Influenza vaccine | 47,144 (80%) | 18,577 (80%) |
| Pneumococcal vaccine | 6,311 (11%) | 3,345 (14%) |
| Any exacerbation in past 12 months | 23,839 (40%) | 6,421 (28%) |

[1]n (%)

Observed outcomes by treatment group, using a cohort defined by a “6 months post-last prescription issue date” exposure definition

| | **ICS**<br>N = 59130[1] | **LABA/LAMA**<br>N = 23201[1] |
|---|---|---|

| | | |
|---|---|---|
| COVID-19 hospitalisation | 559 (0.9%) | 139 (0.6%) |
| COVID-19 death | 316 (0.5%) | 76 (0.3%) |
| All-cause mortality | 2,108 (3.6%) | 595 (2.6%) |
| [1]n (%) | | |

Forest plot of results of logistic regression for COVID-19 hospitalisation, COVID-19 death, using a cohort defined by a "6 months post-last prescription issue date" exposure definition. Effect estimates > 1 indicate an increased risk in the ICS group compared to the LABA/LAMA group. IPTW = inverse probability of treatment weighting, in this case using average treatment effect in the population weights.

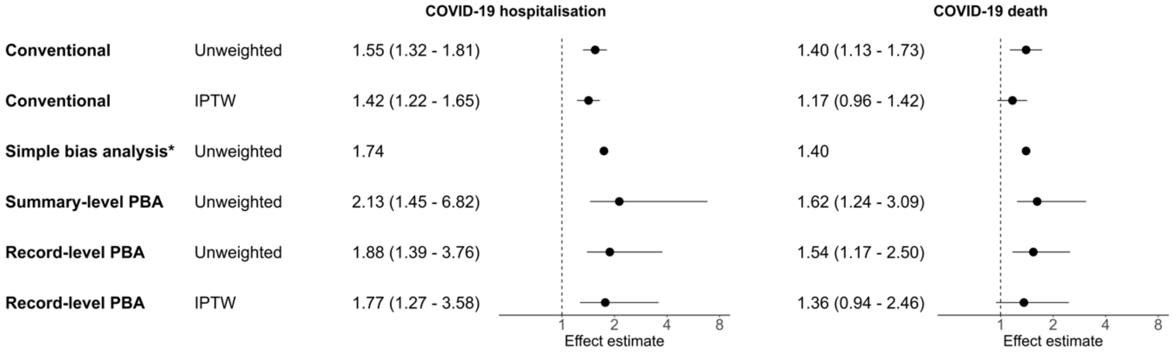

#### S 6.3. Coding all deaths with missing cause of death as COVID-19

Counts of deaths registered in ONS, and deaths missing from ONS, by treatment group

| | ICS | LABA/LAMA | Total |
|---|---|---|---|
| Registered COVID-19 deaths (ONS) | 294 | 72 | 366 |
| Patients with death registered in CPRD but not ONS | 156 | 49 | 205 |
| Total COVID-19 deaths assuming all missing deaths are COVID-19 deaths | 450 | 121 | 571 |
| Increase in COVID-19 deaths assuming all missing deaths were COVID-19 deaths. | 53.1% | 68.1% | 56.0% |

Odds ratios assuming that all deaths missing in ONS would have been COVID-19 deaths. Summary = summary-level probabilistic bias analysis, record = record-level probabilistic bias analysis, IPTW = inverse probability of treatment weighting, in this case using average treatment effect in the population weights.

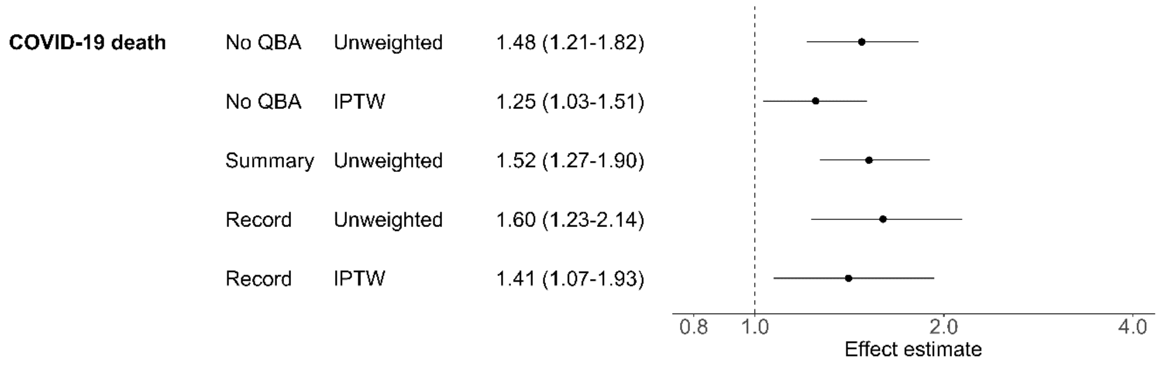

#### S 6.4. Simulating differential misclassification

Heat map showing odds ratios after simulating differential outcome misclassification for COVID-19 deaths. 100,000 simulations were run per combination of sensitivity values. Specificity was set at 0.97. SI = simulation interval

| Sensitivity (control group) \ Sensitivity (ICS) | 0.5 | | | | | 0.6 | | | | | 0.7 | | | | | 0.8 | | | | | 0.9 | | | | | 1.0 |
|---|---|---|---|---|---|---|---|---|---|---|---|---|---|---|---|---|---|---|---|---|---|---|---|---|---|---|
| 1.0 | 5.92 | 5.61 | 5.34 | 5.06 | 4.81 | 4.58 | 4.36 | 4.15 | 3.97 | 3.77 | 3.61 | 3.44 | 3.29 | 3.14 | 3 | 2.86 | 2.74 | 2.62 | 2.51 | 2.39 | 2.29 | 2.19 | 2.09 | 2 | 1.9 | 1.82 |
| | 5.55 | 5.26 | 5 | 4.74 | 4.51 | 4.29 | 4.08 | 3.9 | 3.71 | 3.54 | 3.38 | 3.22 | 3.08 | 2.94 | 2.81 | 2.68 | 2.57 | 2.46 | 2.34 | 2.24 | 2.14 | 2.05 | 1.96 | 1.87 | 1.78 | 1.7 |
| | 5.31 | 5.04 | 4.8 | 4.55 | 4.32 | 4.11 | 3.92 | 3.73 | 3.56 | 3.39 | 3.25 | 3.09 | 2.95 | 2.83 | 2.7 | 2.58 | 2.46 | 2.35 | 2.25 | 2.15 | 2.05 | 1.96 | 1.87 | 1.79 | 1.71 | 1.63 |
| | 5.11 | 4.84 | 4.6 | 4.37 | 4.15 | 3.95 | 3.76 | 3.58 | 3.42 | 3.26 | 3.11 | 2.97 | 2.84 | 2.71 | 2.59 | 2.47 | 2.36 | 2.26 | 2.16 | 2.06 | 1.97 | 1.89 | 1.8 | 1.73 | 1.64 | 1.57 |
| | 4.81 | 4.56 | 4.34 | 4.11 | 3.91 | 3.72 | 3.55 | 3.38 | 3.22 | 3.07 | 2.94 | 2.79 | 2.68 | 2.55 | 2.44 | 2.33 | 2.23 | 2.13 | 2.03 | 1.94 | 1.86 | 1.78 | 1.7 | 1.62 | 1.55 | 1.48 |
| 0.9 | 4.63 | 4.39 | 4.17 | 3.96 | 3.77 | 3.58 | 3.41 | 3.25 | 3.1 | 2.95 | 2.82 | 2.69 | 2.57 | 2.46 | 2.35 | 2.24 | 2.14 | 2.05 | 1.96 | 1.87 | 1.79 | 1.71 | 1.63 | 1.56 | 1.49 | 1.42 |
| | 4.46 | 4.23 | 4.02 | 3.81 | 3.62 | 3.45 | 3.28 | 3.13 | 2.99 | 2.85 | 2.72 | 2.59 | 2.48 | 2.37 | 2.26 | 2.16 | 2.06 | 1.97 | 1.89 | 1.8 | 1.72 | 1.65 | 1.57 | 1.51 | 1.43 | 1.37 |
| | 4.23 | 4 | 3.81 | 3.61 | 3.43 | 3.27 | 3.11 | 2.96 | 2.83 | 2.7 | 2.58 | 2.45 | 2.35 | 2.24 | 2.14 | 2.05 | 1.95 | 1.87 | 1.79 | 1.7 | 1.63 | 1.56 | 1.49 | 1.43 | 1.36 | 1.3 |
| | 4 | 3.8 | 3.6 | 3.43 | 3.26 | 3.1 | 2.95 | 2.81 | 2.68 | 2.55 | 2.44 | 2.33 | 2.23 | 2.13 | 2.03 | 1.94 | 1.85 | 1.77 | 1.69 | 1.62 | 1.55 | 1.48 | 1.41 | 1.35 | 1.29 | 1.23 |
| | 3.87 | 3.67 | 3.49 | 3.31 | 3.15 | 3 | 2.85 | 2.72 | 2.59 | 2.47 | 2.36 | 2.25 | 2.15 | 2.06 | 1.96 | 1.87 | 1.79 | 1.72 | 1.64 | 1.56 | 1.5 | 1.43 | 1.37 | 1.31 | 1.24 | 1.19 |
| 0.8 | 3.69 | 3.49 | 3.32 | 3.15 | 3 | 2.85 | 2.71 | 2.59 | 2.47 | 2.35 | 2.25 | 2.14 | 2.05 | 1.96 | 1.86 | 1.78 | 1.7 | 1.63 | 1.56 | 1.49 | 1.42 | 1.36 | 1.3 | 1.24 | 1.18 | 1.13 |
| | 3.51 | 3.33 | 3.16 | 3 | 2.85 | 2.72 | 2.58 | 2.47 | 2.35 | 2.24 | 2.14 | 2.04 | 1.95 | 1.86 | 1.78 | 1.7 | 1.62 | 1.55 | 1.49 | 1.42 | 1.36 | 1.3 | 1.24 | 1.19 | 1.13 | 1.08 |
| | 3.35 | 3.17 | 3.02 | 2.87 | 2.73 | 2.59 | 2.47 | 2.35 | 2.24 | 2.14 | 2.05 | 1.95 | 1.86 | 1.78 | 1.7 | 1.62 | 1.55 | 1.48 | 1.42 | 1.35 | 1.29 | 1.24 | 1.18 | 1.13 | 1.08 | 1.03 |
| | 3.21 | 3.03 | 2.89 | 2.74 | 2.61 | 2.48 | 2.36 | 2.25 | 2.14 | 2.04 | 1.95 | 1.86 | 1.78 | 1.7 | 1.62 | 1.55 | 1.48 | 1.42 | 1.36 | 1.29 | 1.24 | 1.18 | 1.13 | 1.08 | 1.03 | 0.98 |
| | 3.06 | 2.91 | 2.76 | 2.62 | 2.49 | 2.37 | 2.25 | 2.15 | 2.05 | 1.95 | 1.87 | 1.78 | 1.7 | 1.63 | 1.55 | 1.48 | 1.42 | 1.36 | 1.3 | 1.24 | 1.18 | 1.13 | 1.08 | 1.04 | 0.99 | 0.94 |
| 0.7 | 2.9 | 2.75 | 2.61 | 2.48 | 2.35 | 2.24 | 2.13 | 2.03 | 1.94 | 1.85 | 1.77 | 1.68 | 1.61 | 1.54 | 1.47 | 1.4 | 1.34 | 1.28 | 1.22 | 1.17 | 1.12 | 1.07 | 1.02 | 0.98 | 0.93 | 0.89 |
| | 2.78 | 2.64 | 2.5 | 2.38 | 2.26 | 2.15 | 2.05 | 1.95 | 1.86 | 1.77 | 1.7 | 1.62 | 1.55 | 1.48 | 1.41 | 1.35 | 1.28 | 1.23 | 1.18 | 1.12 | 1.07 | 1.03 | 0.98 | 0.94 | 0.89 | 0.85 |
| | 2.64 | 2.5 | 2.38 | 2.25 | 2.15 | 2.04 | 1.94 | 1.85 | 1.77 | 1.68 | 1.61 | 1.53 | 1.47 | 1.4 | 1.34 | 1.28 | 1.22 | 1.17 | 1.12 | 1.06 | 1.02 | 0.97 | 0.93 | 0.89 | 0.85 | 0.81 |
| | 2.51 | 2.38 | 2.26 | 2.14 | 2.04 | 1.94 | 1.85 | 1.76 | 1.68 | 1.6 | 1.53 | 1.46 | 1.39 | 1.33 | 1.27 | 1.21 | 1.16 | 1.11 | 1.06 | 1.01 | 0.97 | 0.93 | 0.88 | 0.85 | 0.81 | 0.77 |
| | 2.39 | 2.26 | 2.15 | 2.04 | 1.94 | 1.85 | 1.76 | 1.68 | 1.6 | 1.52 | 1.46 | 1.39 | 1.33 | 1.27 | 1.21 | 1.16 | 1.1 | 1.06 | 1.01 | 0.96 | 0.92 | 0.88 | 0.84 | 0.81 | 0.77 | 0.73 |
| 0.6 | 2.28 | 2.16 | 2.05 | 1.95 | 1.85 | 1.76 | 1.68 | 1.6 | 1.53 | 1.45 | 1.39 | 1.32 | 1.27 | 1.21 | 1.15 | 1.1 | 1.05 | 1.01 | 0.96 | 0.92 | 0.88 | 0.84 | 0.8 | 0.77 | 0.73 | 0.7 |
| | 2.15 | 2.04 | 1.94 | 1.84 | 1.75 | 1.67 | 1.59 | 1.51 | 1.44 | 1.37 | 1.31 | 1.25 | 1.2 | 1.14 | 1.09 | 1.04 | 1 | 0.95 | 0.91 | 0.87 | 0.83 | 0.79 | 0.76 | 0.73 | 0.69 | 0.66 |
| | 2.06 | 1.95 | 1.86 | 1.76 | 1.68 | 1.6 | 1.52 | 1.45 | 1.38 | 1.32 | 1.26 | 1.2 | 1.14 | 1.09 | 1.04 | 1 | 0.95 | 0.91 | 0.87 | 0.83 | 0.8 | 0.76 | 0.73 | 0.7 | 0.66 | 0.63 |
| | 1.96 | 1.85 | 1.76 | 1.67 | 1.59 | 1.52 | 1.44 | 1.37 | 1.31 | 1.25 | 1.19 | 1.14 | 1.09 | 1.04 | 0.99 | 0.95 | 0.91 | 0.87 | 0.83 | 0.79 | 0.76 | 0.72 | 0.69 | 0.66 | 0.63 | 0.6 |
| | 1.85 | 1.75 | 1.66 | 1.57 | 1.5 | 1.43 | 1.36 | 1.29 | 1.23 | 1.18 | 1.13 | 1.07 | 1.02 | 0.98 | 0.93 | 0.89 | 0.85 | 0.82 | 0.78 | 0.74 | 0.71 | 0.68 | 0.65 | 0.62 | 0.59 | 0.57 |
| 0.5 | 1.74 | 1.65 | 1.57 | 1.49 | 1.42 | 1.35 | 1.28 | 1.22 | 1.17 | 1.11 | 1.06 | 1.01 | 0.97 | 0.93 | 0.88 | 0.84 | 0.81 | 0.77 | 0.74 | 0.7 | 0.67 | 0.64 | 0.61 | 0.59 | 0.56 | 0.53 |

95% SI < 1
95% SI > 1
SI crosses 1
